# Evaluation of Machine Learning Models for Early Prediction of Gestational Diabetes Using Retrospective Electronic Health Records from Current and Previous Pregnancies

**DOI:** 10.1101/2025.05.12.25327431

**Authors:** Mark Germaine, Amy C O’Higgins, Brendan Egan, Graham Healy

## Abstract

**Objective:** To assess the performance of machine learning (ML) models in predicting gestational diabetes mellitus (GDM) using electronic health record (EHR) data from the first antenatal visit, and determine whether incorporating previous pregnancies data improves performance.

**Methods and Analysis:** In this retrospective cohort study, several ML models were developed to predict GDM using EHR data (n=27,561, GDM 11.6%), from nulliparous and multiparous populations. Past pregnancy data (n=4,005) were incorporated to improve future (preconception) GDM predictions. Four ML algorithms were evaluated: logistic regression (LR), random forest (RF), XGBoost (XGB), and explainable boosting machine (EBM). Model performance was evaluated on an internal validation set assessing model discrimination (AUROC) and model calibration (plots, slope and intercept).

**Results:** The Feature Agnostic Model (all features) achieved AUROC 0.832 (slope 0.967; intercept −0.088) with LR, similar to more complex models such as XGB (AUROC 0.828; slope 0.976; intercept −0.072) and EBM (AUROC 0.829; slope 0.939; intercept −0.131). The Sequential Model that included first trimester and previous pregnancy data demonstrated the highest predictive performance, with XGB achieving an AUROC 0.904 (slope 0.618; intercept −0.136). Subset models using top clinical features maintained strong performance, particularly with the Sequential Model achieving an AUROC 0.897 (slope 1.137; intercept 0.161) using only eight features.

**Conclusion:** Incorporating previous pregnancy data improved ML performance for GDM prediction. Further, using a subset of clinically relevant features yielded similar performance, supporting potential integration into clinical decision support systems. These findings highlight the promise of early GDM risk identification in both nulliparous and multiparous populations; however, additional research, including external validation and clinical trials, is needed to determine the models’ practical utility and effect on maternal and neonatal outcomes.

**KEY MESSAGES:** **What is already known on this topic?**

Machine learning (ML) approaches have been used to identify gestational diabetes mellitus (GDM) risk in early pregnancy, but most studies focus only on data from the current pregnancy, and do not leverage prior obstetric history.

**What this study adds**

Incorporating data from previous pregnancies substantially improved ML models’ predictive performance, and reducing the models to a small set of key features still yielded good performance, assessed by model discrimination and calibration, suggesting feasibility for real-time clinical use at or even before the first antenatal visit.

**How this study might affect research, practice or policy**

These findings support the possibility of earlier GDM detection and intervention, and highlight the need for external validation and prospective trials to confirm broader utility, inform clinical workflows, and guide policy on integrating ML-driven risk prediction into routine maternity care.

## INTRODUCTION

Gestational diabetes mellitus (GDM) is a form of diabetes presenting during pregnancy[1]. A 2018 Lancet series[2–4] underscored that lifestyle modifications initiated early in pregnancy could influence maternal and neonatal outcomes. However, the efficacy of such lifestyle interventions remains unclear, with several strategies offering limited benefits in terms of dietary[5,6] and physical activity interventions[7–9], possibly due to the intervention’s commencement time, which is often during late stage pregnancy. Two meta-analyses[10,11] on lifestyle interventions during pregnancy highlight this point: the timing of intervention is key. Lifestyle changes initiated during the first trimester were most effective in reducing GDM risk and improving maternal health. Furthermore, early interventions are expected to provide significant cost-saving benefits for healthcare systems by reducing complications associated with GDM[12].

Machine learning (ML) offers a promising solution for the early identification of women at risk of GDM[13], addressing a key challenge of late stage (24-28 week) diagnosis. By leveraging large datasets, such as electronic health records (EHRs), ML creates predictive models that can identify women at increased risk for GDM before the traditional screening window[14]. By identifying high-risk individuals at antenatal visit, or preconception [15], ML models could facilitate earlier lifestyle and clinical interventions. Such an approach could optimise the timing of interventions, potentially enhancing delivery outcomes and improving the health of both mother and child[16,17].

Despite the growing evidence on ML models for GDM prediction[18], most studies using EHRs to date have been conducted in Asian populations[19], often yielding promising but population-specific results. Furthermore, most have primarily utilised data from the current pregnancy, with limited incorporation of past pregnancy information. One study incorporated data from a previous pregnancy to predict GDM, but it involved only East Asian women with prior GDM (n=553) and also used a current first-trimester glucose measurement[20]. These constraints underscore the need for broader examinations of whether including previous pregnancy data can enhance early GDM prediction across diverse populations, including those in Europe, and whether such models can generalise beyond women already known to be at high risk.

Therefore, the aim of this study was to develop and evaluate the performance of ML models in predicting GDM using EHR data collected in the first trimester and to determine whether incorporating data from previous pregnancies could improve predictive performance. By including data from past pregnancies, the aim was to evaluate the potential for predicting GDM risk even before conception, thereby allowing for preconception risk assessment. Additionally, by developing models tailored to both nulliparous and multiparous populations, the study explored potential differences in performance while keeping in mind future applications in clinical decision support systems (CDSS)[21]. Finally, this study also investigated using a reduced set of clinically relevant features, recognising that limiting the feature set (variables) to be collected could enhance the practicality of model deployment by clinicians in a healthcare facility.

## METHODS AND ANALYSIS

### Study Design and Population

This retrospective cohort study employed ML techniques on EHRs from The Coombe Hospital, Dublin, spanning a five-year period (2018–2022). Ethical approval was granted by The Coombe Hospital Research Ethics Committee (Study No. 06–2023). The primary aim was to develop ML models that could predict the diagnosis of GDM by using EHR data collected during the first antenatal visit (∼12^th^ week of gestation). Three distinct cohorts were used to build the ML models (see Study Populations). The study was designed following the TRIPOD+AI guidelines for reporting clinical prediction models [22]. Patients or members of the public were not involved in the design, conduct, reporting, or dissemination plans of this study due to its retrospective nature.

### Inclusion and Exclusion Criteria

Eligible records were from women were 18 years or older, attended an antenatal visit at or before 16 weeks, with complete EHR data up to 16 weeks, and validated GDM status. Exclusions included women with pre-existing diabetes (type 1 or 2), first antenatal visit after 16 weeks, those with missing or incomplete data for critical variables, invalidated GDM status, and pregnancies from 2020 (see below).

### Study Populations

Three distinct study populations were defined. The Feature Agnostic Model (FAM) included all eligible pregnancies with data up to the 16^th^ week of gestation. The Nulliparous Model (NPM), a subset of the general population, focused on women experiencing their first pregnancy. Finally, the Sequential Pregnancy Model (SPM) identified women with at least one previous pregnancy in the dataset, allowing for the inclusion of historical data to predict GDM in subsequent pregnancies.

### Data Source and Validation

Data were routinely collected by trained midwives using standardized questionnaires and entered into the hospital’s electronic health system, “Euroking K2” (Euroking Maternity Software Systems, UK)[23]. GDM was diagnosed according the International Association of Diabetes and Pregnancy Study Groups (IADPSG) criteria[1]. EHR entries indicating “Diabetes developed during pregnancy” were labelled as GDM, while all others were labelled non-GDM. GDM status was cross-referenced with a real-time clinical database (2018–2022)[24]. Records were included only if both databases provided consistent labels. Data from 2020 were excluded due to COVID-19-related fluctuations in GDM detection[24,25]. The final dataset included 27,561 pregnancies and 108 retained variables. This sample size was deemed sufficient based on the BMJ guidelines for calculating the required sample size for developing a clinical prediction model[26].

### Data Cleaning and Preprocessing

Data quality was ensured through extensive cleaning and preprocessing. Critical variables included maternal age, body mass index (BMI) at booking, key medical history items (e.g., family history of diabetes, endocrine problems), and obstetric history (e.g., previous GDM, parity). Pregnancy records missing any critical variables were excluded, and features with more than 20% missing data were removed unless the missing value could be interpreted as a null finding. Inconsistencies (e.g., negative BMI) were either corrected if plausible or excluded if not. Categorical variables were simplified; for instance, the “Cardiac Problems” feature, initially containing over 2,600 string values, was simplified into a binary “NO”/“YES,” with nulls interpreted as “NO” after clinician consultation. Any remaining missing data were then removed.

Two deviations from this general approach were the “Occupation of the Mother” mapped to the International Standard Classification of Occupations (ISCO)[27] and grouped into skill level categories, with 4 representing the highest skill level and 0 representing unemployment. Second, birthweight percentiles were calculated using the weight of the baby and the gestational age at delivery, based on the percentile ranges presented by Nicolaides et al[28].

For sequential pregnancies, two additional features, Inter-Pregnancy Interval (days from the delivery date of one pregnancy to the conception date of the next) and Inter-Pregnancy Weight Gain (difference in maternal booking weight between successive pregnancies), were calculated.

### Model Development

Four ML models were developed to address clinically relevant different aspects of the data: (1) a FAM with all available data up to week 16 of gestation, (2) a Feature Subset Model using the most important features identified across multiple ML models (reported in Table 1), (3) a NPM for nulliparous pregnancies, and (4) a SPM that incorporated previous pregnancy variables to predict GDM in subsequent pregnancies. The latter also included variations using data up to the first trimester in the current pregnancy. Model cohort information is presented in Table 1 and a list of all variables in the EHR are reported in the supplementary files. The chosen ML algorithms were Random Forest Classifier (RF), Logistic Regression (LR), XGBoost Classifier (XGB), and Explainable Boosting Machine (EBM). LR has historically been popular in GDM modelling [18], but recent evidence suggests that advanced models like RF and XGB yield better results[29,30]. EBMs have been shown to match the performance of these advanced models while retaining interpretability[31]. A Dummy Classifier from scikit-learn was included as a baseline to establish a minimal threshold for performance.

**Table 1.** Model development sets.

| Model Development Set | Data Used | No. Pregnancies | No. Predictors | GDM % |
| --- | --- | --- | --- | --- |
| Feature Agnostic Model | EHR data up to the 16<sup>th</sup> week of pregnancy | 27,561 | 79 | 11.6 |
| Feature Subset Model | Top 9 features from EHRs | 27,561 | 9 | 11.6 |
| Nulliparous Model | 1<sup>st</sup> pregnancy data, up to the 16<sup>th</sup> week of pregnancy | 11,623 | 79 | 9.9 |
| Sequential Pregnancy Model | Past pregnancy data (incl. birth outcomes) | 4,005 | 95 | 12.2 |
| SPM with 1<sup>st</sup> Trimester | Past & current pregnancy data | 4,005 | 176 | 12.2 |
| SPM Subset Model | Past & current pregnancy data | 4,005 | 8 | 12.2 |
| **Variables used for Feature Subset Model:** Ethnicity, Family History of Diabetes (binary), Previous History of Diabetes (binary), Endocrine Problems (PCOS/thyroid conditions, binary), Parity (integer), Age (integer), BMI (float), Blood Pressure (float). |  |  |  |  |
| **Variables used for SPM Subset Model:** Ethnicity, Age (integer), Interpregnancy weight change (float), History of GDM (binary), BMI (float), Time between pregnancies (integer), Birthweight Percentile (float), Family history of diabetes (binary) |  |  |  |  |

### Data Preprocessing

A pipeline was developed for data preprocessing and modelling. Categorical variables underwent one-hot encoding with the first category dropped, and numerical variables were standardized using StandardScaler. Scikit-learn’s ColumnTransformer ensured separate treatment of categorical and numerical data, and the processed DataFrame retained patient identifiers to facilitate group-based splitting. To avoid data leakage, the dataset was split into training, validation, and test sets using GroupShuffleSplit, ensuring all records from the same patient were allocated to the same split. An 80–10-10 initial split created a training set (80%) and a validation (10%) and test (10%) set. This approach ensured patient-specific splits, providing separate data for training, hyperparameter tuning, and model evaluation.

### Model Training

Hyperparameter tuning using RandomizedSearchCV with a stratified k-fold cross-validation strategy was used for each model. This approach involved a randomized search over specified parameter ranges (see Supplementary Table 1) with 10 iterations, evaluating model performance on multiple splits of the training set. Internal validation occurred at three stages: (1) 5-fold cross-validation during RandomizedSearchCV, (2) additional evaluation on the 10% validation set, and (3) final testing on the reserved 10% test set. External validation was attempted but we were unable to source an external dataset[19]. The final hyperparameter space for each model is reported in Supplementary Table 5.

### Model Performance Evaluation

The performance of the ML models were primarily evaluated, on the test set, using the Area Under the Receiver Operating Characteristic Curve (AUROC) to measure discrimination with bootstrapping (1,000 iterations) employed to compute 95% confidence intervals for the AUC, each created by sampling, with replacement, the same number of observations as the original test set[32]. Model calibration was assessed both qualitatively with the use of calibration plots, and quantitatively, with the use of slope, intercept and observed to expected ratio (O:E ratio), as has been recommended for clinical prediction models [33]. AUROC measures the model’s ability to distinguish between classes without being sensitive to class imbalances[34], while calibration assesses the agreement between the observed and predicted outcomes. Model performance was further assessed using the average precision score (AP), confusion matrices, model sensitivity, specificity and F-1 score with a default threshold of 0.5 for determining the binary classification from the classifier output, while Brier score was calculated to measure agreement between predicted probabilities and observed outcomes across all models[35]. Further, to determine whether the models’ predictive capabilities generalised across diverse patient backgrounds, performance metrics were also stratified by ethnicity.

### Addressing Class Imbalance

The dataset exhibited class imbalance, with a GDM rate of 11.6%. Pilot experiments with the Synthetic Minority Over-sampling Technique did not yield improvements, and adjusting class weights resulted in a bias toward higher false positives compared to false negatives. This trade-off was considered undesirable for clinical application. Therefore, no further techniques were applied.

### Feature Importance and Interpretability

Feature importance was determined after each model had been fully trained on the training set, ensuring that no information from the test set influenced model parameters. Relative importance scores from RF (feature_importances_) and coefficient-derived odds ratios from LR provided basic importance measures. In XGB, SHapley Additive exPlanations (SHAP) summarised each feature’s contribution to predictions, while EBM’s built-in interpretability tools offered global and per-feature visualisations (interpret package). Although SHAP values were computed on the test data for final interpretability assessments, all feature importance metrics reflected the model parameters learned from the training set only.

### Software and Computational Resources

Model development were conducted in Python 3.8.8, with pandas (1.2.4) for data manipulation, numpy (1.23.5), scikit-learn (1.2.1) for ML workflows, seaborn (0.11.1) and matplotlib (3.3.4) for visualisation, xgboost (1.7.6), shap (0.41.0), and interpretML (0.3.0). Computations were carried out on an Apple M1 workstation with 16 GB RAM. Training times ranged from several minutes models (e.g., LR) to longer durations for EBM.

## RESULTS

### Population Characteristics

Initially 37,651 pregnancy records were identified, of which 10,090 were excluded (2,696 for missing/incomplete data and 7,394 from 2020), resulting in N = 27,561 for analysis. Table 2 presents the baseline characteristics of the study participants across these populations, including key demographics, medical history, and clinical measurements. Further details of baseline characteristics by ethnicity are provided in the Supplementary Table 2.

**Table 2:** Patient characteristics of the validated dataset.

| Characteristics | N=27,561 (%) | Rate of GDM (%) |
| --- | --- | --- |
| **Ethnicity** |  |  |
| Caucasian | 24,180 (87.8) | 10.0 |
| Black | 554 (2.0) | 14.3 |
| Southeast Asian | 1,360 (4.9) | 33.9 |
| Other | 824 (3.0) | 13.7 |
| Asian | 489 (1.8) | 21.7 |
| Middle Eastern | 154 (0.6) | 13.6 |
| **Family history of Diabetes** |  |  |
| No | 21,154 (76.7) | 8.4 |
| Yes | 6,407 (23.3) | 21.9 |
| **Previous History of GDM** |  |  |
| No | 26,483 (96.1) | 9.5 |
| Yes | 1,078 (3.9) | 62.5 |
| **Other Endocrine Problems** |  |  |
| No | 21,707 (78.8) | 8.5 |
| Yes | 5,854 (21.2) | 22.8 |
| **Current GDM** |  |  |
| No | 24,373 (88.4) | 0 |
| Yes | 3,188 (11.6) | 100 |
| **Parity** |  |  |
| 0 | 11,377 (41.3) | 9.9 |
| 1 | 9,875 (35.8) | 11.4 |
| >=2 | 6,309 (22.9) | 15.0 |
| **Age** |  |  |
| Age (Mean ± SD) | 32 ± 5 | |
| >=40 | 2,178 (7.9) | 17.2 |
| <40 | 25,383 (92.1) | 11.1 |
| **BMI** |  |  |
| BMI (Mean ± SD) | 26.2 ± 5.3 | |
| <25 | 13,695 (49.7) | 4.9 |
| 25 to <30 | 8,423 (30.6) | 11.6 |
| 30 to <35 | 3,537 (12.8) | 24.2 |
| 35 to <40 | 1,280 (4.6) | 33.0 |
| >=40 | 626 (2.3) | 41.9 |
| **Blood Pressure** |  |  |
| Systolic BP (Mean ± SD) | 111 ± 11 | |
| Diastolic BP (Mean ± SD) | 67 ± 8 | |

### GDM Prediction in the First Trimester

In the FAM, the LR model achieved the highest AUROC of 0.832 (0.810–0.853) (Figure 1A), calibration slope 0.967, intercept −0.088 and an AP of 0.449 (Figure 1C). The sensitivity was 0.207, specificity was 0.982, F1 score was 0.309, and the Brier score was 0.083. Other models showed similar performance, with XGB achieving an AUROC of 0.828 (0.807– 0.850) (AP 0.438) (Figure 1B), and EBM AUROC of 0.829 (0.806–0.850).

**Figure 1.**
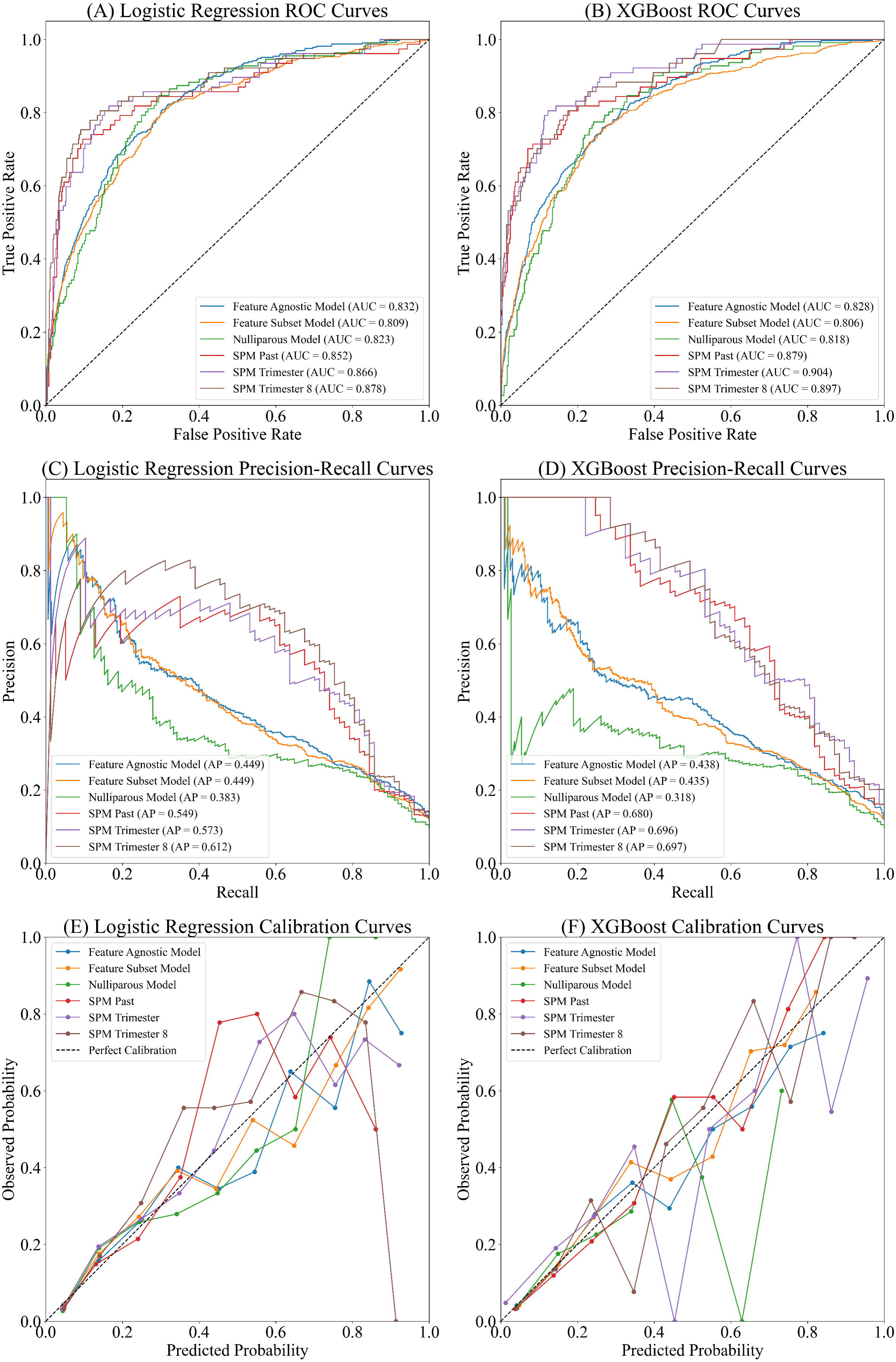
Model performance and calibration for Logistic Regression (left panel) and XGBoost (right panel). Subplots A, C, and E show the AUROC, average precision, and calibration plots, respectively, for the Logistic Regression models, while subplots B, D, and F show the corresponding plots for the XGBoost models.

For the Subset Model, LR maintained an AUROC of 0.809 (0.788–0.829) (Figure 1A), calibration slope 0.978, intercept −0.006 and an AP of 0.449 (Figure 1C) using only the top 9 features (Table 2). The sensitivity was 0.209, specificity was 0.983, F1 score was 0.314, and the Brier score was 0.087. An interactive example of the model can be found here ([ML Labs Tool](https://ml-labs-gdm-prediction-tool.streamlit.app/)).

In the NPM, LR again performed strongly, achieving an AUROC of 0.823 (0.785– 0.859) (Figure 1A), calibration slope 0.967, intercept −0.088, and an AP of 0.383 (Figure 1C). The sensitivity was 0.126, specificity was 0.991, F1 score was 0.209, and the Brier score was 0.074. Performance metrics for each model across different datasets are summarised in Table 3.

**Table 3.**
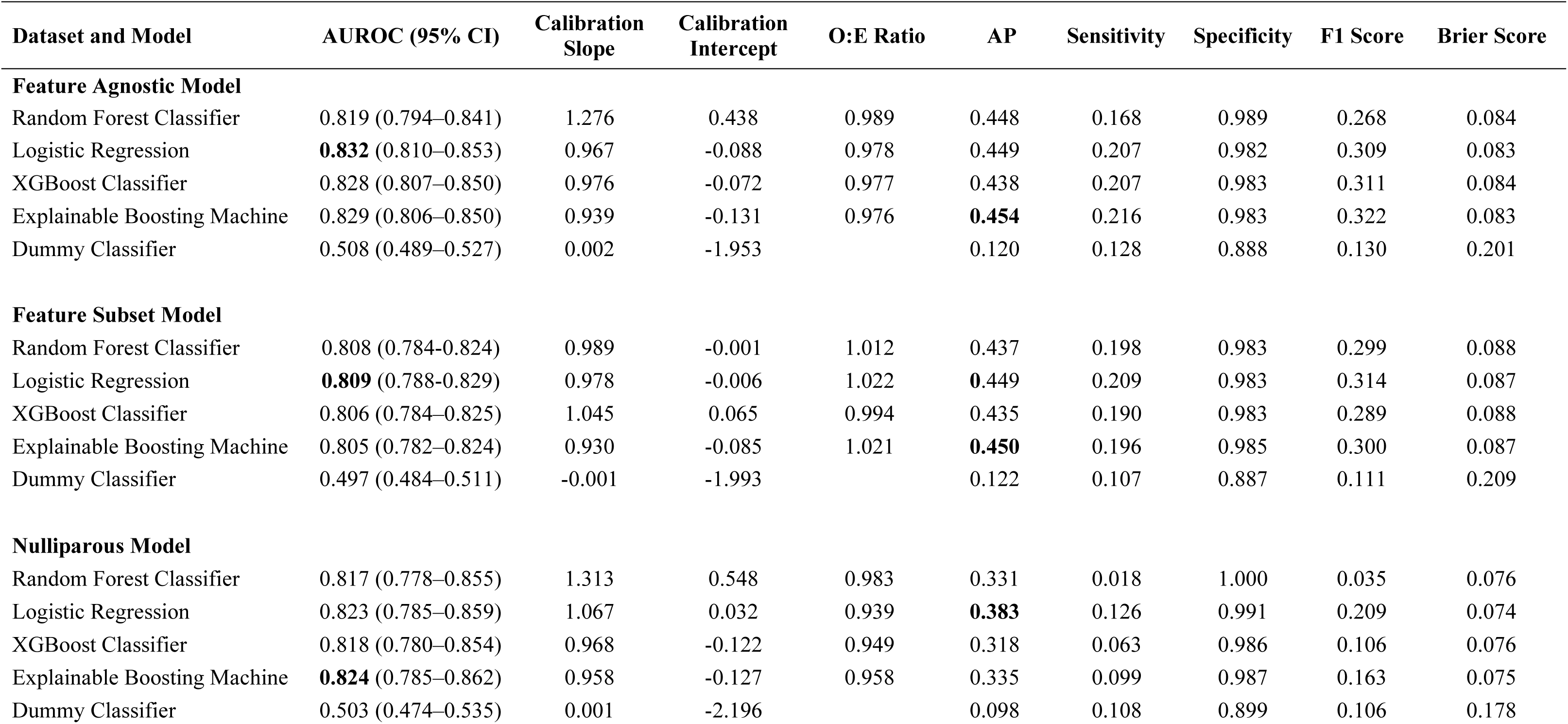
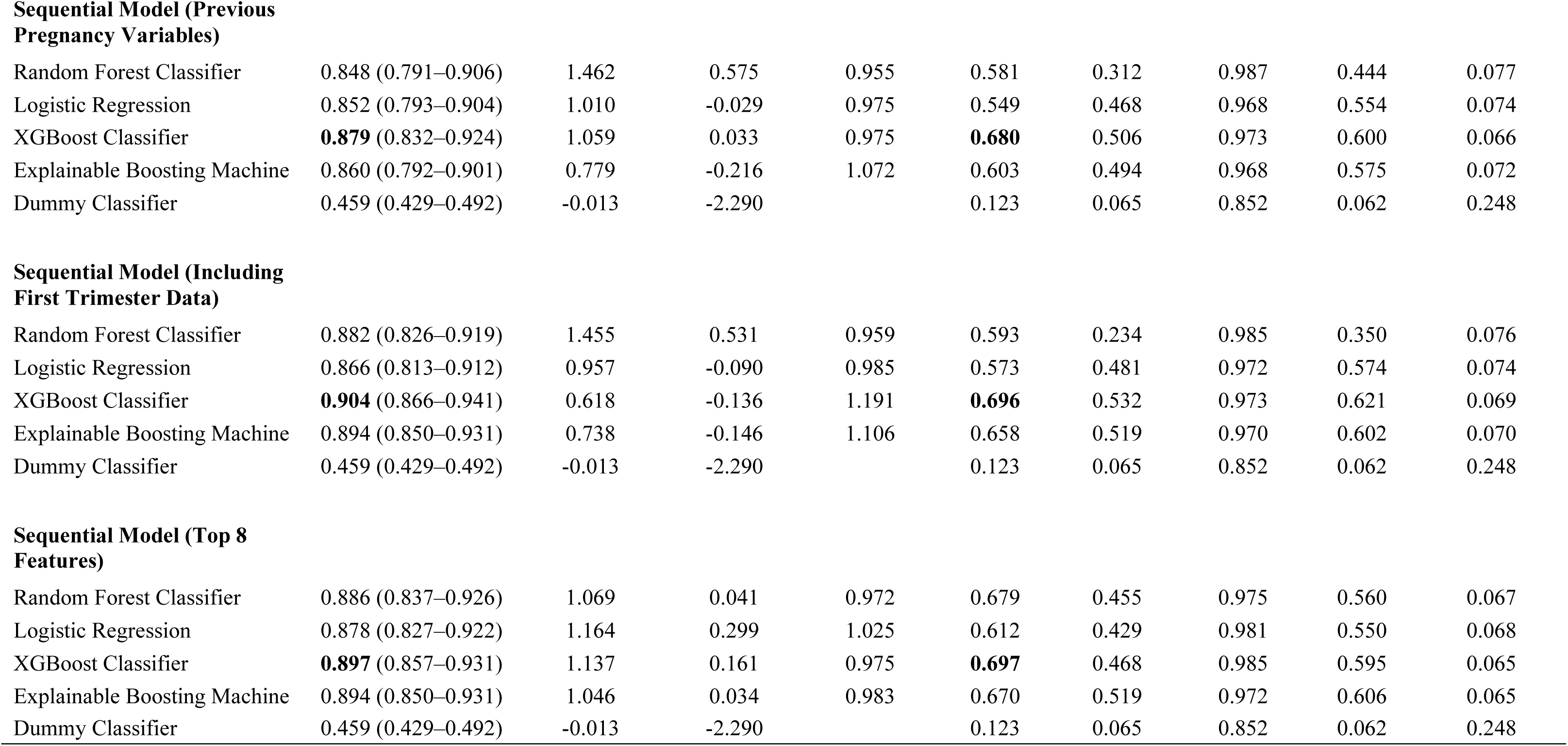
Comparative Performance Metrics of Machine Learning Models Evaluated on the Test Set Across Various Datasets. AUROC (Area Under the Receiver Operating Characteristic Curve), AP (Average Precision Score), O:E Ratio (Observed to Expected Ratio).

| Dataset and Model | AUROC (95% CI) | Calibration Slope | Calibration Intercept | O:E Ratio | AP | Sensitivity | Specificity | F1 Score | Brier Score |
| --- | --- | --- | --- | --- | --- | --- | --- | --- | --- |
| **Feature Agnostic Model** |  |  |  |  |  |  |  |  |  |
| Random Forest Classifier | 0.819 (0.794–0.841) | 1.276 | 0.438 | 0.989 | 0.448 | 0.168 | 0.989 | 0.268 | 0.084 |
| Logistic Regression | **0.832** (0.810–0.853) | 0.967 | -0.088 | 0.978 | 0.449 | 0.207 | 0.982 | 0.309 | 0.083 |
| XGBoost Classifier | 0.828 (0.807–0.850) | 0.976 | -0.072 | 0.977 | 0.438 | 0.207 | 0.983 | 0.311 | 0.084 |
| Explainable Boosting Machine | 0.829 (0.806–0.850) | 0.939 | -0.131 | 0.976 | **0.454** | 0.216 | 0.983 | 0.322 | 0.083 |
| Dummy Classifier | 0.508 (0.489–0.527) | 0.002 | -1.953 |  | 0.120 | 0.128 | 0.888 | 0.130 | 0.201 |
| **Feature Subset Model** |  |  |  |  |  |  |  |  |  |
| Random Forest Classifier | 0.808 (0.784-0.824) | 0.989 | -0.001 | 1.012 | 0.437 | 0.198 | 0.983 | 0.299 | 0.088 |
| Logistic Regression | **0.809** (0.788-0.829) | 0.978 | -0.006 | 1.022 | **0.449** | 0.209 | 0.983 | 0.314 | 0.087 |
| XGBoost Classifier | 0.806 (0.784–0.825) | 1.045 | 0.065 | 0.994 | 0.435 | 0.190 | 0.983 | 0.289 | 0.088 |
| Explainable Boosting Machine | 0.805 (0.782–0.824) | 0.930 | -0.085 | 1.021 | **0.450** | 0.196 | 0.985 | 0.300 | 0.087 |
| Dummy Classifier | 0.497 (0.484–0.511) | -0.001 | -1.993 |  | 0.122 | 0.107 | 0.887 | 0.111 | 0.209 |
| **Nulliparous Model** |  |  |  |  |  |  |  |  |  |
| Random Forest Classifier | 0.817 (0.778–0.855) | 1.313 | 0.548 | 0.983 | 0.331 | 0.018 | 1.000 | 0.035 | 0.076 |
| Logistic Regression | 0.823 (0.785–0.859) | 1.067 | 0.032 | 0.939 | **0.383** | 0.126 | 0.991 | 0.209 | 0.074 |
| XGBoost Classifier | 0.818 (0.780–0.854) | 0.968 | -0.122 | 0.949 | 0.318 | 0.063 | 0.986 | 0.106 | 0.076 |
| Explainable Boosting Machine | **0.824** (0.785–0.862) | 0.958 | -0.127 | 0.958 | 0.335 | 0.099 | 0.987 | 0.163 | 0.075 |
| Dummy Classifier | 0.503 (0.474–0.535) | 0.001 | -2.196 |  | 0.098 | 0.108 | 0.899 | 0.106 | 0.178 |
| Sequential Model (Previous Pregnancy Variables) |  |  |  |  |  |  |  |  |  |
| Random Forest Classifier | 0.848 (0.791–0.906) | 1.462 | 0.575 | 0.955 | 0.581 | 0.312 | 0.987 | 0.444 | 0.077 |
| Logistic Regression | 0.852 (0.793–0.904) | 1.010 | -0.029 | 0.975 | 0.549 | 0.468 | 0.968 | 0.554 | 0.074 |
| XGBoost Classifier | **0.879** (0.832–0.924) | 1.059 | 0.033 | 0.975 | **0.680** | 0.506 | 0.973 | 0.600 | 0.066 |
| Explainable Boosting Machine | 0.860 (0.792–0.901) | 0.779 | -0.216 | 1.072 | 0.603 | 0.494 | 0.968 | 0.575 | 0.072 |
| Dummy Classifier | 0.459 (0.429–0.492) | -0.013 | -2.290 |  | 0.123 | 0.065 | 0.852 | 0.062 | 0.248 |
| Sequential Model (Including First Trimester Data) |  |  |  |  |  |  |  |  |  |
| Random Forest Classifier | 0.882 (0.826–0.919) | 1.455 | 0.531 | 0.959 | 0.593 | 0.234 | 0.985 | 0.350 | 0.076 |
| Logistic Regression | 0.866 (0.813–0.912) | 0.957 | -0.090 | 0.985 | 0.573 | 0.481 | 0.972 | 0.574 | 0.074 |
| XGBoost Classifier | **0.904** (0.866–0.941) | 0.618 | -0.136 | 1.191 | **0.696** | 0.532 | 0.973 | 0.621 | 0.069 |
| Explainable Boosting Machine | 0.894 (0.850–0.931) | 0.738 | -0.146 | 1.106 | 0.658 | 0.519 | 0.970 | 0.602 | 0.070 |
| Dummy Classifier | 0.459 (0.429–0.492) | -0.013 | -2.290 |  | 0.123 | 0.065 | 0.852 | 0.062 | 0.248 |
| Sequential Model (Top 8 Features) |  |  |  |  |  |  |  |  |  |
| Random Forest Classifier | 0.886 (0.837–0.926) | 1.069 | 0.041 | 0.972 | 0.679 | 0.455 | 0.975 | 0.560 | 0.067 |
| Logistic Regression | 0.878 (0.827–0.922) | 1.164 | 0.299 | 1.025 | 0.612 | 0.429 | 0.981 | 0.550 | 0.068 |
| XGBoost Classifier | **0.897** (0.857–0.931) | 1.137 | 0.161 | 0.975 | **0.697** | 0.468 | 0.985 | 0.595 | 0.065 |
| Explainable Boosting Machine | 0.894 (0.850–0.931) | 1.046 | 0.034 | 0.983 | 0.670 | 0.519 | 0.972 | 0.606 | 0.065 |
| Dummy Classifier | 0.459 (0.429–0.492) | -0.013 | -2.290 |  | 0.123 | 0.065 | 0.852 | 0.062 | 0.248 |

### Sequential GDM Prediction

The sequential models using only previous pregnancy variables showed improved performance. XGB achieved the highest AUROC of 0.879 (0.832–0.924) (Figure 1B), calibration slope 1.059, intercept 0.033, and an AP of 0.680 (Figure 1D), with a sensitivity of 0.537, specificity of 0.963, F1 score of 0.604, and Brier score of 0.070.

When including first trimester data in the SPM, XGB achieved an AUROC of 0.904 (0.866–0.941) (Figure 1B), calibration slope 0.681, intercept −0.136, and an AP of 0.696 (Figure 1D). The sensitivity increased to 0.532, specificity was 0.973, F1 score was 0.621, and the Brier score improved to 0.069.

In the SPM focusing on the top 8 features (identified from the training data), XGB achieved the highest AUROC of 0.897 (0.857 – 0.931)(Figure 1B), calibration slope 1.137, intercept 0.161, and an AP of 0.697 (Figure 1D). The sensitivity was 0.468, specificity was 0.985, F1 score was 0.595, and the Brier score was 0.065. All models are presented in Table 3.

Calibration plots for the LR and XGB models are presented in Figure 1E and 1F, showing that the predicted probabilities closely align with observed outcomes. The Brier scores ranged from 0.063 to 0.088 across all models, indicating good calibration, with predicted probabilities closely aligning with observed outcomes.

### Model Performance Across Ethnicities

Performance metrics stratified by ethnicity are shown in Supplementary Table 3. Generally, the models maintained performance for Caucasian patients (AUROC >0.8), with results closely tracking those of the full dataset. However, there was a consistent reduction in AUROCs for Asian, Southeast Asian, and Black populations (<0.8), indicating reduced predictive power in these smaller or more diverse subgroups. Conversely, the models continued to perform well among Other and Middle Eastern patients, although very small sample sizes may have skewed these results.

### Feature Importance

The feature importance analysis identified the top predictors of GDM, including a history of GDM, maternal BMI at booking, maternal age, family history of diabetes, ethnicity, inter-pregnancy weight gain (in SPM), time between pregnancies (in SPM), ethnicity, and occupational skill level. The SHAP plots for the XGB are shown in Supplementary Figure 2. The feature importance for all of the models can be found in Supplementary Figures 2-5.

## DISCUSSION

This study evaluated the performance of several ML models for early prediction of GDM using data available at the first antenatal visit, and examined whether incorporating information from previous pregnancies enhances predictive performance. Overall, the findings confirmed that incorporating prior pregnancy history can improve early risk prediction for GDM. For example, the SPM that included both first-trimester and previous pregnancy features achieved an AUROC of 0.904, higher than models using first-visit data alone, suggesting that a woman’s obstetric history is highly informative for forecasting GDM in a subsequent pregnancy. Notably, even a simplified sequential model using only the top eight features achieved nearly equivalent discrimination (AUROC 0.897), highlighting that a focused set of clinical predictors can retain most of the predictive signal.

In the FAM, the LR model performed similarly to more complex models such as XGB and EBM (AUROC ∼0.832). This contrasts with much of the existing literature, highlighted in a recent meta-analysis that found LR models achieved a pooled AUROC of 0.815, compared to 0.889 for non-linear models[18]. This discrepancy suggests that more sophisticated ML algorithms tend to perform better, potentially capturing complex, non-linear relationships in the data. However, the comparisons in the meta-analysis were not always direct comparisons of model performance, as the models were often tested on different datasets, each with varying characteristics, sample sizes, and feature availability. Nevertheless, even when looking at studies that implemented LR alongside more advanced models on the same dataset the pattern of increased performance with non-linear models like XGB or RF remains evident[36–40]. The lack of a gap in LR performance may indicate that the relationships between early-pregnancy predictors and GDM are largely linear or additive, meaning a well-specified linear model can capture them adequately. It is also possible that the complex models were limited by the data quality or volume, or by the fact that only routine, non-invasive features were intentionally used. This underscores an important point for clinical machine learning, more complex is not always better, especially if interpretability and ease of use are priorities. From a clinician’s perspective, a simpler model that offers similar performance might be preferable for integration into practice, due to its transparency and reliability[41].

A strength of this study is the exclusive use of non-invasive, routinely collected data available at booking, which enhances the practicality and scalability of implementing the model in clinical settings. With the exception of height, body mass and blood pressure, all predictors in the models were questionnaire-based or existing records. Among previous studies that used non-invasive features, the majority demonstrate moderate predictive power (AUROC 0.7-0.8)[13], with two exceptions to date that demonstrate much better performance[36,42]. In comparison, our models that used non-invasive features achieved AUROCs ranging from 0.809 in the Subset Model up to 0.897 in the SPM that focussed on the top 8 features, suggesting that leveraging a patient’s obstetric history and optimising the ML methodology can substantially boost performance even without biochemical markers. Notably, the SPM with XGB and the EBM demonstrated the highest performance by achieving AUROCs of 0.904 and 0.894, respectively, which were in the range reported by Belsti et al.[36] (0.921) and Sweeting et al.[42] (0.880), and in the range of models that use biochemical predictors in addition to non-invasive features[38].

Including previous pregnancy variables in multiparous women (SPM) improved model performance compared to the FAM and NPM, with XGB achieving an AUROC of 0.884. These results align favourably with findings from other studies that emphasise the importance of early pregnancy and preconception data for GDM prediction, particularly when considering non-invasive data collection. For instance, Artzi et al.[43] utilised features accessible ‘at the beginning of pregnancy’ and achieved an AUROC of 0.799 using just nine features collected from questionnaires. It could be argued that this is a similar approach to the Feature Subset Model without blood pressure measures, which have minimal impact on the model. Additional work has demonstrated the benefit (AUROC 0.930) of incorporating biochemical markers like glycosylated haemoglobin (HbA1c) with other features collected during the preconception stage of pregnancy[15]. Thus, these data demonstrate the potential for predicting diagnosis of GDM with data available prior to conception. The inclusion of inter-pregnancy factors, such as weight gain and time interval between pregnancies, provided an additional perspective in our study, enhancing the models’ predictive power to an AUROC 0.904. This finding aligns with work demonstrating the value of including more comprehensive data in the prediction of recurrent GDM, which achieved an AUROC of 0.942 with Light Gradient Boosting and 0.924 with XGB, by incorporating biomarkers such as the OGTT results from the index pregnancy, and fasting plasma glucose (FPG) and triglycerides (TG) in the first trimester of the ongoing pregnancy[20].

The reported AUROC values in our study demonstrate consistency across some subgroups, which is important for ensuring generalisability of the models, while performing poorly across others. The NPM achieved an AUROC of 0.823, indicating promising predictive performance in identifying diagnosis of GDM among nulliparous pregnancies. This result compares favourably to that of Kang et al.[44], who reported variability in AUROC between multiparous (0.720) and nulliparous (0.672) populations, suggesting that our model was better able to consistently capture key predictors for nulliparous women. Further comparison with Cooray et al.[45] (AUROC 0.732) and Donovan et al.[46] (AUROC 0.710), both of which focused on nulliparous populations using LR, also highlights the favourable performance of our models.

Beyond parity-based analysis, we also explored model performance across ethnic subgroups (see Supplementary Table 3). While results remained consistent among Caucasian participants, performance declined in certain minority groups, particularly Asian, Southeast Asian, and Black populations. This finding is consistent with previous research in Ireland demonstrating a marked decrease in model performance in non-Caucasian populations[47]. In contrast, models continued to perform favourably for Other and Middle Eastern individuals, though the small sample sizes for these subgroups limit the generalisability of these findings. However, models trained on diverse populations in California, USA, noted the opposite trend, whereby models performed well in Hispanic populations but tended to underperform in Caucasian populations[46]. This highlights the need for including more diverse data in model training.

The feature importance analysis confirmed that a history of GDM, maternal BMI, maternal age, and ethnicity were strong predictors in the early pregnancy GDM models, and aligns with meta-analytical analysis in this field [18]. Other frequent predictors that were not available in the current feature set were FPG, TG and HbA1c. In our SPM, inter-pregnancy weight gain also emerged as a significant predictor, consistent with previous work[20]. These findings are confirmatory, as they reinforce the known clinical relevance of these features, and their effectiveness as strong predictors in different studies and populations.

In this study, we developed subset versions of each model by selecting the most clinically relevant features, aiming to enhance the potential for CDSS[21] by relying on a limited set of predictors rather than an entire EHR. Although the subset version of the FAM dropped in performance (0.832 to 0.809), the NPM (0.823 to 0.825, Supplementary Table 4) and SPM remained robust, with the latter achieving an AUROC of 0.897 using only eight features, comparable to its 176-feature counterpart. This streamlined approach could facilitate earlier GDM risk prediction, potentially as early as 12 weeks’ gestation or even preconception, thereby enabling more proactive interventions such as earlier diagnostic testing long before standard screening. However, to integrate these models seamlessly into antenatal workflows, clinicians need clear guidelines for classifying high-risk and moderate-risk patients, and the chosen thresholds must carefully balance the risks of false positives (unnecessary interventions) and false negatives (missed high-risk cases).

This study has several limitations. First, the dataset used for model development was derived from a single institution, which may limit the generalisability of the findings. External validation using data from other populations is necessary to confirm the models’ robustness, as emphasised by others[39,46,48], who validated their model performance across independent populations. The use of historical data from previous pregnancies also means that model performance may vary based on the quality and availability of such data across healthcare systems. The method by which features were pre-processed could have resulted in a loss of potentially important information. One-hot encoding broad categorical features, such as “Endocrine problems,” into binary variables may have discarded valuable context that could have improved predictions. Moreover, we did not have access to blood-based biomarkers in this study, such as FPG or HbA1c, which have been shown to be strong predictors of GDM[15]. While the goal of this study was to develop a model that could be used at the booking visit in the first trimester without the need for extensive laboratory tests, incorporating point-of-care biomarkers could further enhance model performance. Socioeconomic and educational status were also not directly available in this dataset. Previous research has shown that educational attainment, particularly in women, is correlated with health outcomes, including the risk of GDM and inter-pregnancy weight gain[49]. Therefore, we used occupation as a proxy for socioeconomic status, but this may be a weak estimate. Finally, our models did not include OGTT results, limiting predictions to the IADPSG criteria. Modelling GDM using OGTT results could provide a more versatile tool applicable to multiple diagnostic standards.

In conclusion, these ML models, particularly those incorporating data from previous pregnancies, have the potential early in pregnancy to identify women at greater risk of later diagnosis of GDM. Early identification may allow for timely interventions, which could mitigate the adverse maternal and foetal outcomes associated with GDM. Future research should focus on validating the developed models in external datasets to assess their generalisability. Incorporating additional features, such as lifestyle factors and biomarkers, may further improve model performance and sensitivity. Additionally, prospective studies evaluating the integration of these models into clinical workflows would be valuable to determine their impact on clinical decision-making and patient outcomes.

## Data Availability

Due to patient confidentiality and data use agreements, individual-level data cannot be shared publicly.

## Acknowledgments

The authors are grateful to The Coombe Hospital for their collaboration without whom this analysis would not be possible.

## Data and Resource Availability

Due to patient confidentiality and data use agreements, individual-level data cannot be shared publicly. The models are intended for research purposes and require external validation before clinical implementation.

## Code Availability

The authors will share the code and saved models in a GitHub repository or open science framework so that other researchers can reuse the models for research purposes.

## Funding and Assistance

This work has emanated from research supported in part by a grant from Research Ireland under Grant Number 18/CRT/6183.

## Author Contributions

M.G., A.C.O., B.E., and G.H. were involved in the conception, design, and conduct of the study and the analysis and interpretation of the results. M.G. wrote the first draft of the manuscript, and all authors edited, reviewed, and approved the final version of the manuscript. M.G. is the guarantor of this work and, as such, had full access to all the data in the study and takes responsibility for the integrity of the data and the accuracy of the data analysis.

## Protocol and Registration

The protocol was not prepared for this study, and the study was not pre-registered.

## Prior Presentation

Part of this work was presented in abstract form at the American Diabetes Association Annual Meeting, 21-24 June, 2024, Orlando, Florida, USA: doi.org/10.2337/db24-1968-LB

## Abbreviations

AP: Average Precision
AUROC: area under the receiver operating characteristic curve
BMI: Body mass index
CDSS: clinical decision support systems
EHR: Electronic health record
EBM: Explainable Boosting Machine
FAM: Feature Agnostic Model
FPG: fasting plasma glucose
GDM: Gestational diabetes mellitus
HbA1c: glycosylated haemoglobin
IADPSG: International Association of Diabetes and Pregnancy Study Groups
IDs: patient identifiers
LR: Logistic Regression
ML: Machine learning
NPM: Nulliparous Model
OGTT: oral glucose tolerance test
RF: Random Forest
SPM: Sequential Pregnancy Model
SHAP: SHapley Additive exPlanations
TG: triglycerides
XGB: eXtreme Gradient Boosting

## Supplementary Tables

**S Table 1.** Hyperparameter space searched for the models during training.

| Hyperparameter Search Space (n_iter = 10) |  |
| --- | --- |
| **Random Forest Classifier** | • n_estimators: [100, 200, 300]<br>• max_depth: [None, 10, 20, 30]<br>• min_samples_split: [2, 5, 10]<br>• min_samples_leaf: [1, 2, 4] |
| **Logistic Regression** | • C: np.logspace(-4, 4, 20)<br>• solver: ['liblinear', 'lbfgs'] |
| **XGB Classifier** | • n_estimators: [100, 200, 300]<br>• learning_rate: [0.01, 0.1, 0.2]<br>• max_depth: [3, 4, 5, 10] |
| **Explainable Boosting Classifier** | • learning_rate: [0.01, 0.1, 1]<br>• max_leaves: [3, 10, 20] |

**S Table 2.** Patient characteristics grouped by ethnicity expressed as number of occurrences (%).

| Characteristic | Caucasian<br>n=24,180 | Southeast Asian<br>n=1,360 | Black<br>n=554 | Asian<br>n=489 | Middle Eastern<br>n=154 | Other<br>n=824 |
| --- | --- | --- | --- | --- | --- | --- |
| **FH** |  |  |  |  |  |  |
| **Diabetes\*** |  |  |  |  |  |  |
| No | 19,161 (79.2) | 603 (44.3) | 390 (70.4) | 343 (70.1) | 83 (53.9) | 574 (69.7) |
| Yes | 5,019 (20.8) | 757 (55.7) | 164 (29.6) | 146 (29.9) | 71 (46.1) | 250 (30.3) |
| **Hx of GDM\*** |  |  |  |  |  |  |
| No | 23,385 (96.7) | 1,187 (87.3) | 518 (93.5) | 465 (95.1) | 140 (90.1) | 788 (95.6) |
| Yes | 795 (3.3) | 173 (12.7) | 36 (6.5) | 24 (4.9) | 14 (9.9) | 36 (4.4) |
| **GDM** |  |  |  |  |  |  |
| No | 21,772 (90.0) | 899 (66.1) | 475 (85.7) | 383 (78.3) | 133 (86.4) | 711 (86.3) |
| Yes | 2,408 (10.0) | 461 (33.9) | 79 (14.3) | 106 (21.7) | 21 (13.6) | 113 (13.7) |
| **Parity<sup>+</sup>** |  |  |  |  |  |  |
| 0 | 10,022 (41.4) | 511 (37.6) | 229 (41.3) | 222 (45.4) | 54 (35.1) | 406 (49.3) |
| 1 | 8,741 (36.1) | 485 (35.7) | 167 (30.1) | 173 (35.4) | 51 (33.1) | 264 (32.0) |
| ≥2 | 5,417 (22.4) | 364 (26.8) | 158 (28.5) | 94 (19.2) | 49 (31.8) | 154 (18.7) |
| **Age<sup>#</sup>** |  |  |  |  |  |  |
| Age (Mean ± SD) | 33 ± 5 | 32 ± 5 | 31 ± 6 | 33 ± 5 | 31 ± 5 | 33 ± 5 |
| ≥40 | 1,934 (8.0) | 50 (3.7) | 50 (9.0) | 51 (10.4) | 4 (2.6) | 89 (10.8) |
| <40 | 22,246 (92.0) | 1,310 (96.3) | 504 (91.0) | 438 (89.6) | 150 (97.4) | 735 (89.2) |
| **BMI** |  |  |  |  |  |  |
| BMI (Mean ± SD) | 26.2 ± 4.3 | 26.1 ± 4.6 | 28.5 ± 6.1 | 24.2 ± 4.3 | 25.8 ± 4.0 | 25.6 ± 5.1 |
| <25 | 12,092 (50.0) | 601 (44.2) | 182 (32.9) | 316 (64.6) | 76 (49.4) | 439 (53.3) |
| 25 to <30 | 7,298 (30.2) | 540 (39.0) | 173 (31.2) | 117 (23.9) | 56 (37.0) | 237 (28.8) |
| 30 to <35 | 3,081 (12.7) | 176 (12.9) | 116 (20.9) | 44 (9.0) | 18 (11.7) | 102 (12.4) |
| 35 to <40 | 1,149 (4.8) | 36 (2.6) | 52 (9.4) | 10 (2.0) | 1 (0.6) | 32 (3.9) |
| ≥40 | 560 (2.3) | 17 (1.3) | 31 (5.6) | 2 (0.4) | 2 (1.3) | 14 (1.7) |
| Systolic BP (Mean ± SD) | 111 ± 11 | 109 ± 10 | 113 ± 11 | 109 ± 11 | 107 ± 12 | 111 ± 10 |
| Diastolic BP (Mean ± SD) | 67 ± 8 | 67 ± 8 | 69 ± 9 | 66 ± 8 | 65 ± 8 | 67 ± 8 |

**S Table 3.** Performance of machine learning models predicting across the different ethnicity sub-groups. Performance measured by ROC area under the curve evaluated against the validation set combined with the test set. Minimum of 15 samples required.

| Dataset and Model | Caucasian | Southeast Asian | Black | Asian | Middle Eastern | Other |
| --- | --- | --- | --- | --- | --- | --- |
| **Feature Agnostic Model** | n=4,826 | n=282 | n=124 | n=87 | n=42 | n=165 |
| Random Forest Classifier | 0.803 | 0.718 | 0.793 | 0.705 | 0.895 | 0.884 |
| Logistic Regression | 0.814 | 0.740 | 0.804 | 0.735 | 0.934 | 0.856 |
| XGBoost Classifier | 0.810 | 0.708 | 0.782 | 0.730 | 0.895 | 0.883 |
| Explainable Boosting Machine | 0.810 | 0.724 | 0.806 | 0.730 | 0.934 | 0.876 |
| **Feature Subset Model** |  |  |  |  |  |  |
| Random Forest Classifier | 0.807 | 0.728 | 0.759 | 0.718 | 0.846 | 0.876 |
| Logistic Regression | 0.813 | 0.742 | 0.752 | 0.743 | 0.843 | 0.867 |
| XGBoost Classifier | 0.811 | 0.733 | 0.774 | 0.714 | 0.836 | 0.875 |
| Explainable Boosting Machine | 0.810 | 0.738 | 0.772 | 0.729 | 0.841 | 0.863 |
| **Nulliparous Model** | n=2,011 | n=87 | n=36 | n=32 | n=12 | n=97 |
| Random Forest Classifier | 0.804 | 0.741 | 0.250 | 0.691 | - | 0.929 |
| Logistic Regression | 0.811 | 0.738 | 0.294 | 0.686 | - | 0.902 |
| XGBoost Classifier | 0.806 | 0.718 | 0.221 | 0.662 | - | 0.885 |
| Explainable Boosting Machine | 0.815 | 0.722 | 0.324 | 0.671 | - | 0.910 |
| **Sequential Model (Previous Pregnancy Variables)** | n=1,114 | n=43 | n=21 | n=15 | n=2 | n=20 |
| Random Forest Classifier | 0.887 | 0.860 | 0.907 | 0.955 | - | 0.987 |
| Logistic Regression | 0.826 | 0.812 | 0.778 | 0.887 | - | 0.960 |
| XGBoost Classifier | 0.887 | 0.889 | 0.852 | 0.932 | - | 0.947 |
| Explainable Boosting Machine | 0.866 | 0.876 | 0.907 | 0.864 | - | 0.960 |
| **Sequential Model (Including First Trimester Data)** |  |  |  |  |  |  |
| Random Forest Classifier | 0.874 | 0.864 | 0.815 | 0.932 | - | 0.973 |
| Logistic Regression | 0.849 | 0.783 | 0.796 | 0.864 | - | 0.960 |
| XGBoost Classifier | 0.884 | 0.830 | 0.852 | 0.909 | - | 0.973 |
| Explainable Boosting Machine | 0.883 | 0.885 | 0.648 | 0.886 | - | 1.000 |
| **Sequential Model (Top 8 Features)** |  |  |  |  |  |  |
| Random Forest Classifier | 0.881 | 0.808 | 0.796 | 0.977 | - | 0.987 |
| Logistic Regression | 0.867 | 0.805 | 0.852 | 0.864 | - | 0.987 |
| XGBoost Classifier | 0.886 | 0.781 | 0.796 | 0.955 | - | 0.987 |
| Explainable Boosting Machine | 0.885 | 0.819 | 0.852 | 0.886 | - | 0.987 |

**S Table 4.** Performance of Nulliparous Model trained with the same top 9 features from the Feature Subset Model.

| Dataset and Model | AUROC | AP | Sensitivity | Specificity | F1 Score | Brier Score |
| --- | --- | --- | --- | --- | --- | --- |
| **Nulliparous Model Subset** |  |  |  |  |  |  |
| Random Forest Classifier | 0.822 | 0.377 | 0.028 | **0.997** | 0.053 | 0.078 |
| Logistic Regression | **0.825** | 0.388 | **0.106** | 0.990 | **0.178** | 0.078 |
| XGBoost Classifier | 0.824 | 0.359 | 0.050 | 0.993 | 0.091 | 0.078 |
| Explainable Boosting Machine | 0.825 | **0.390** | 0.084 | 0.995 | 0.078 | 0.078 |
| Dummy Classifier | 0.503 | 0.098 | 0.108 | 0.899 | 0.106 | 0.178 |

**S Table 5.** Final hyperparameters chosen for all models as a result of hyperparameter tuning. Only the tuned hyperparameters are displayed, all other parameters are thus default values.

| Dataset and Model | Hyperparameters |
| --- | --- |
| **Feature Agnostic Model** |  |
| Random Forest Classifier | RandomForestClassifier(max_depth=10, min_samples_leaf=4) |
| Logistic Regression | LogisticRegression(C=0.08858667904100823, max_iter=1000) |
| XGBoost Classifier | XGBClassifier(max_depth=3, n_estimators=100, learning_rate=1) |
| Explainable Boosting Machine | ExplainableBoostingClassifier() |
| **Feature Subset Model** |  |
| Random Forest Classifier | RandomForestClassifier(max_depth=10, min_samples_leaf=4) |
| Logistic Regression | LogisticRegression(C=0.23357214690901212, max_iter=1000, solver='liblinear') |
| XGBoost Classifier | XGBClassifier(learning_rate=0.01, max_depth=5, n_estimators=300) |
| Explainable Boosting Machine | ExplainableBoostingClassifier(learning_rate=0.1, max_bins=256) |
| **Nulliparous Model** |  |
| Random Forest Classifier | RandomForestClassifier(max_depth=10, min_samples_leaf=4) |
| Logistic Regression | LogisticRegression(C=0.03359818286283781, max_iter=1000, solver='liblinear') |
| XGBoost Classifier | XGBClassifier(max_depth=3, n_estimators=100, learning_rate=0.1) |
| Explainable Boosting Machine | ExplainableBoostingClassifier(learning_rate=0.1, max_bins=64) |
| **Sequential Model (Previous Pregnancy Variables)** |  |
| Random Forest Classifier | RandomForestClassifier(max_depth=20, min_samples_leaf=4, min_samples_split=5, n_estimators=300) |
| Logistic Regression | LogisticRegression(C=0.03359818286283781, max_iter=1000, solver='liblinear') |
| XGBoost Classifier | XGBClassifier(max_depth=3, n_estimators=100, learning_rate=0.1) |
| Explainable Boosting Machine | ExplainableBoostingClassifier(learning_rate=1) |
| **Sequential Model (Including First Trimester Data)** |  |
| Random Forest Classifier | RandomForestClassifier(min_samples_leaf=2, n_estimators=300) |
| Logistic Regression | LogisticRegression(C=0.03359818286283781, max_iter=1000, solver='liblinear') |
| XGBoost Classifier | XGBClassifier(max_depth=10, n_estimators=100, learning_rate=0.1) |
| Explainable Boosting Machine | ExplainableBoostingClassifier() |
| **Sequential Model (Top 8 Features)** |  |
| Random Forest Classifier | RandomForestClassifier(max_depth=10, min_samples_split=5) |
| Logistic Regression | LogisticRegression(C=0.08858667904100823, max_iter=1000) |
| XGBoost Classifier | XGBClassifier(max_depth=5, n_estimators=300, learning_rate=0.01) |
| Explainable Boosting Machine | ExplainableBoostingClassifier() |

**S Table 6.**
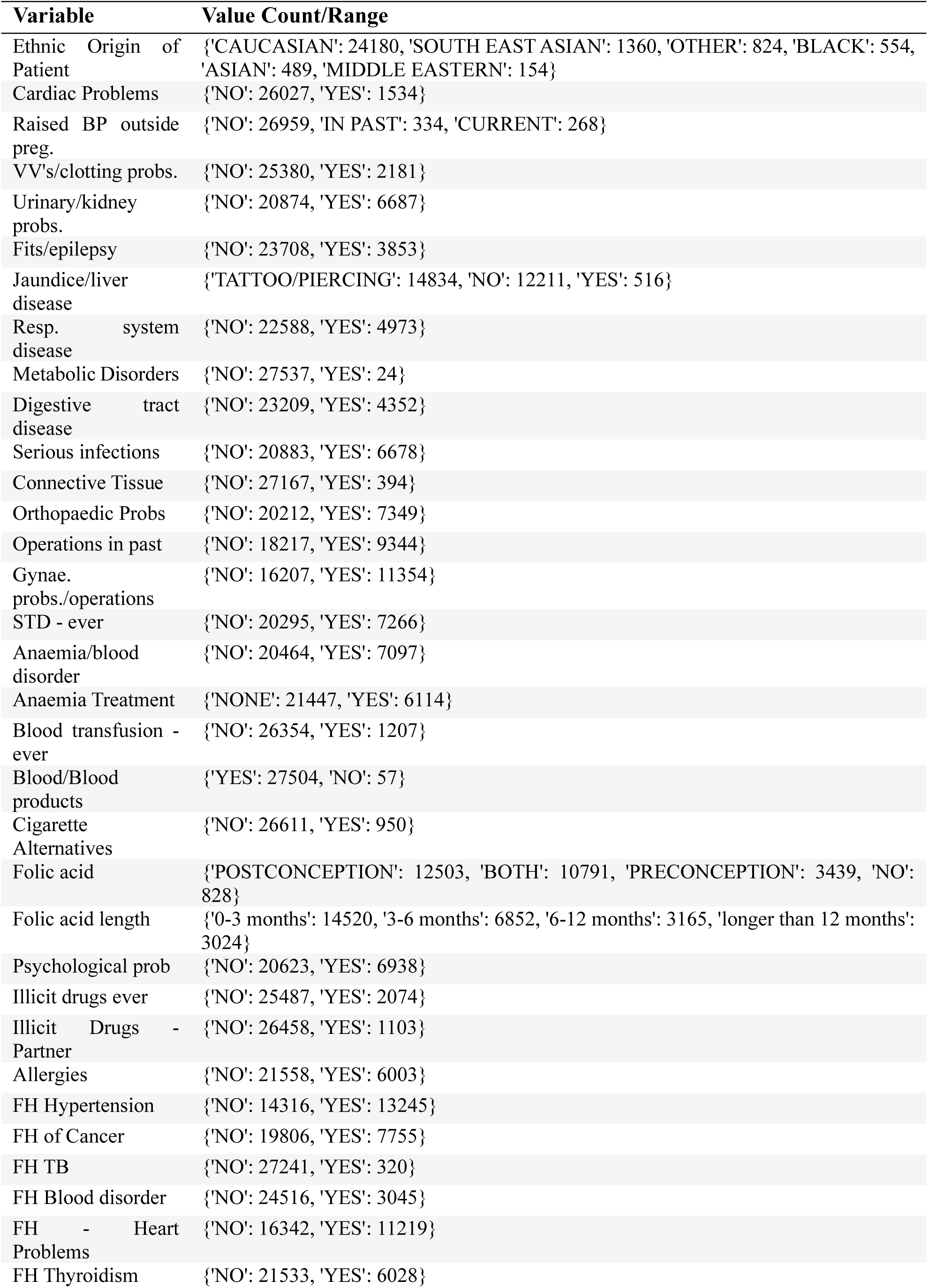
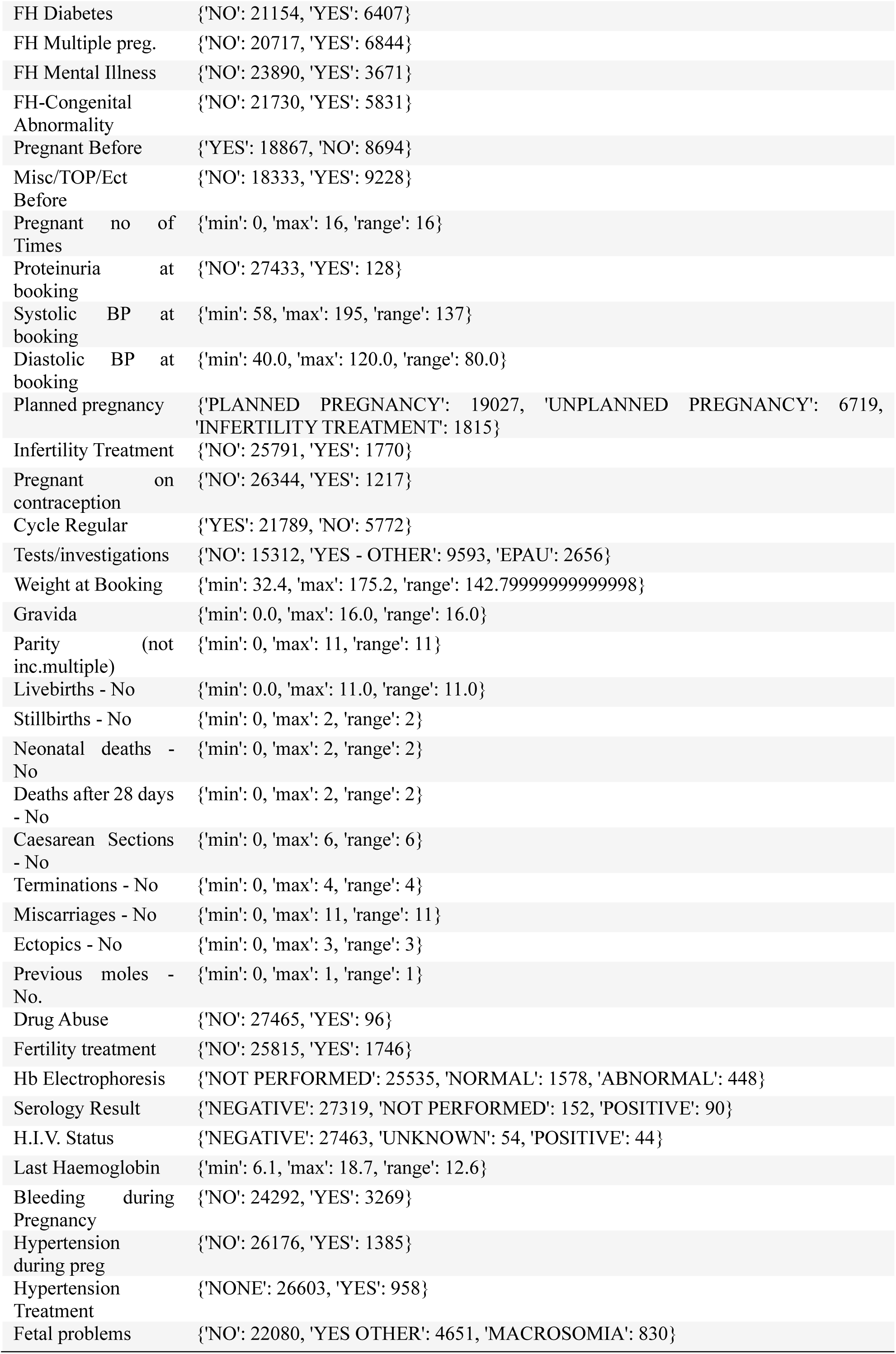
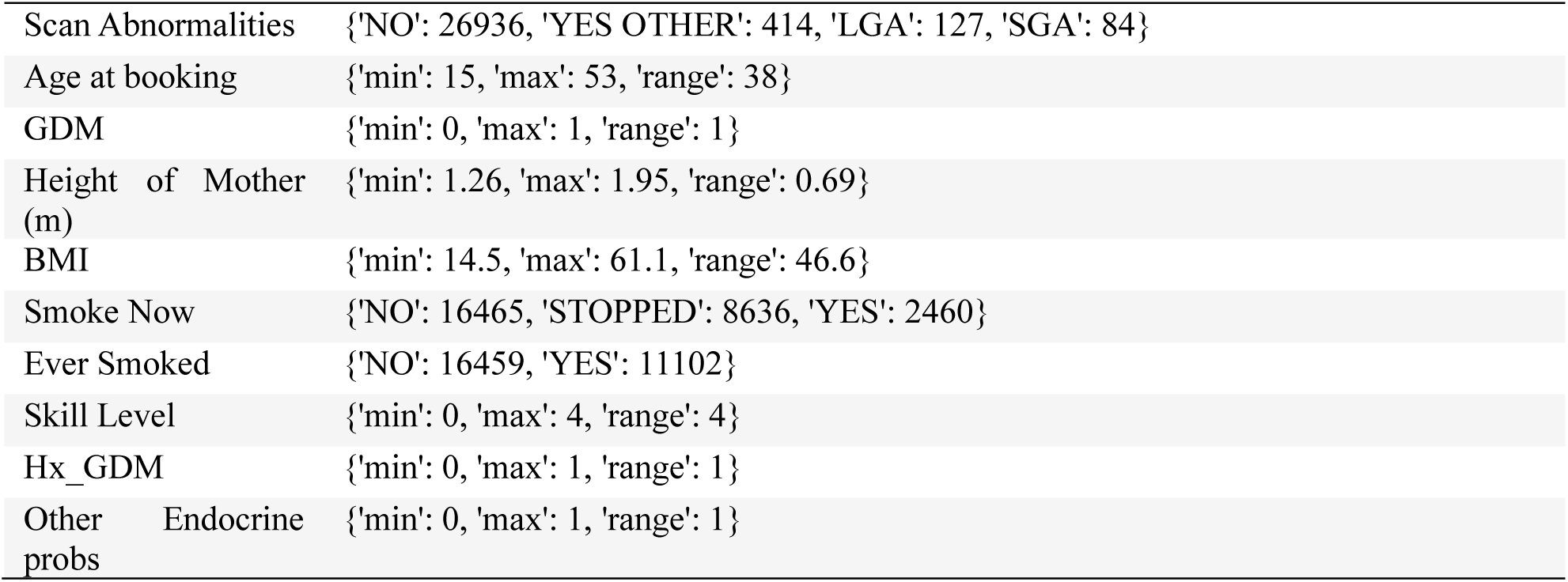
List, value count and ranges of all data available in the electronic health record during the first trimester.

