## Supplementary figures and images for "Evaluation of Machine Learning Models for Early Prediction of Gestational Diabetes Using Retrospective Electronic Health Records from Current and Previous Pregnancies"

### Supplementary Figure 1.

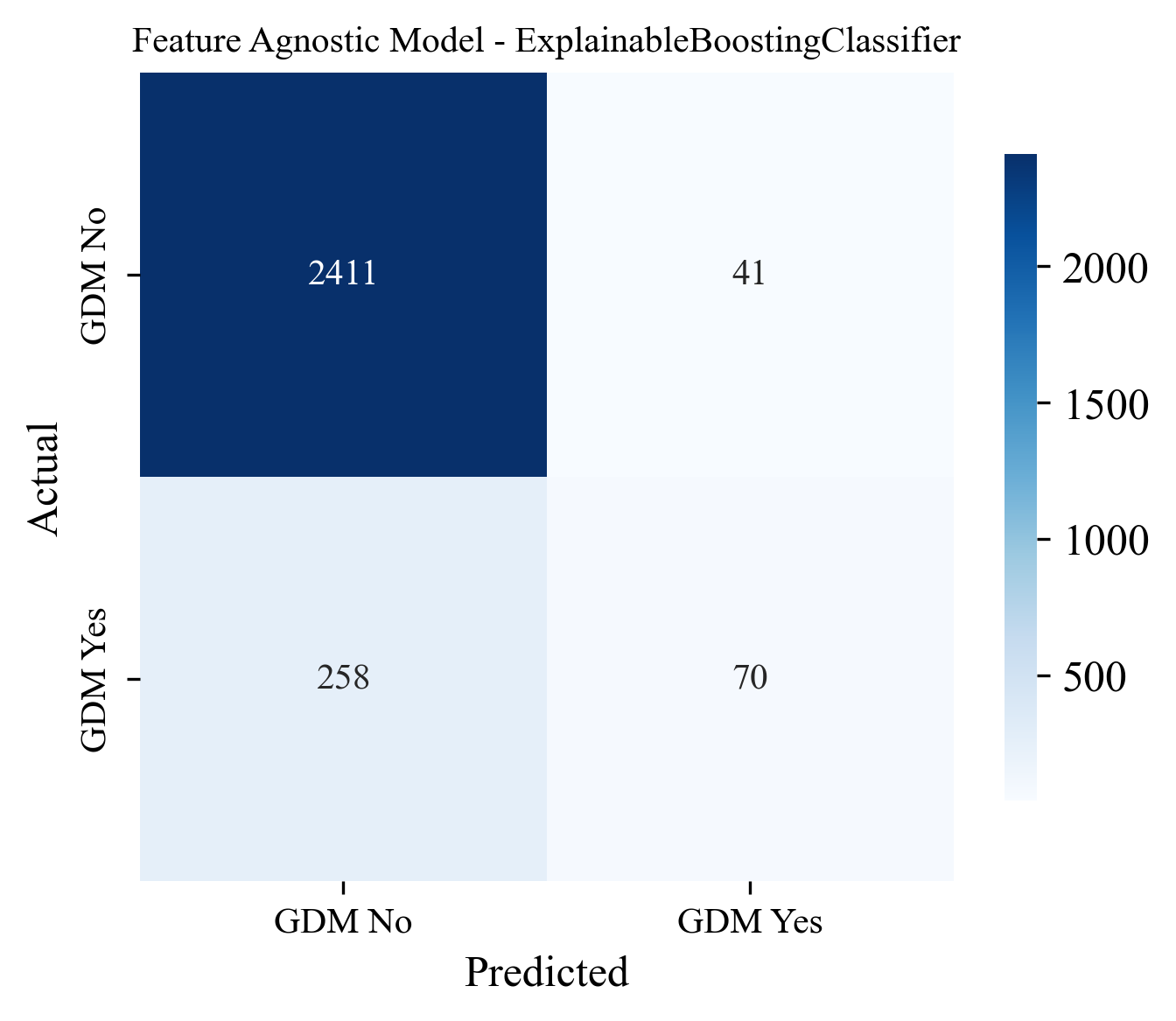

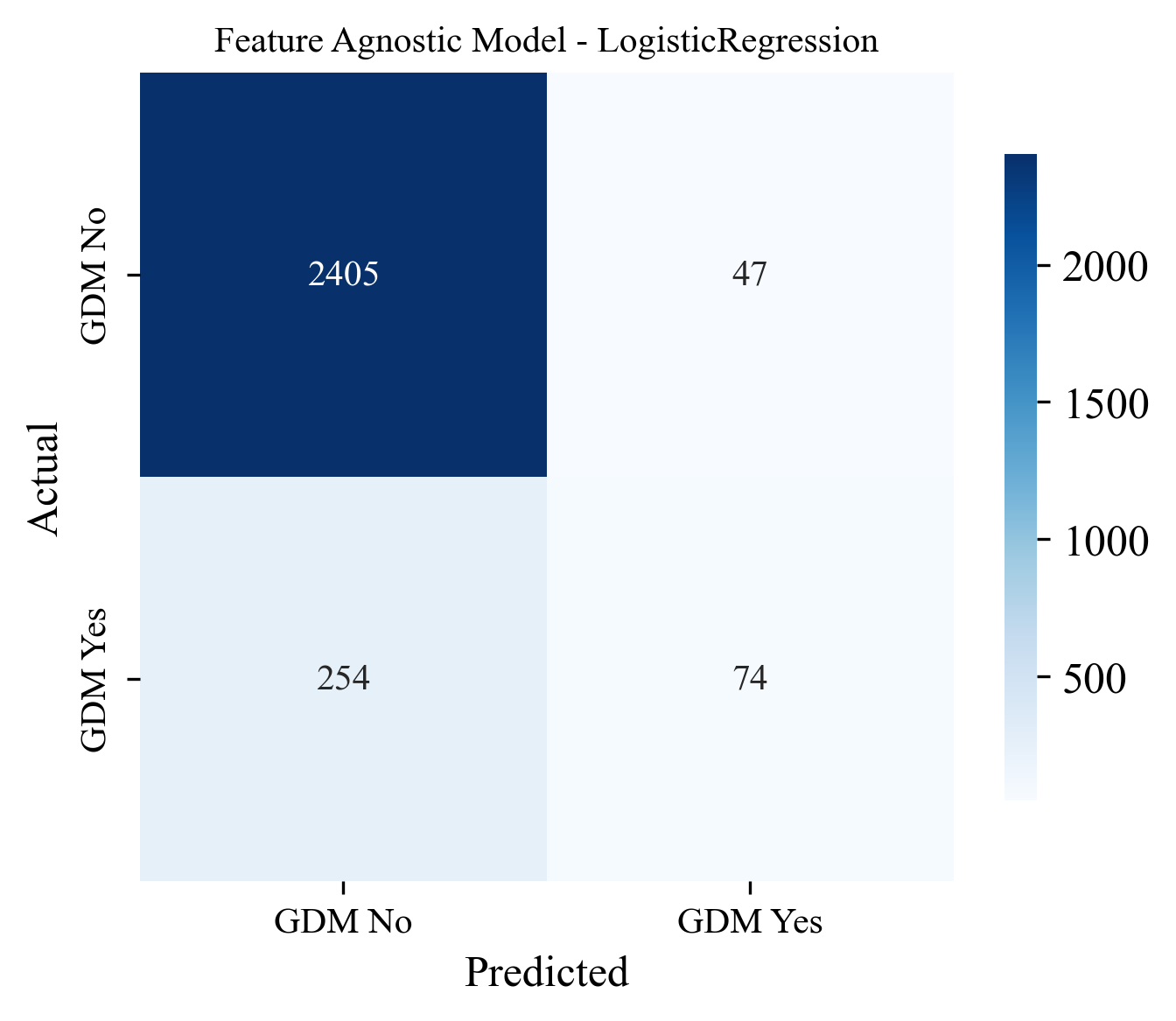

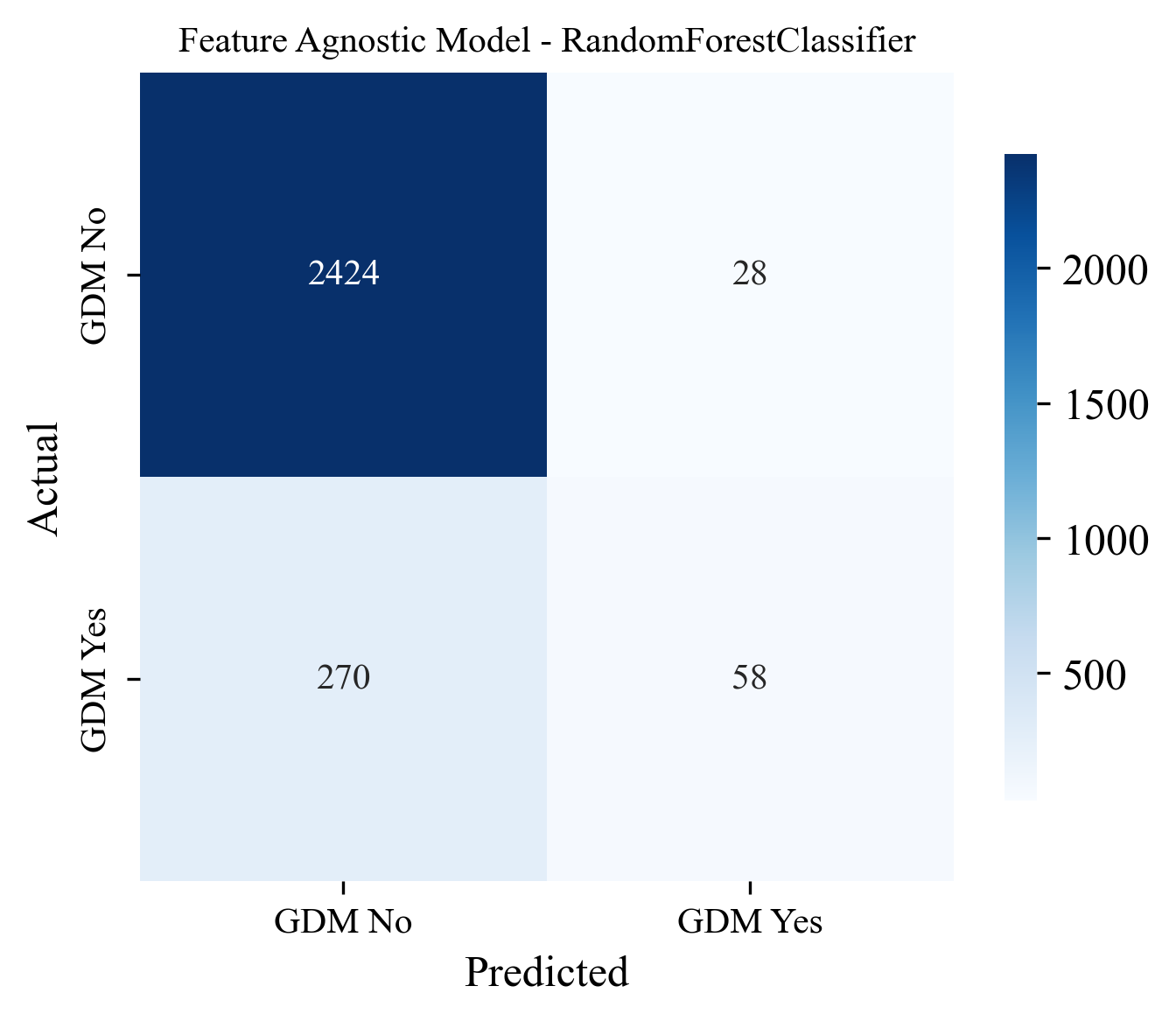

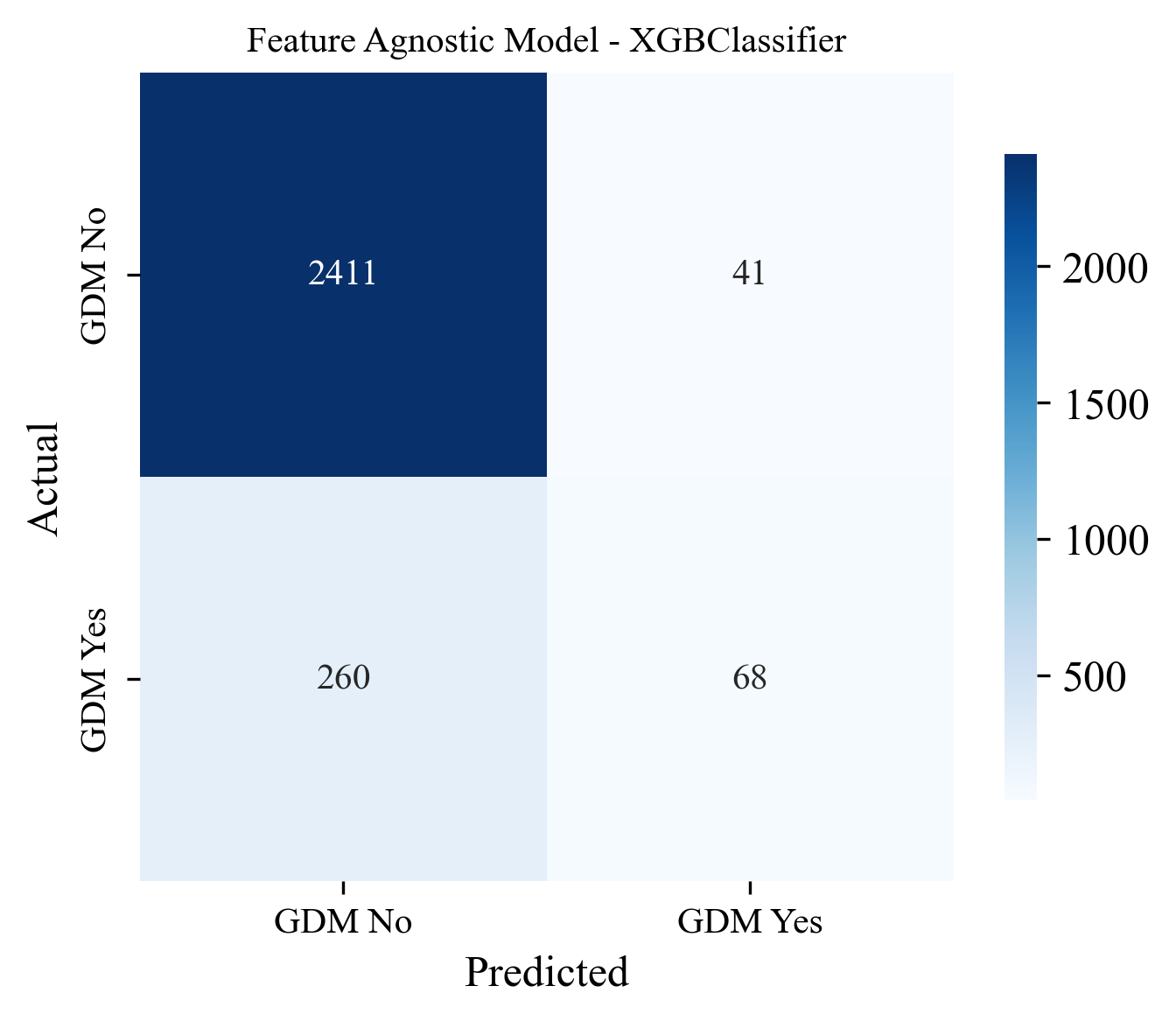

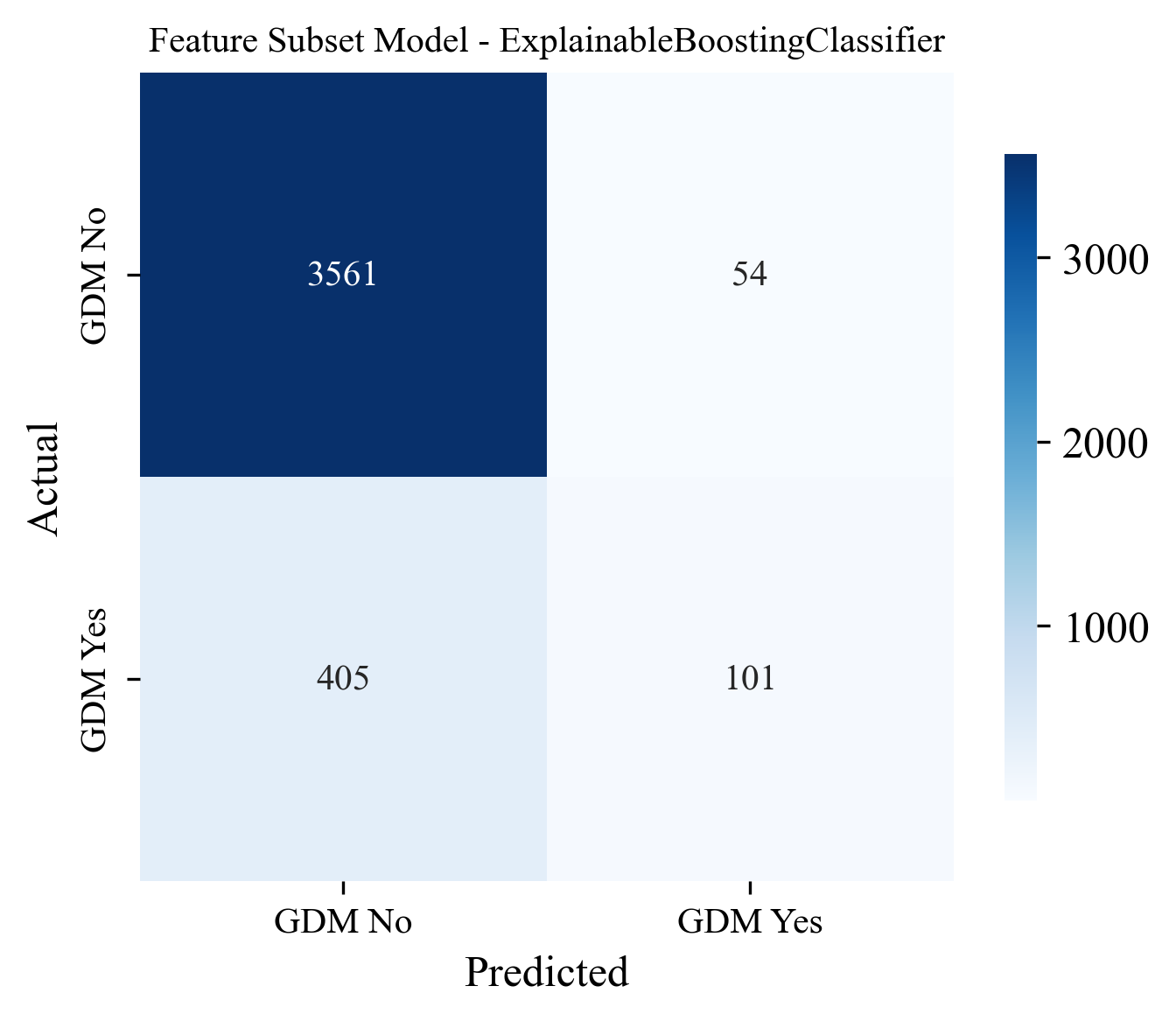

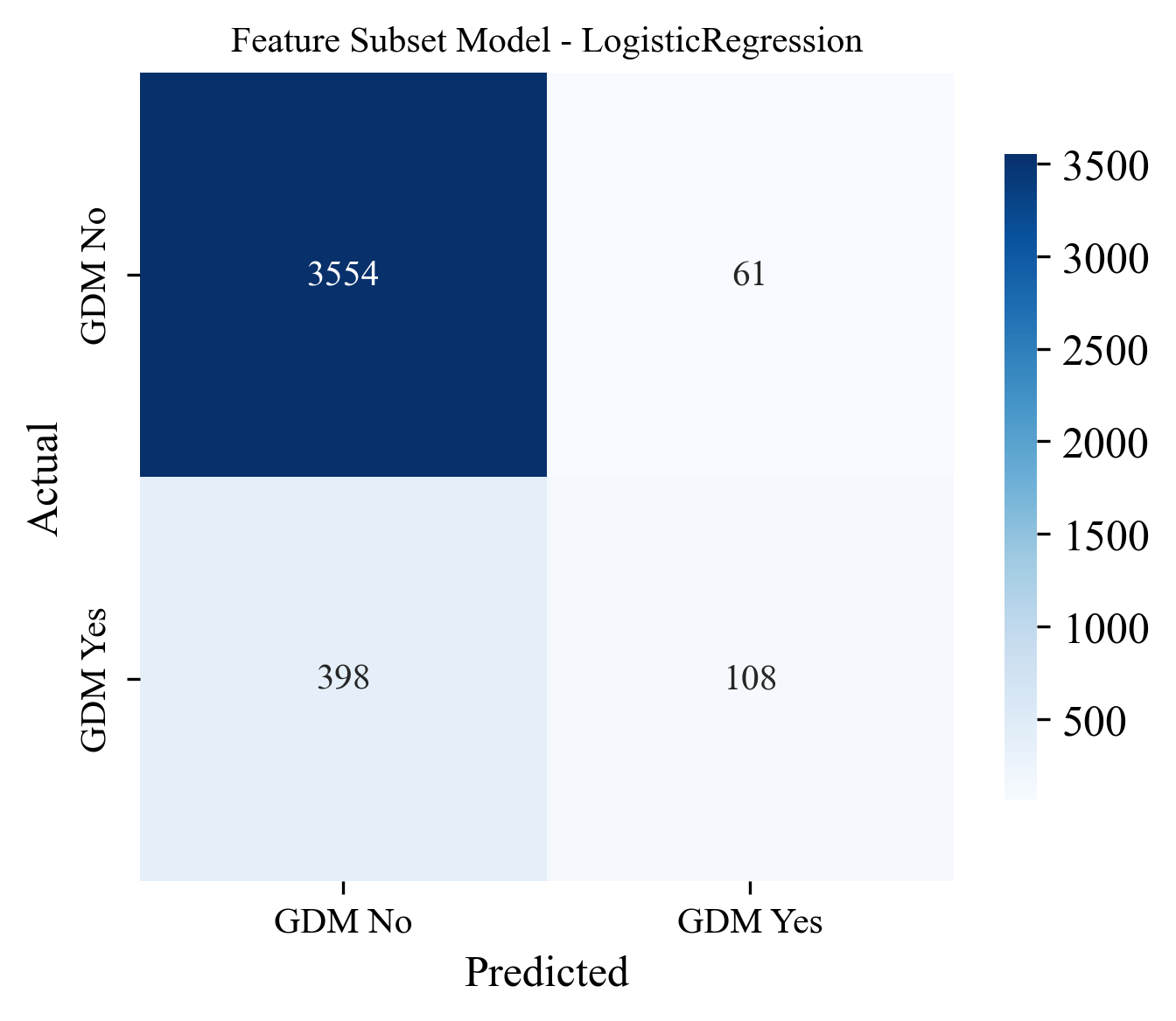

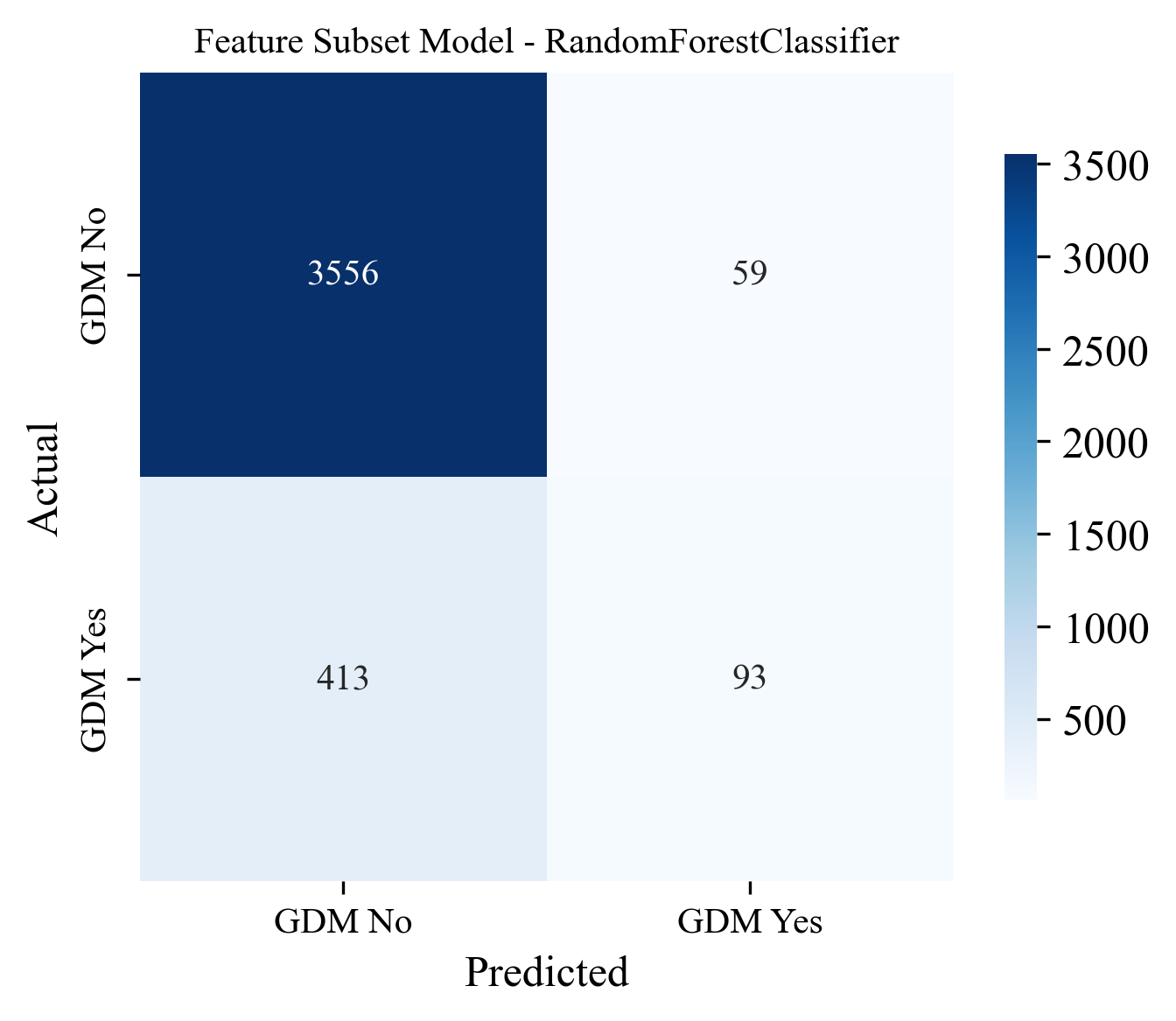

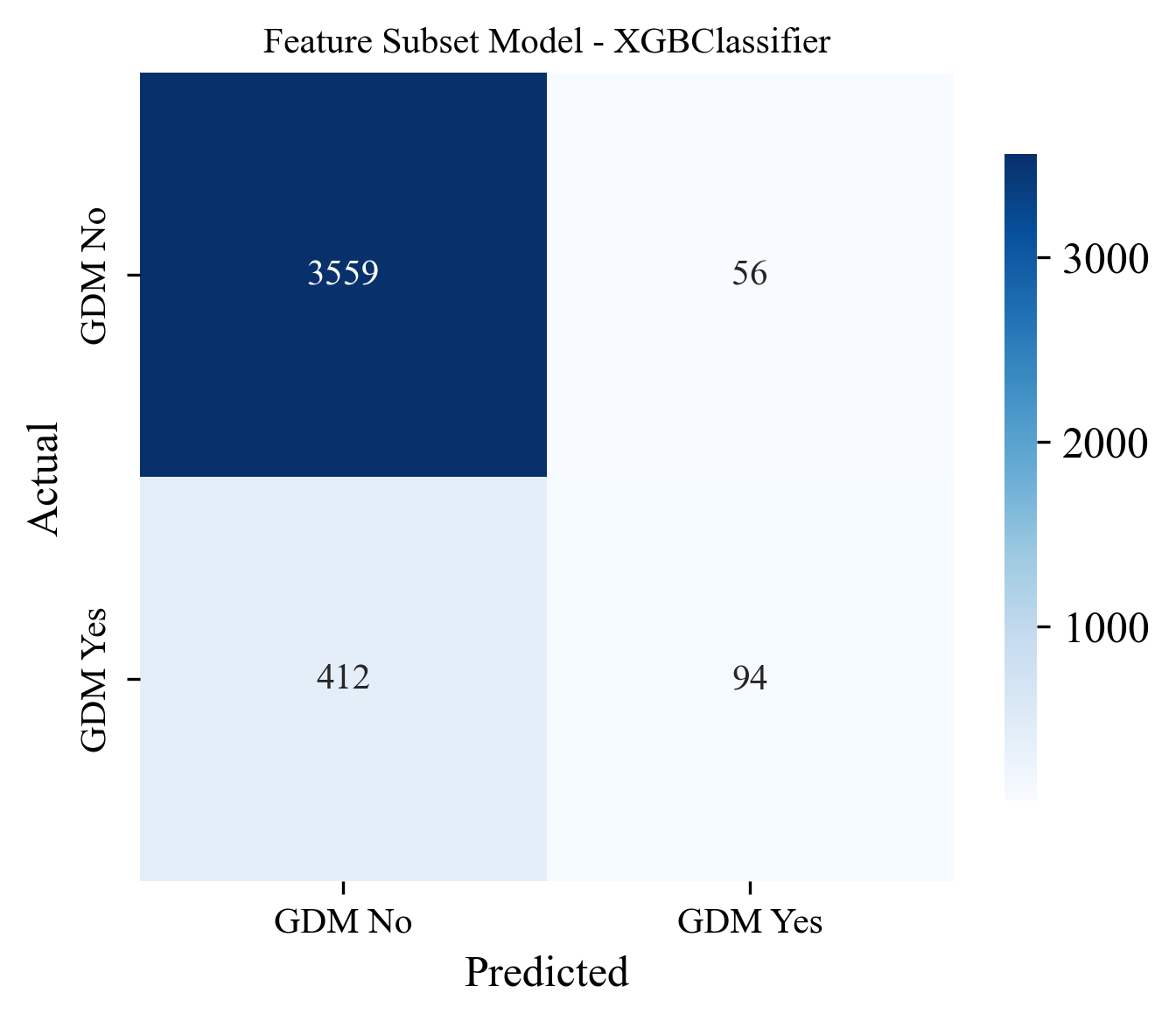

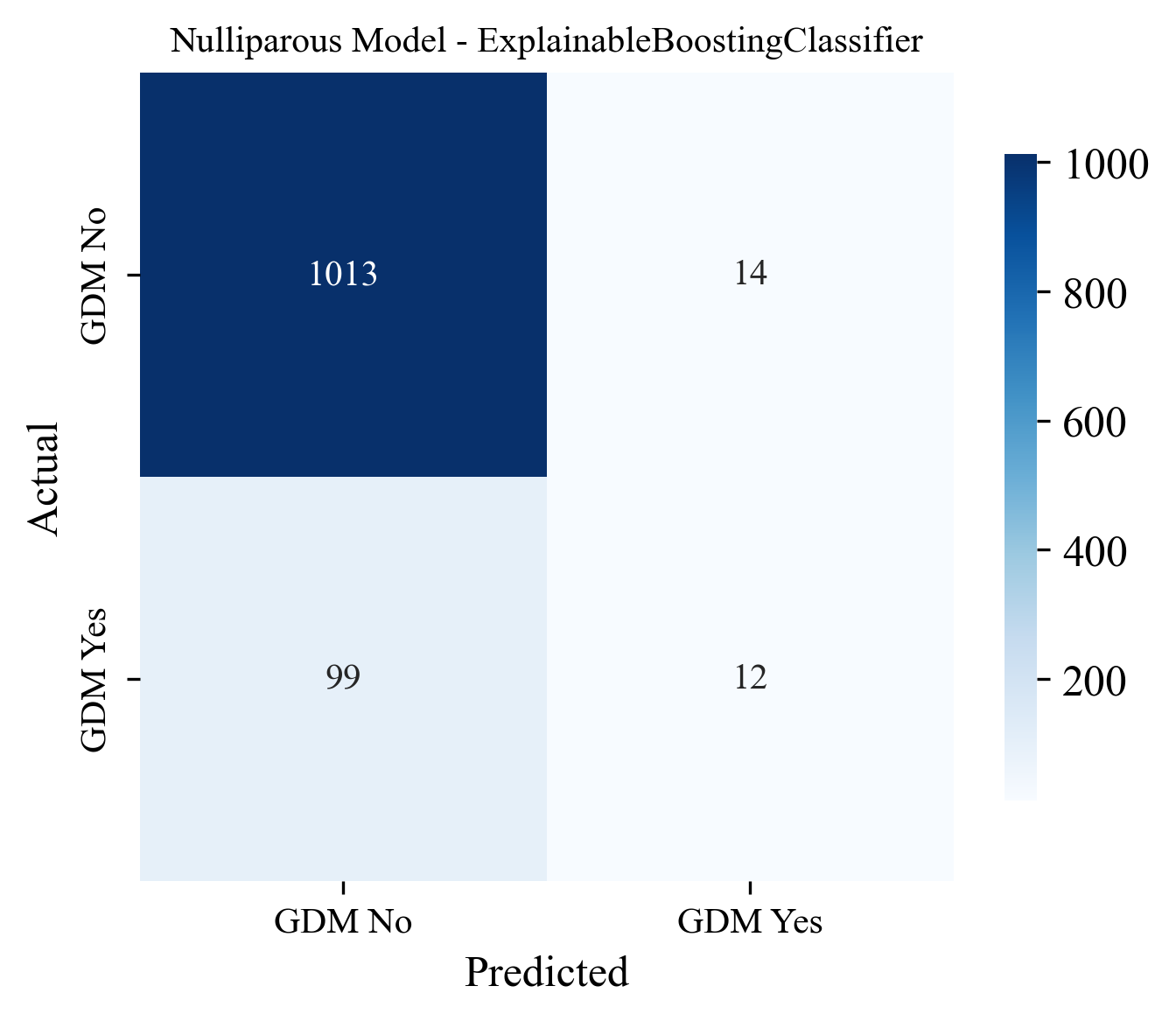

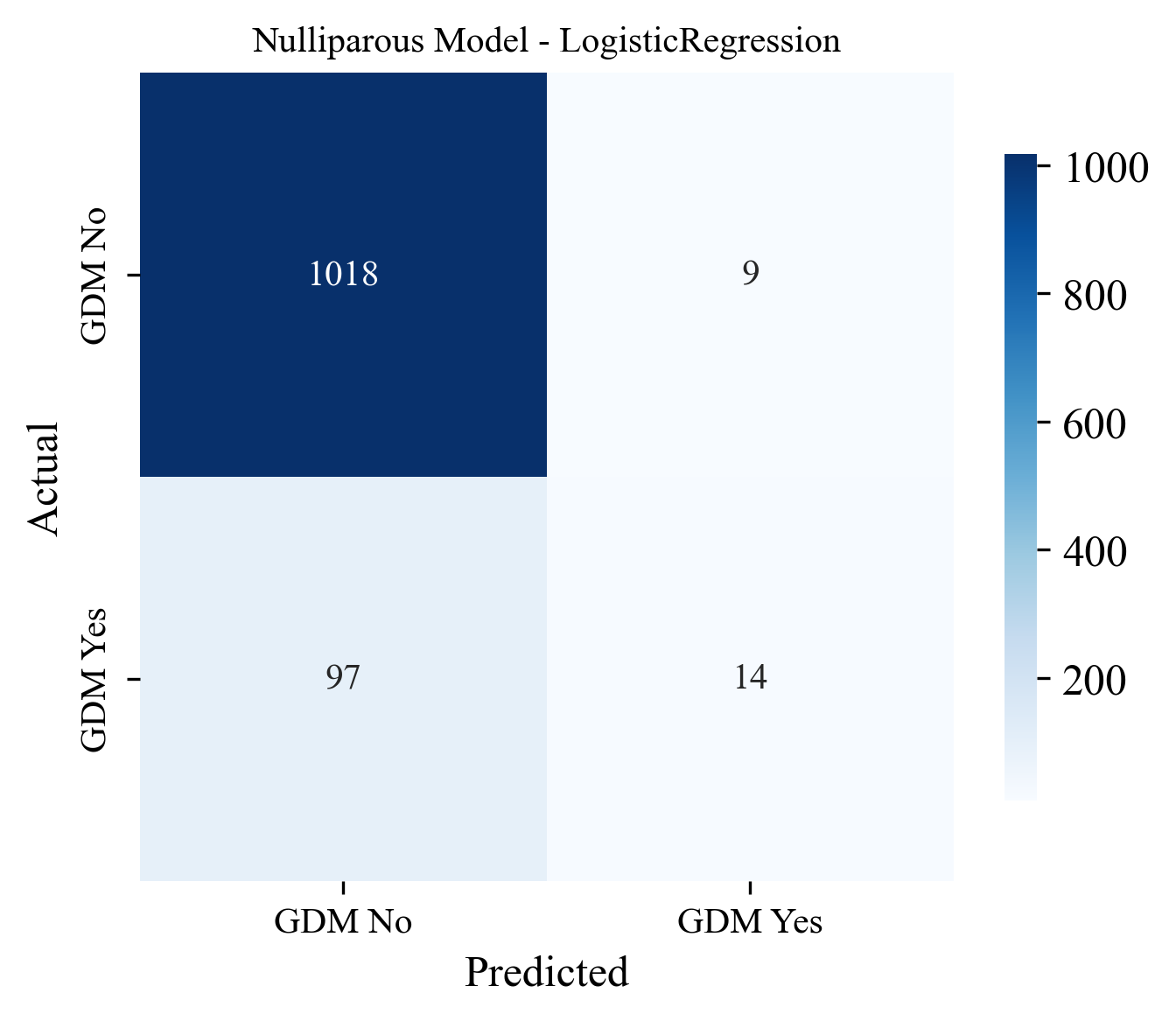

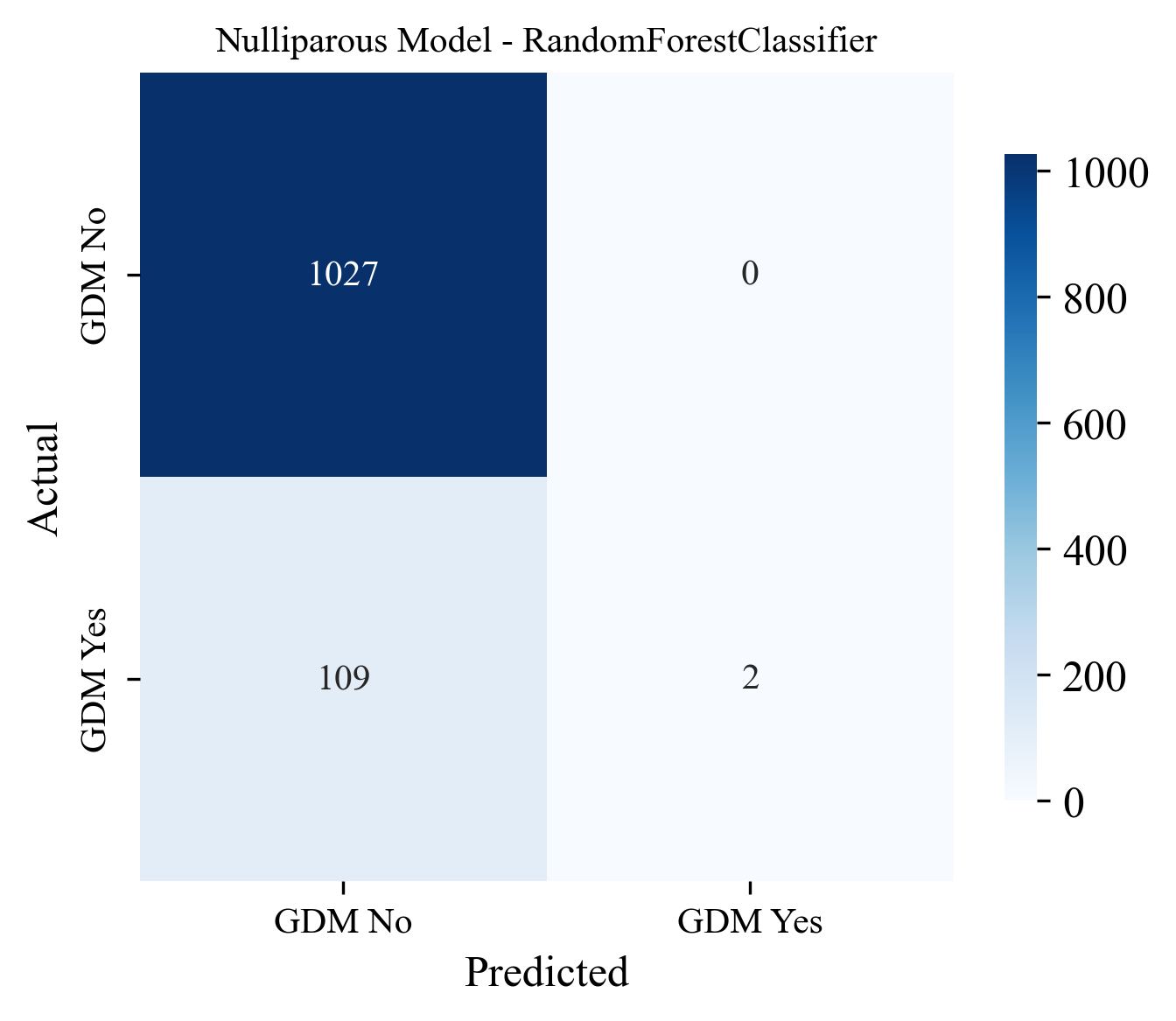

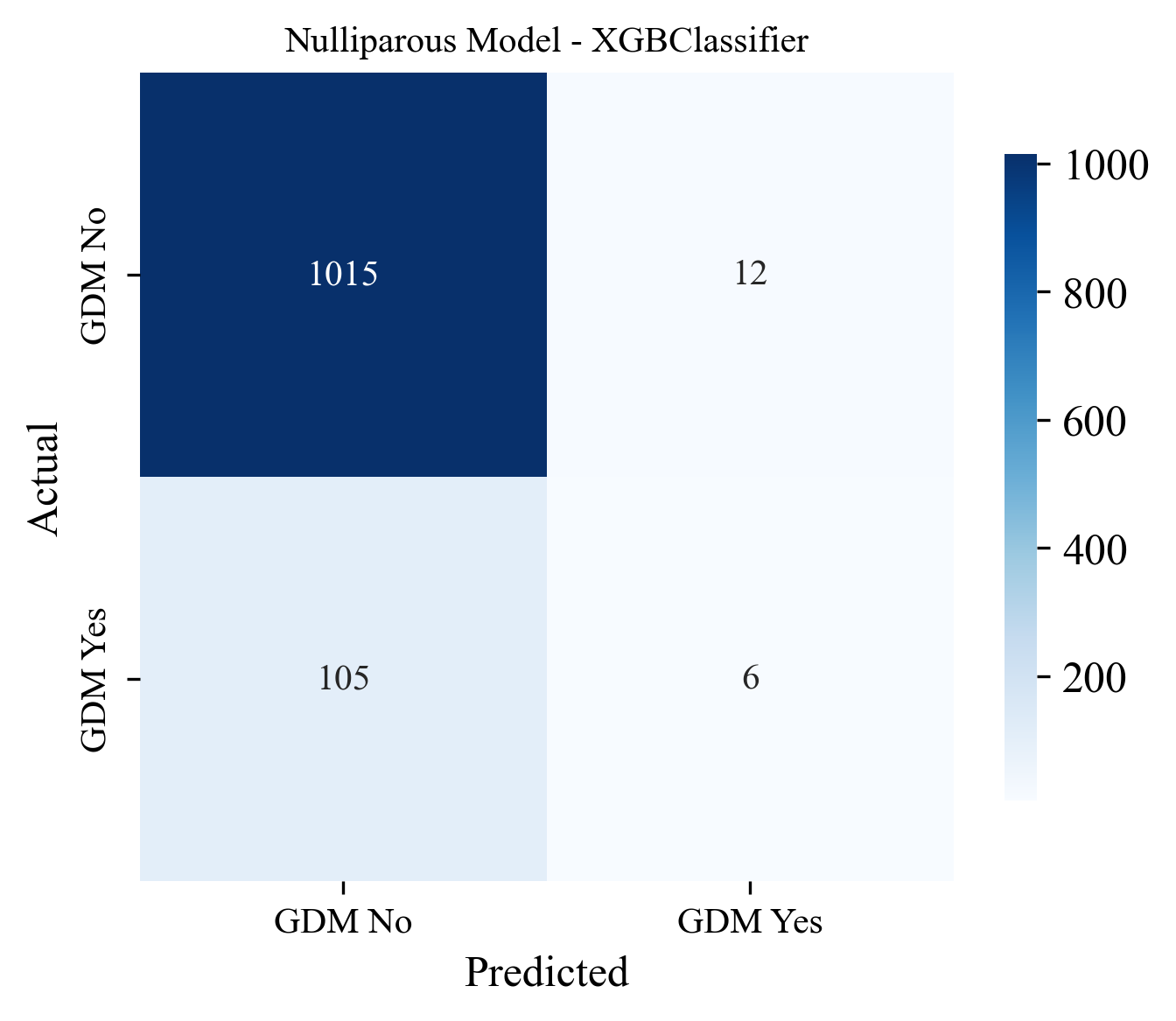

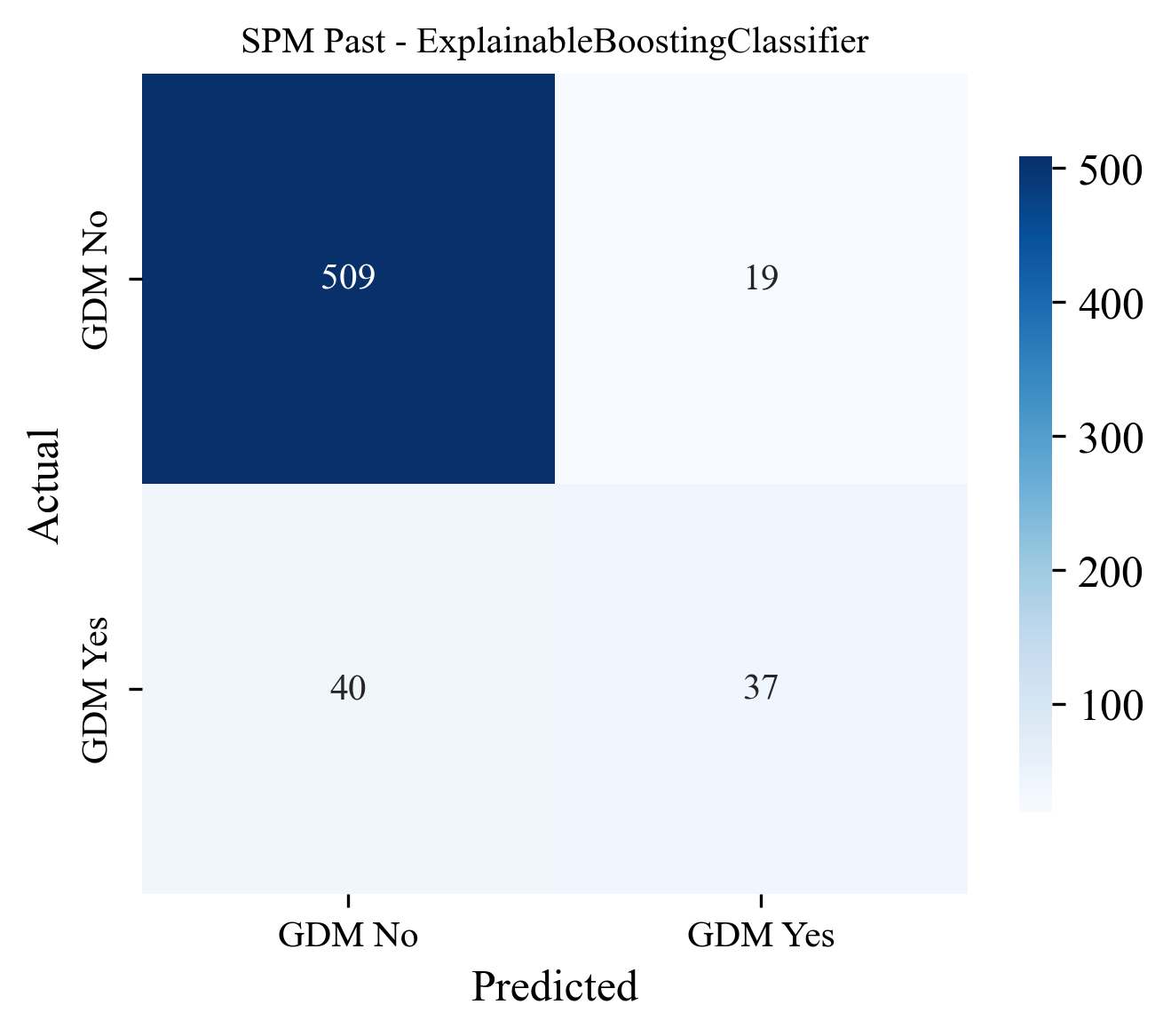

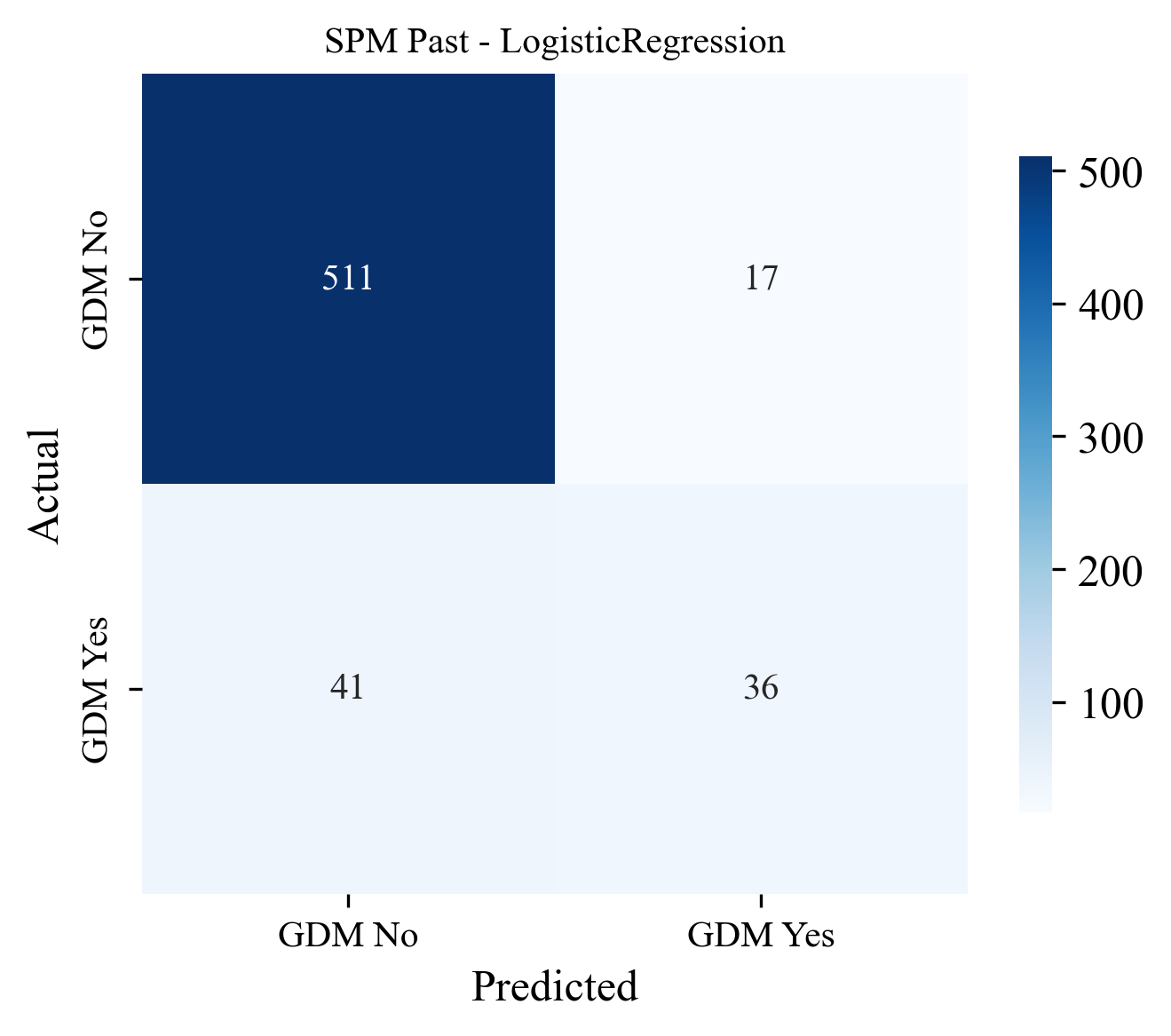

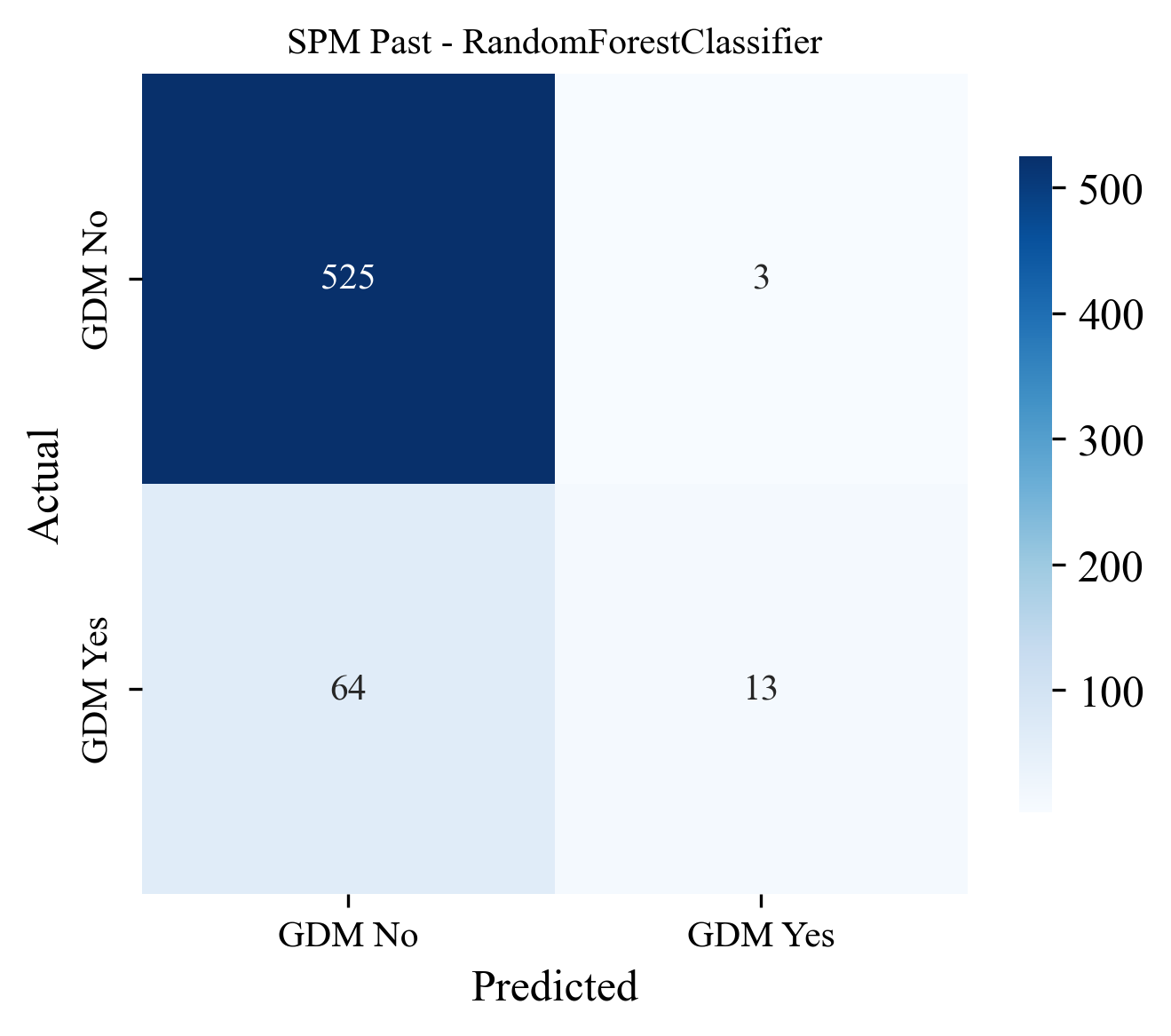

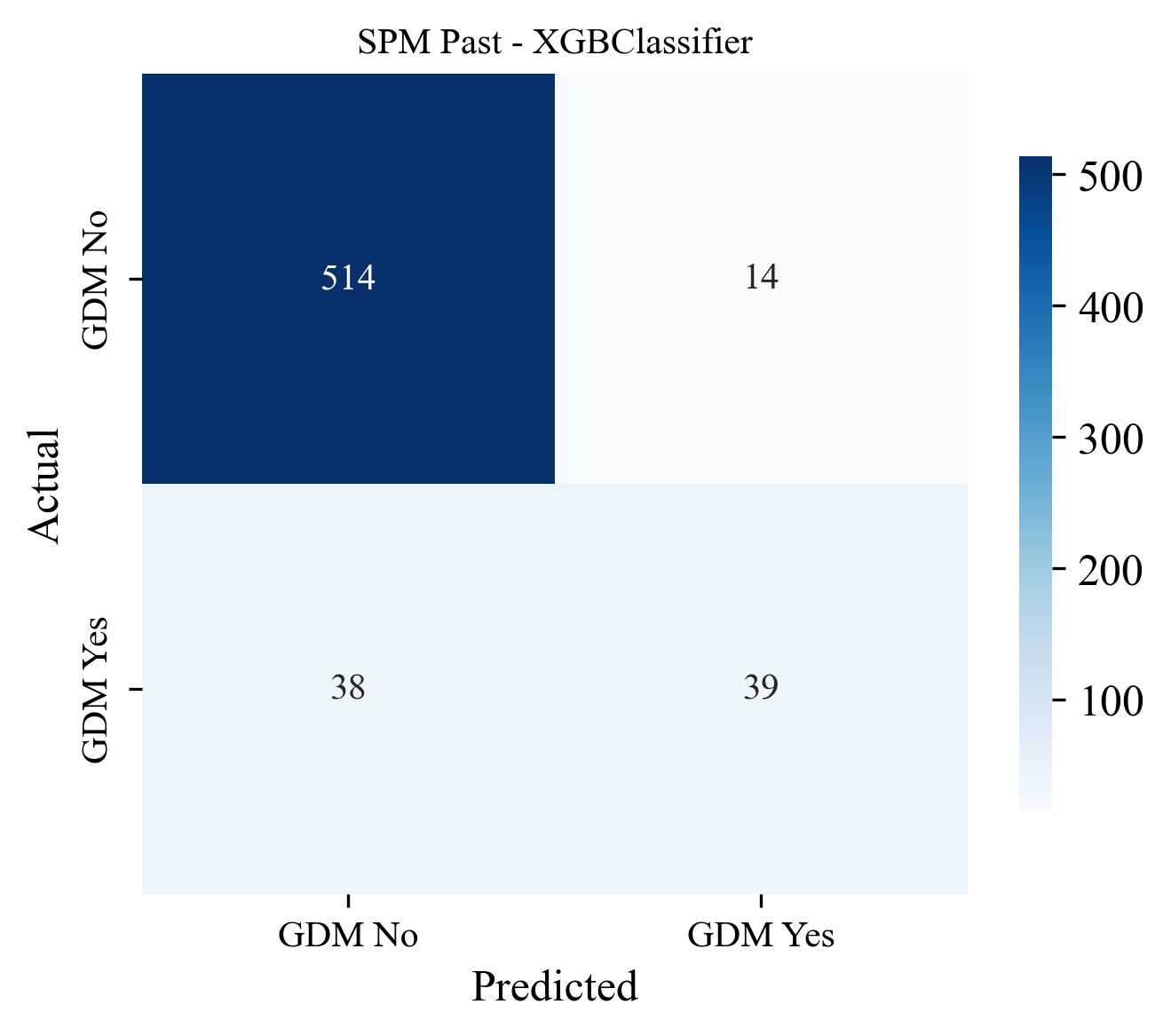

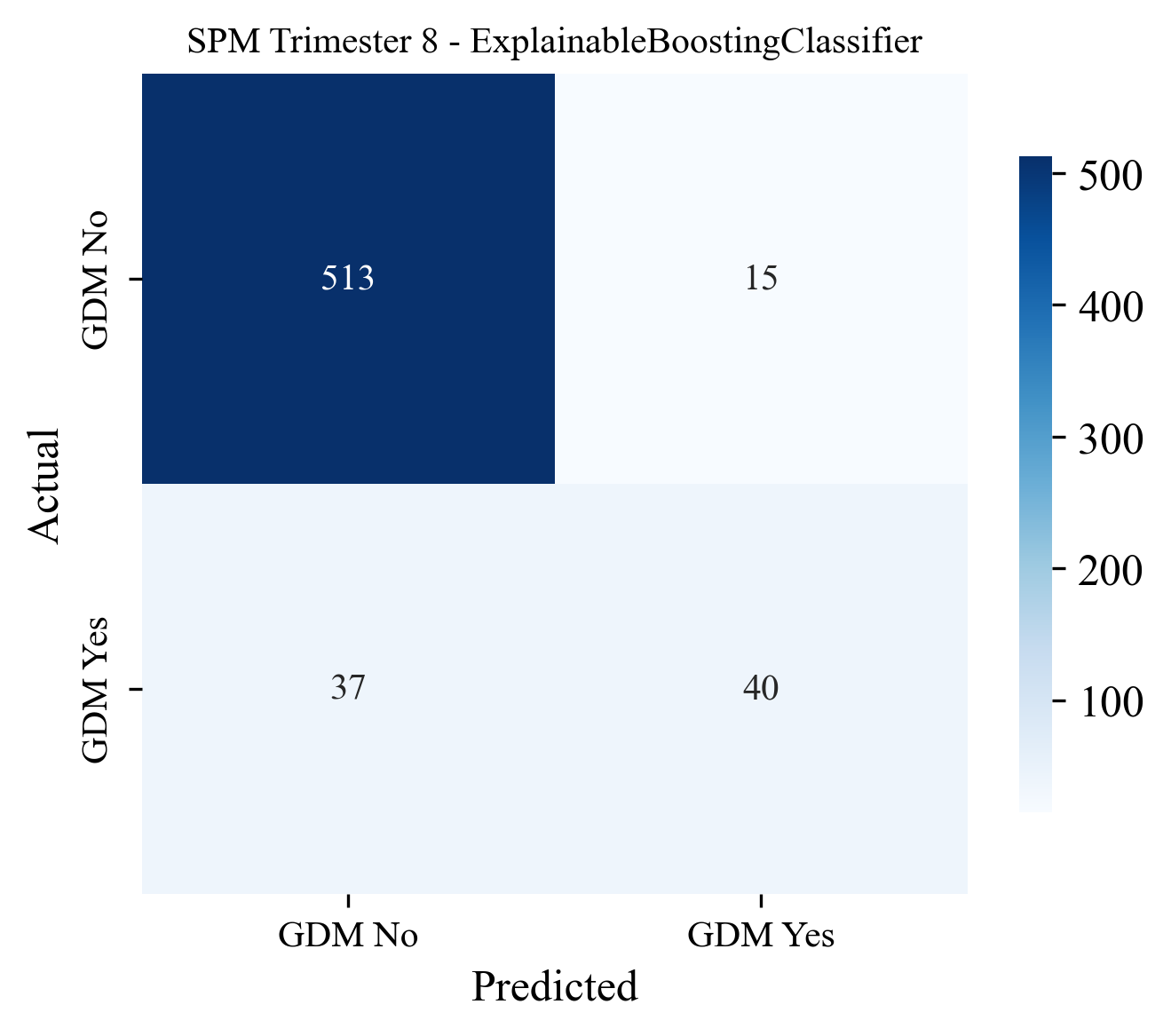

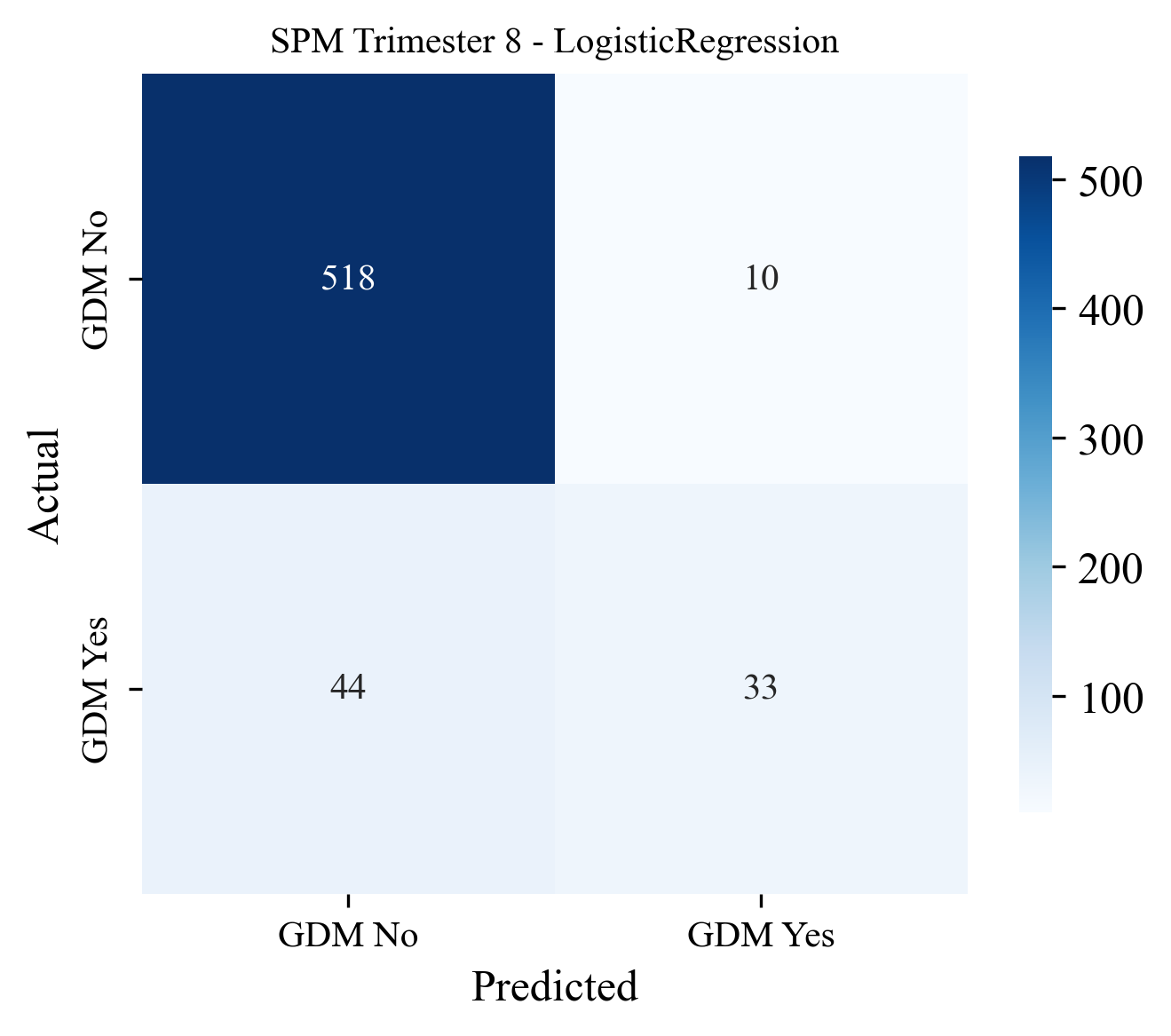

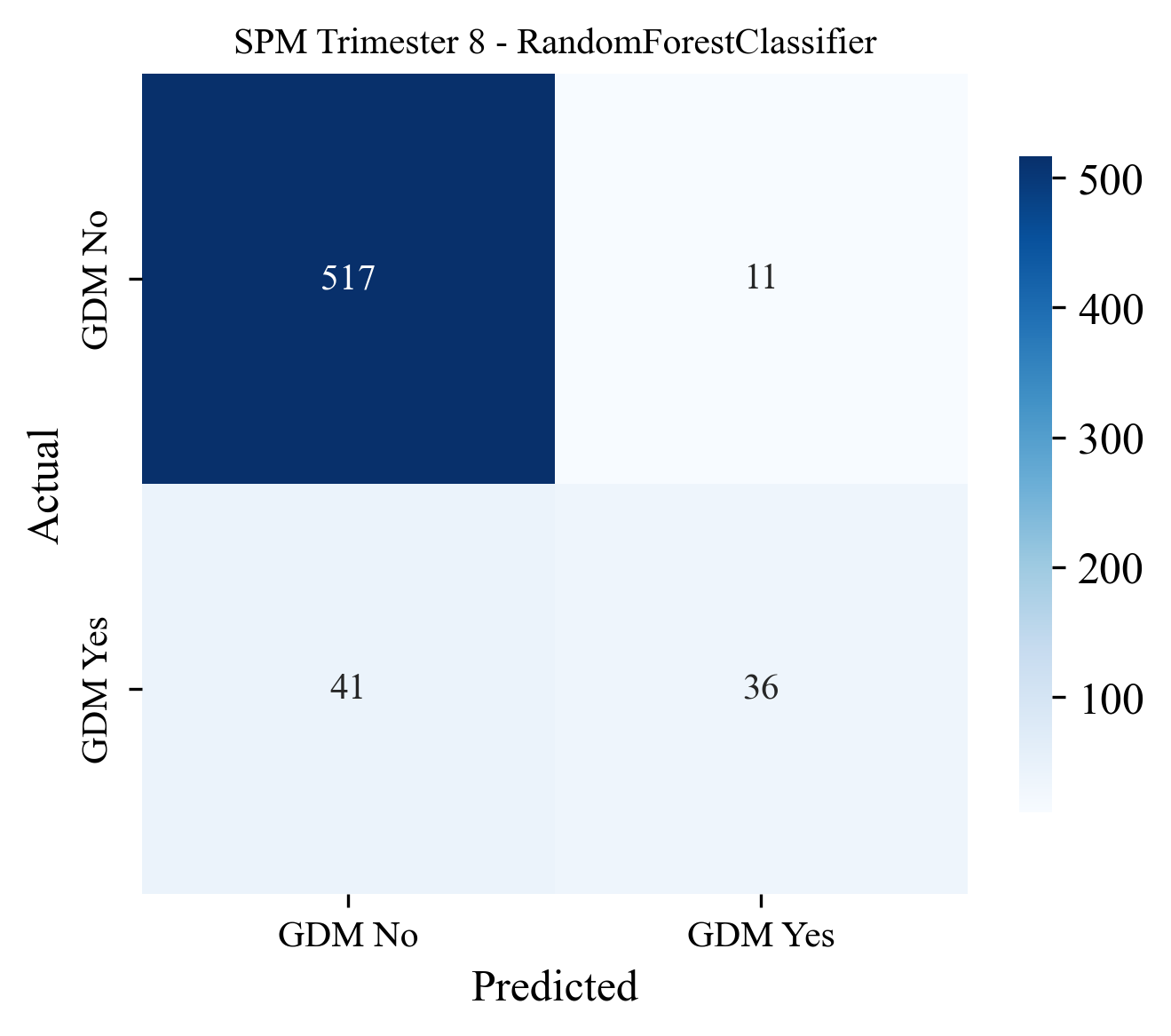

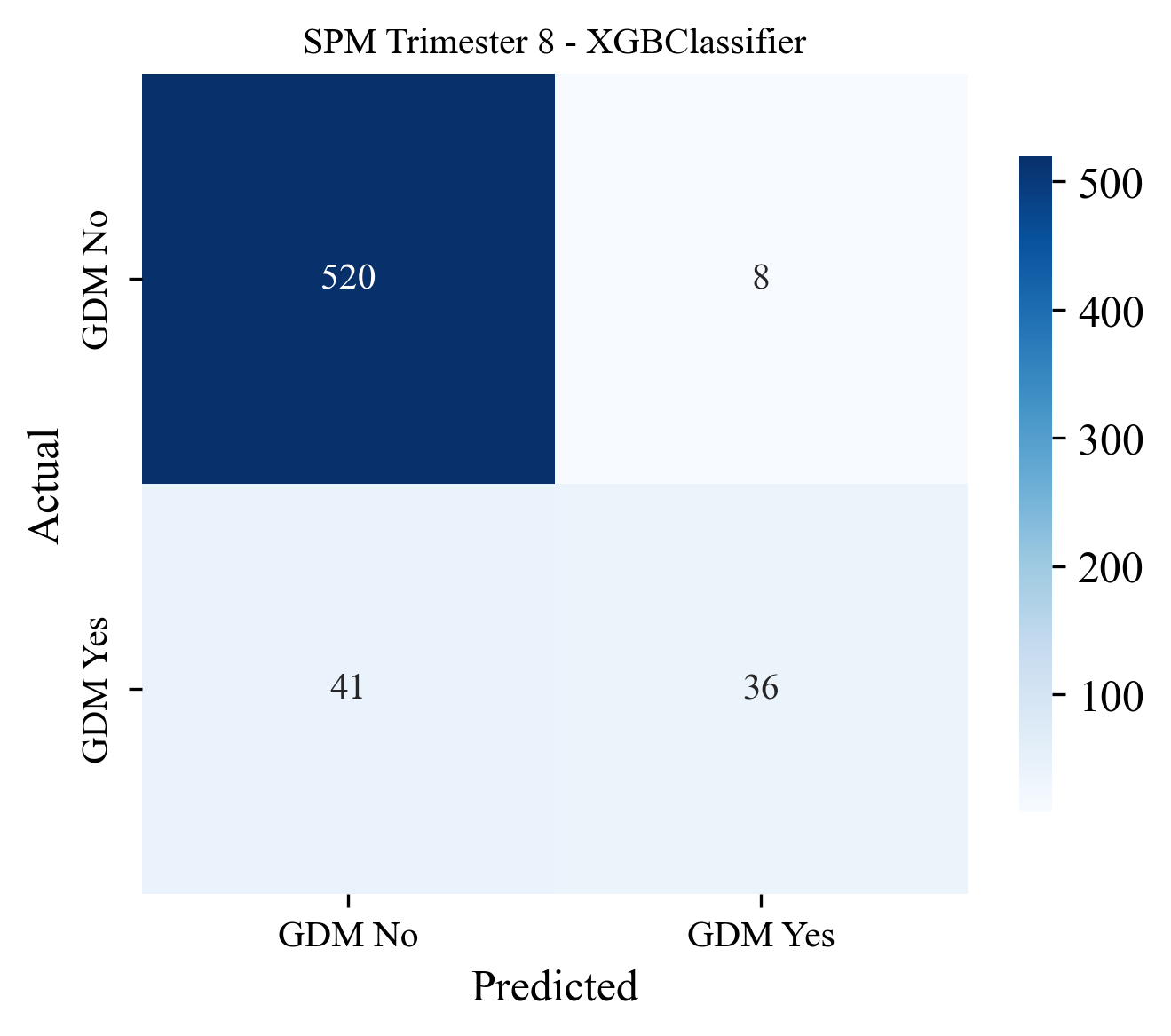

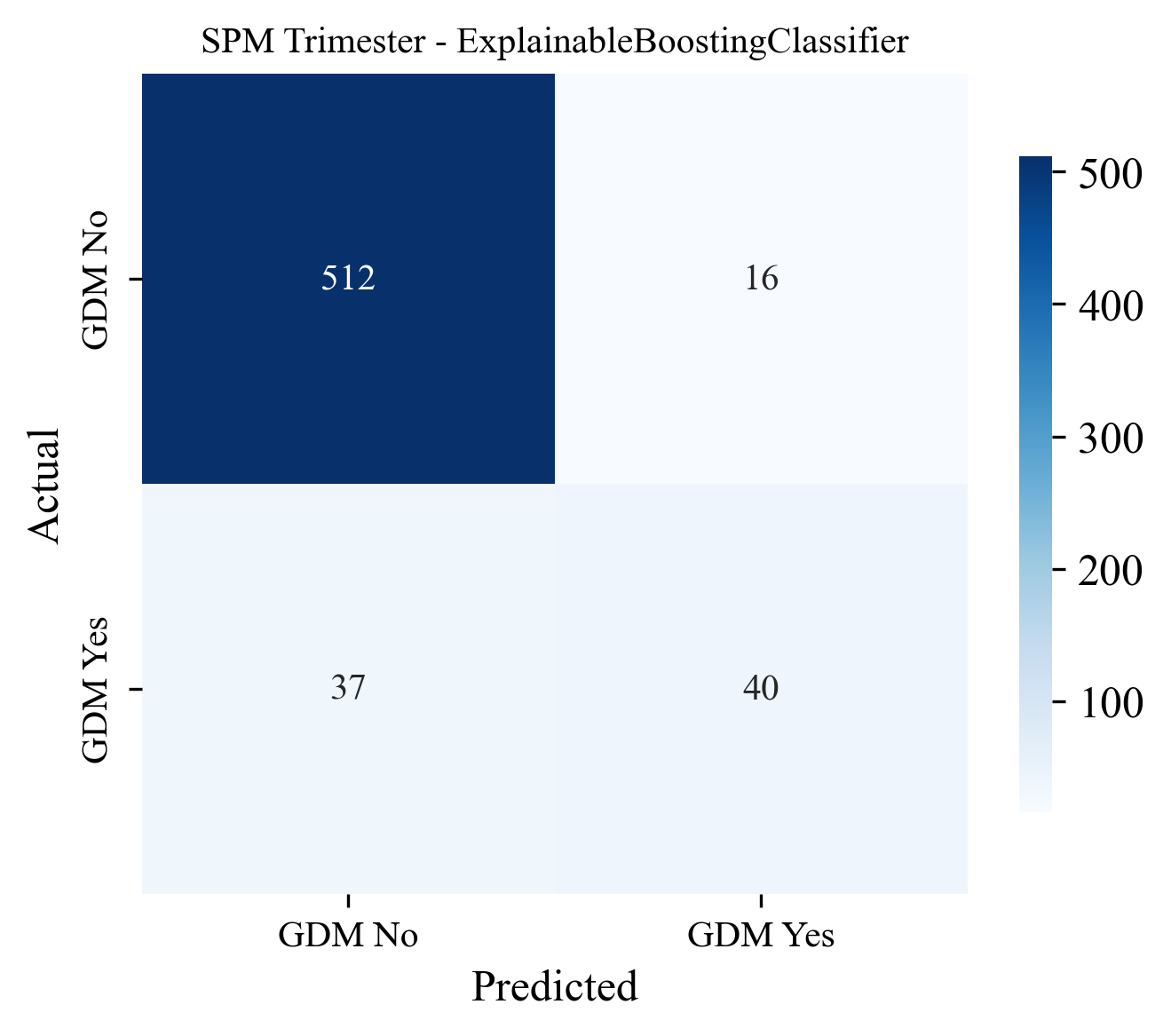

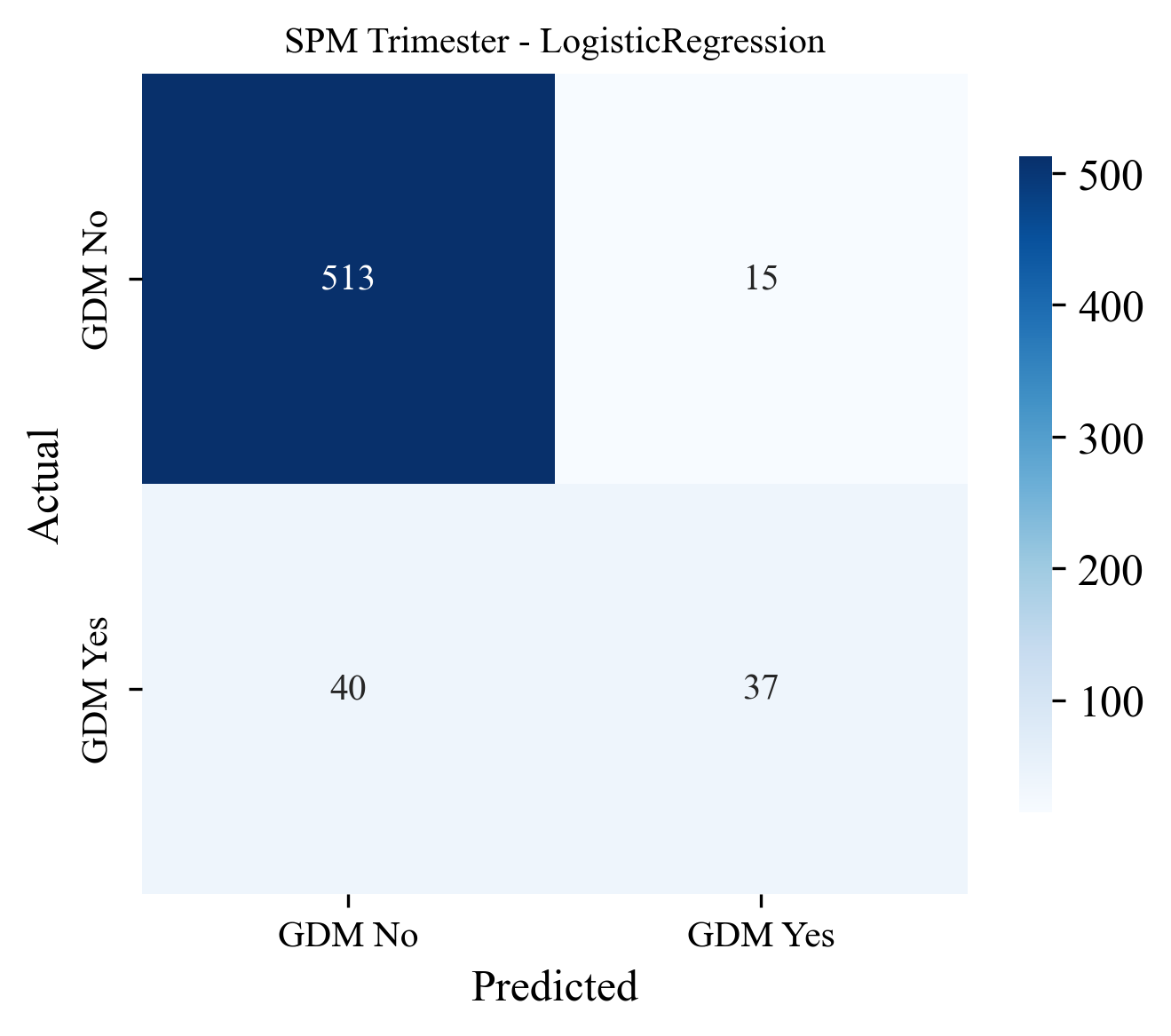

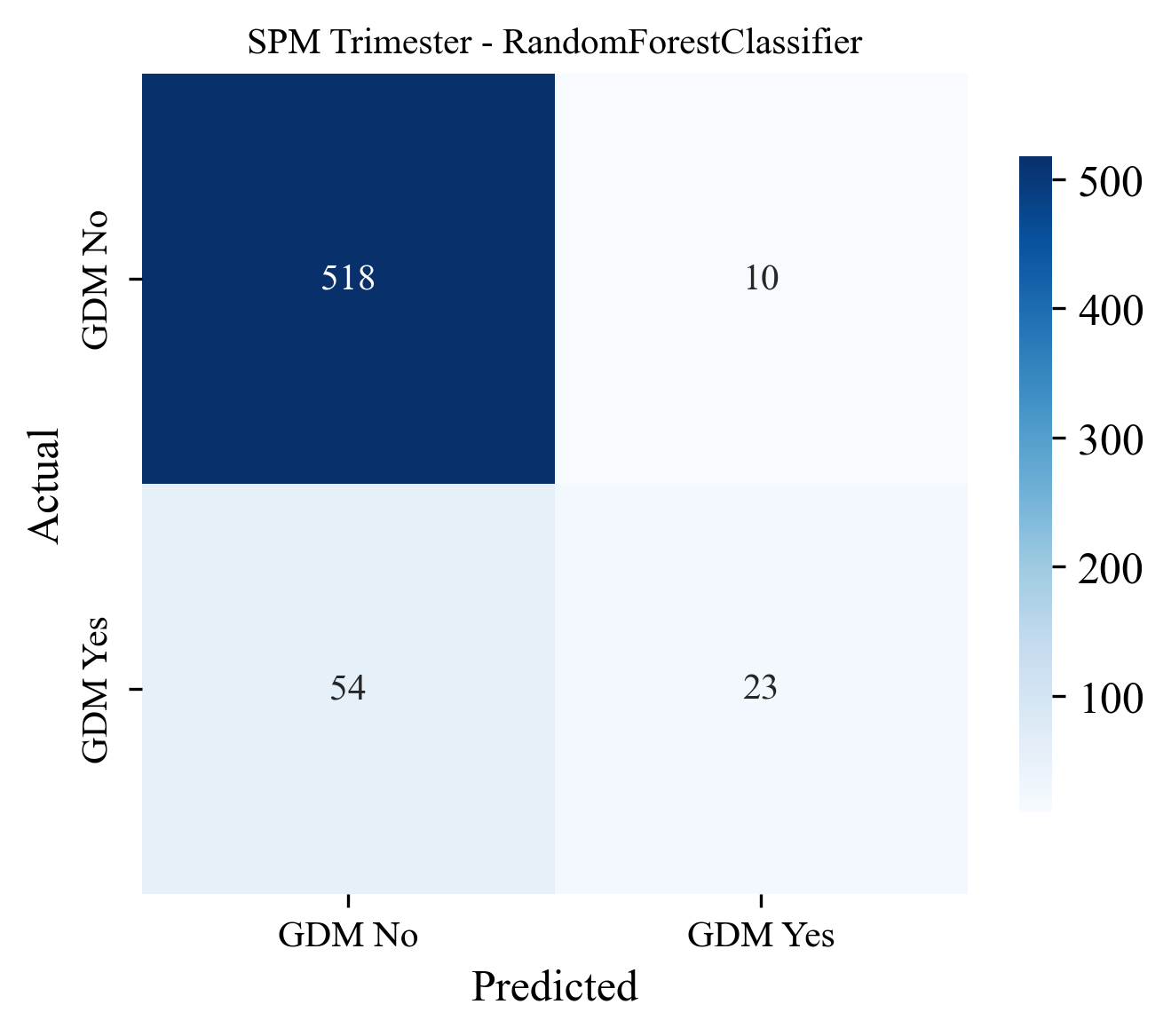

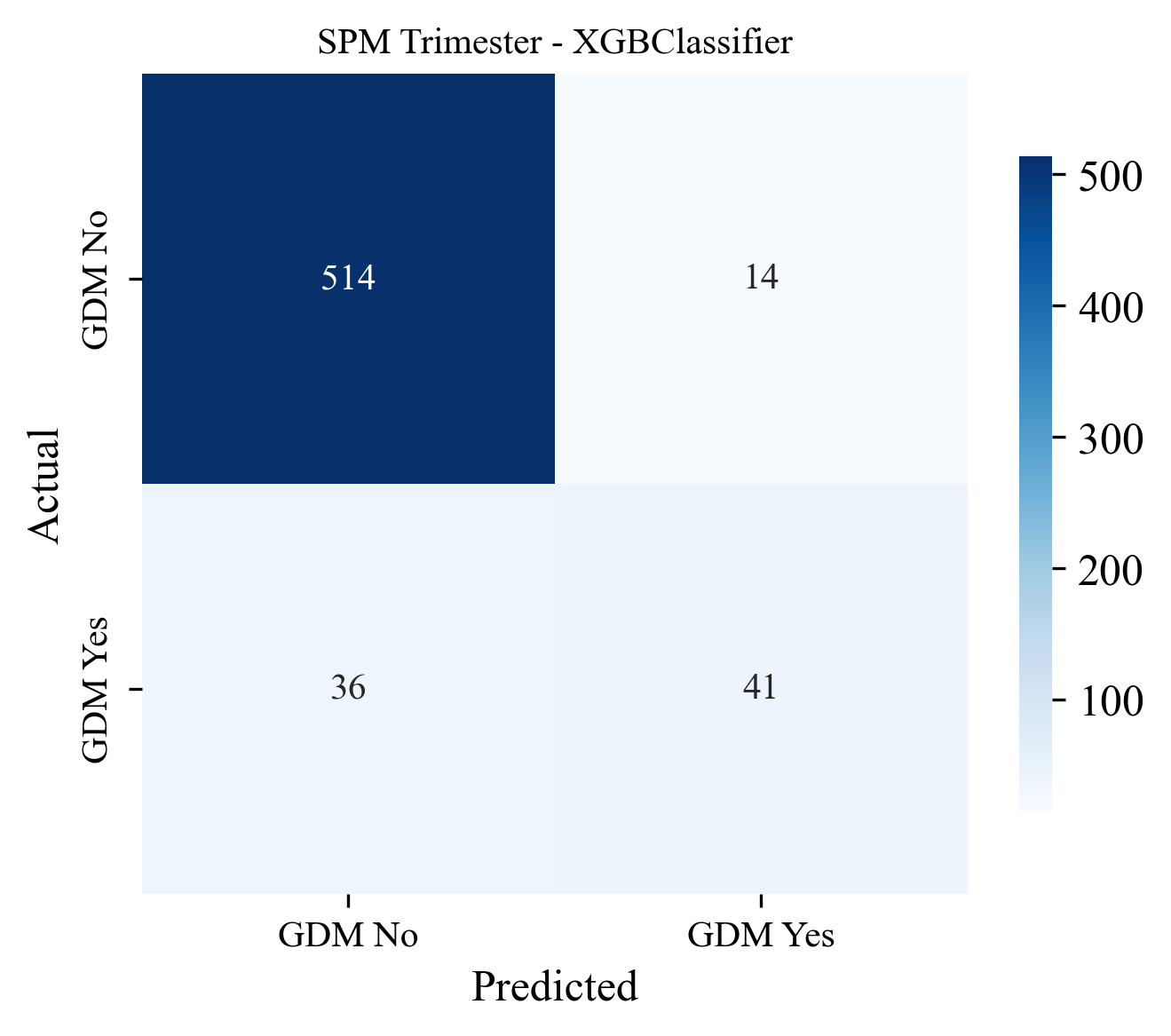
