## Supplementary Figure 2. for "Evaluation of Machine Learning Models for Early Prediction of Gestational Diabetes Using Retrospective Electronic Health Records from Current and Previous Pregnancies"

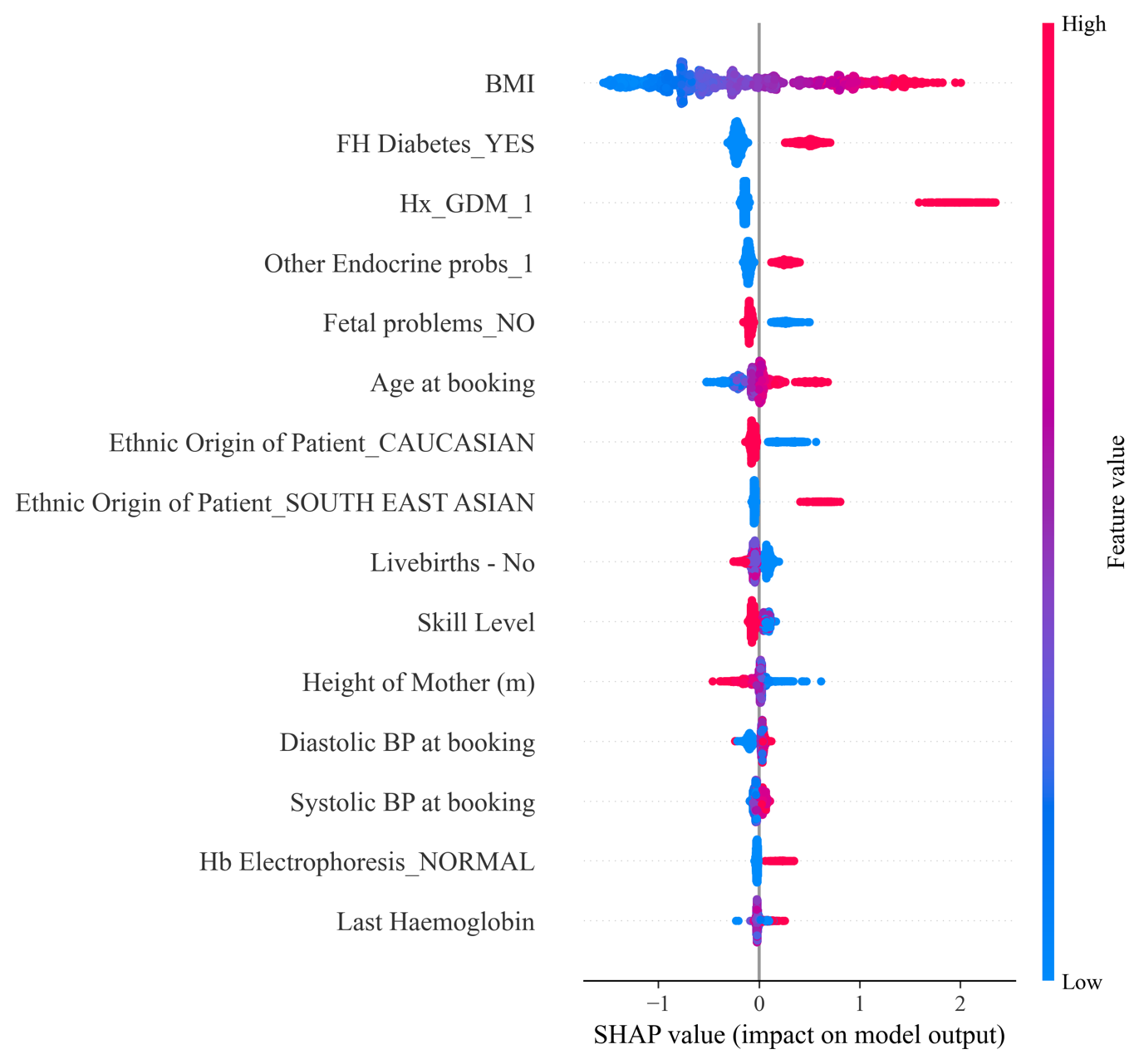

1. Feature Agnostic Model

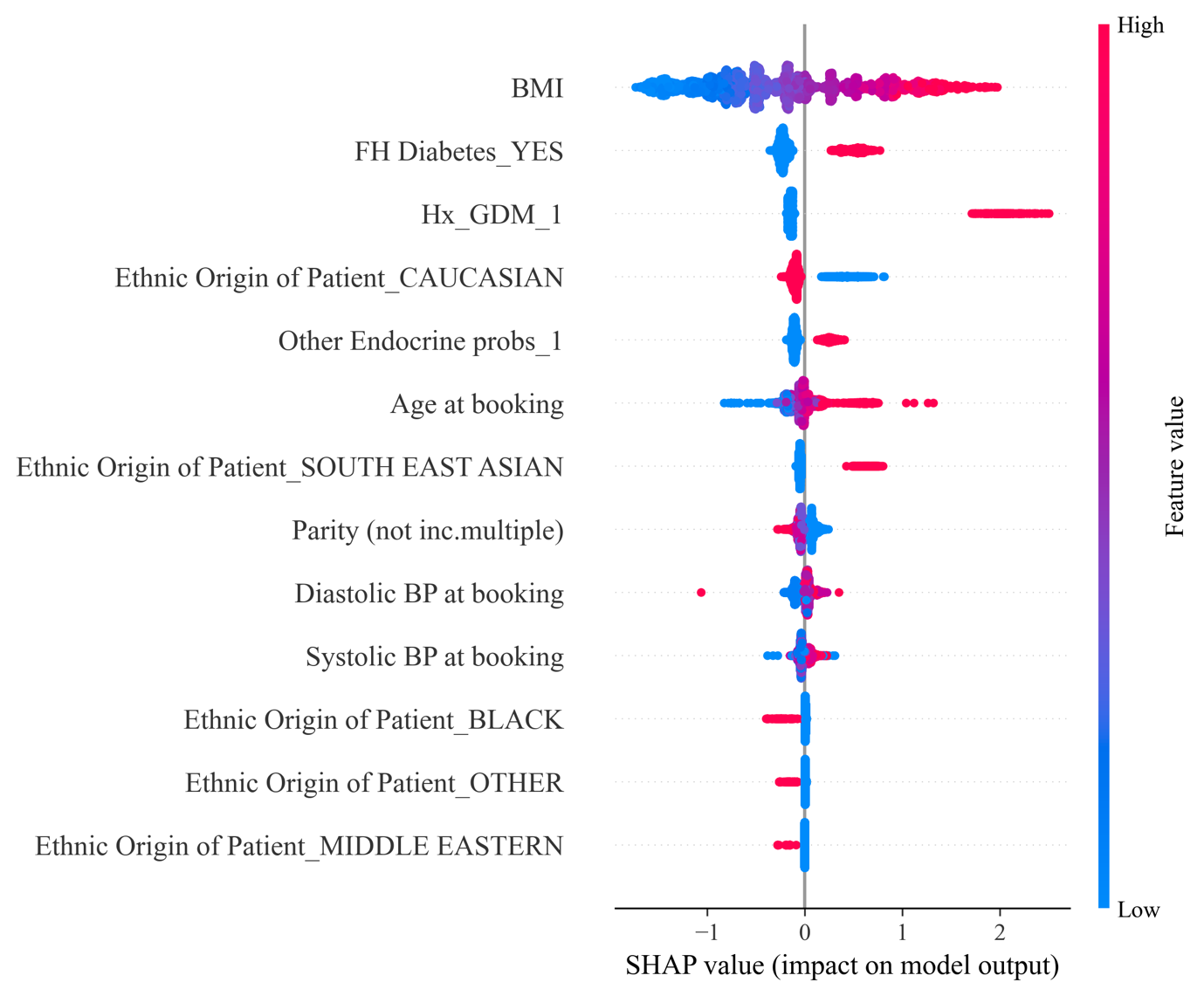

2. Feature Subset Model

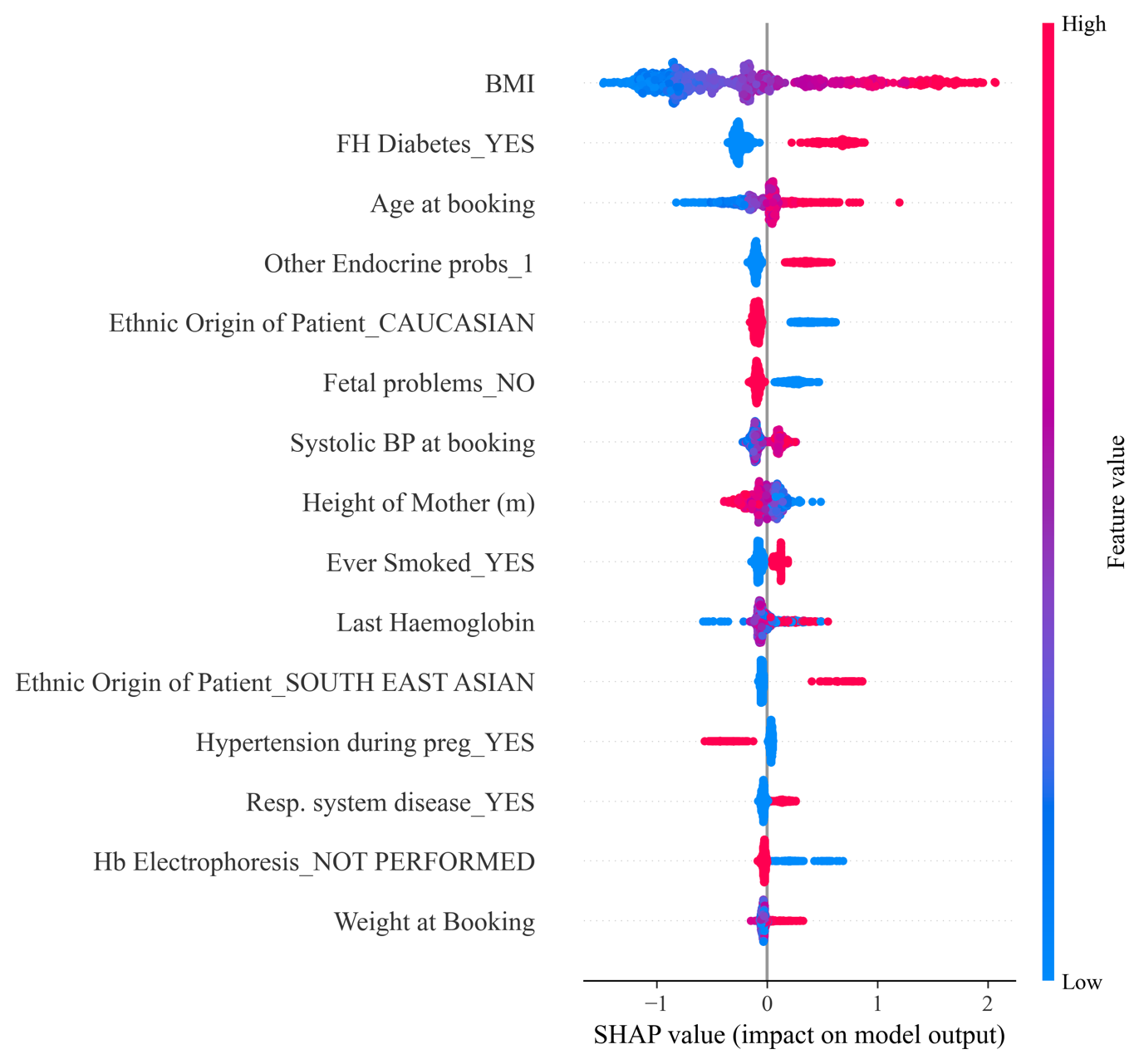

3. Nulliparous Model

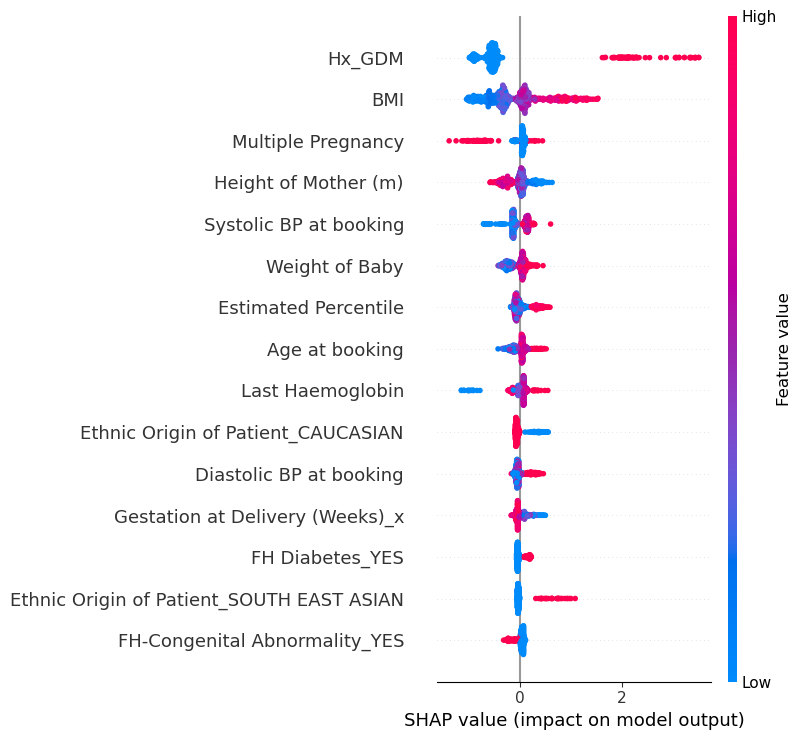

4. SPM Previous Pregnancy only.

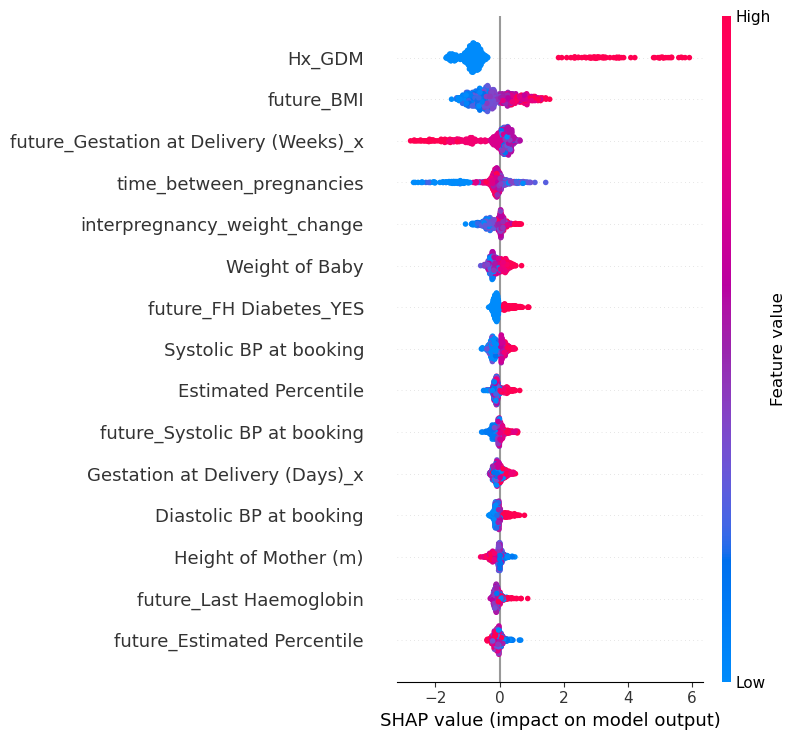

5. Including 1^st^ trimester data

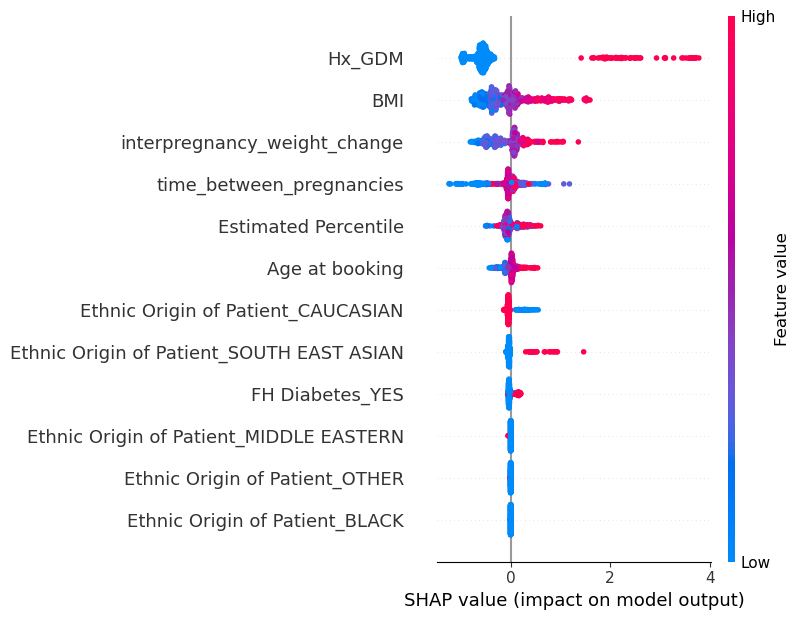

6. Sequential pregnancy subset model.
