## Supplementary Figure 3. for "Evaluation of Machine Learning Models for Early Prediction of Gestational Diabetes Using Retrospective Electronic Health Records from Current and Previous Pregnancies"

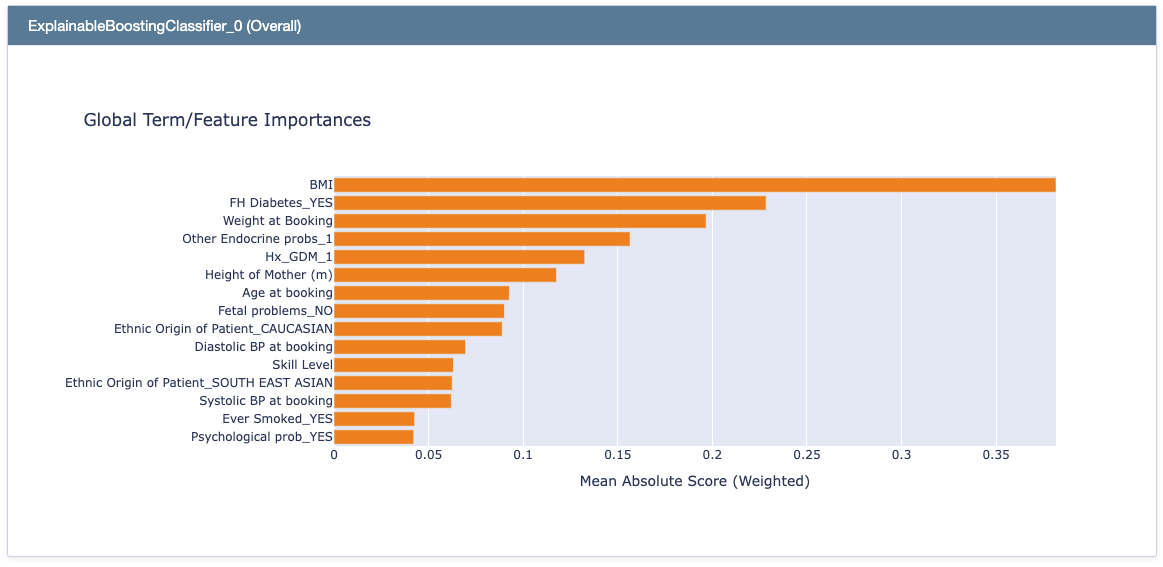

1. EBM Global Explanations: Feature Agnostic Model

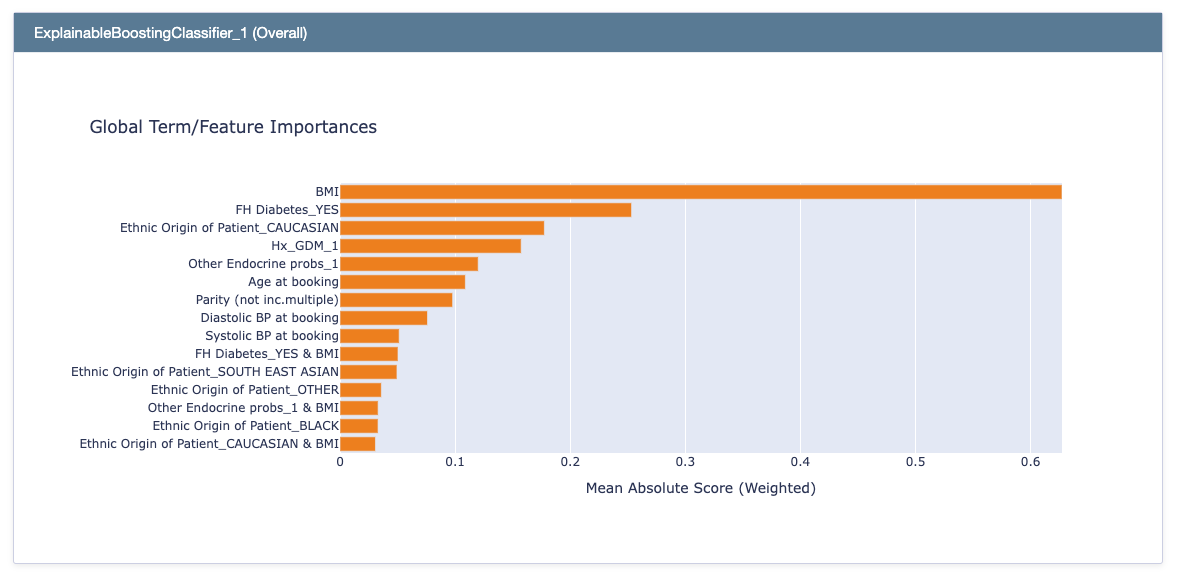

2. EBM Global Explanations: Feature Subset Model

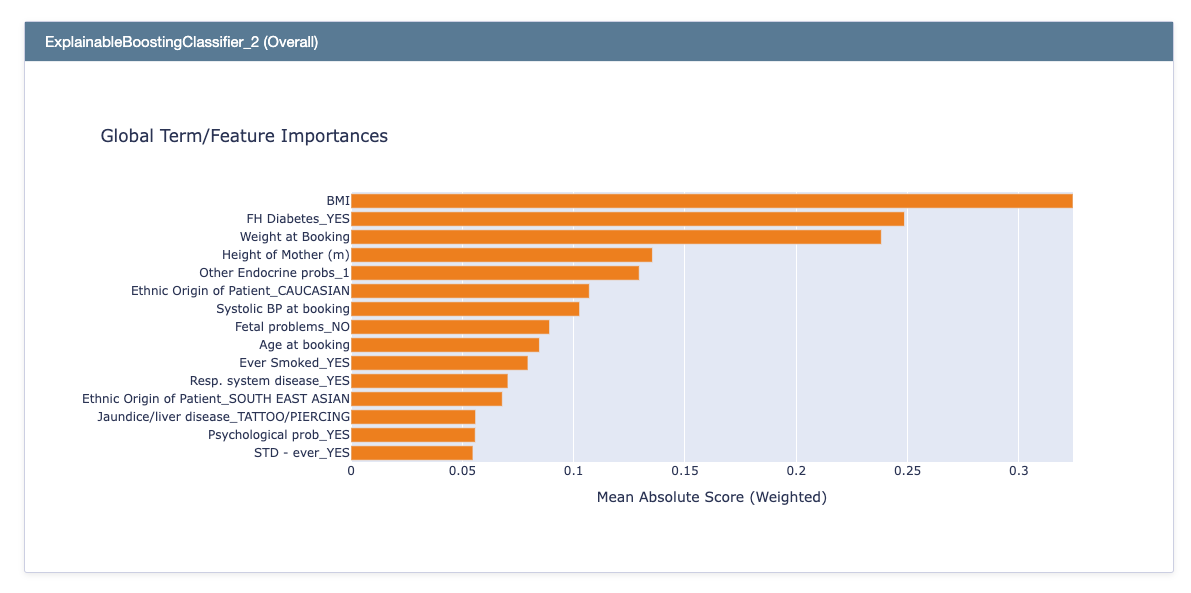

3. EBM Global Explanations: Nulliparous Model

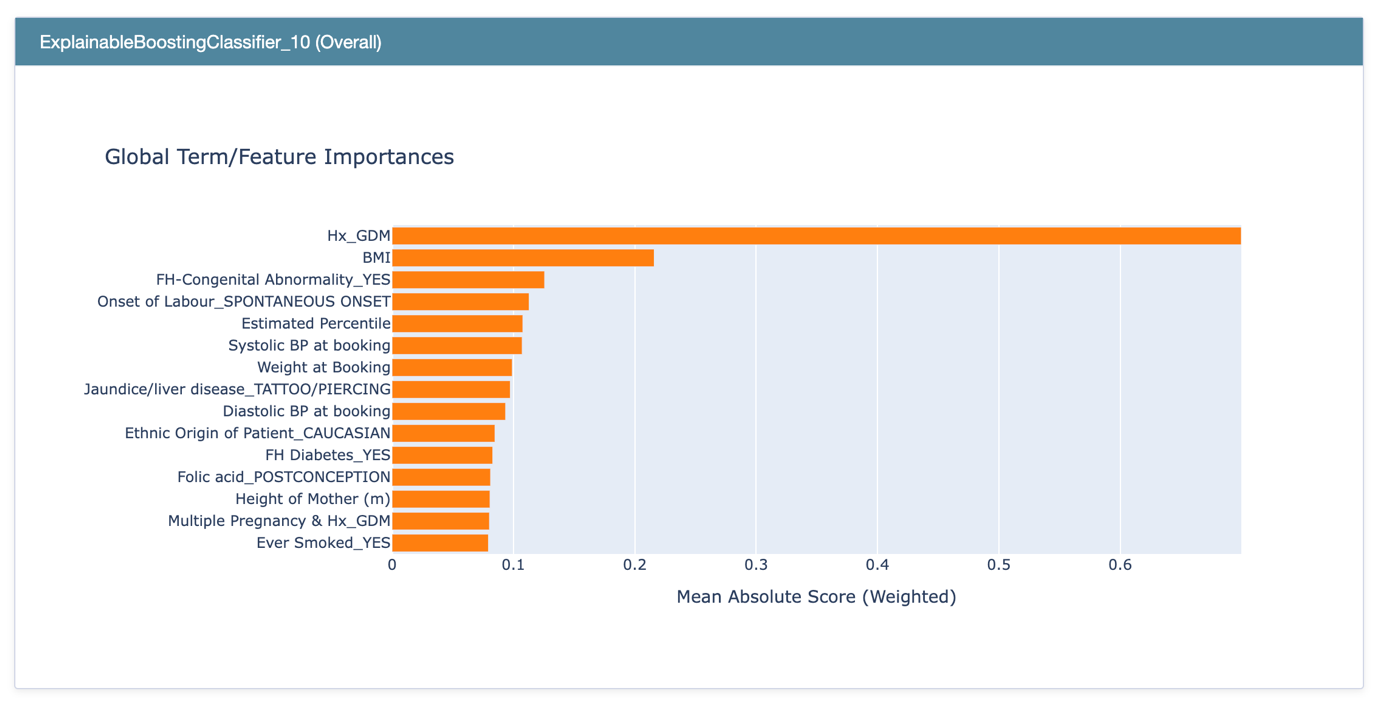

4. EBM Global explanations: SPM Previous Pregnancy only

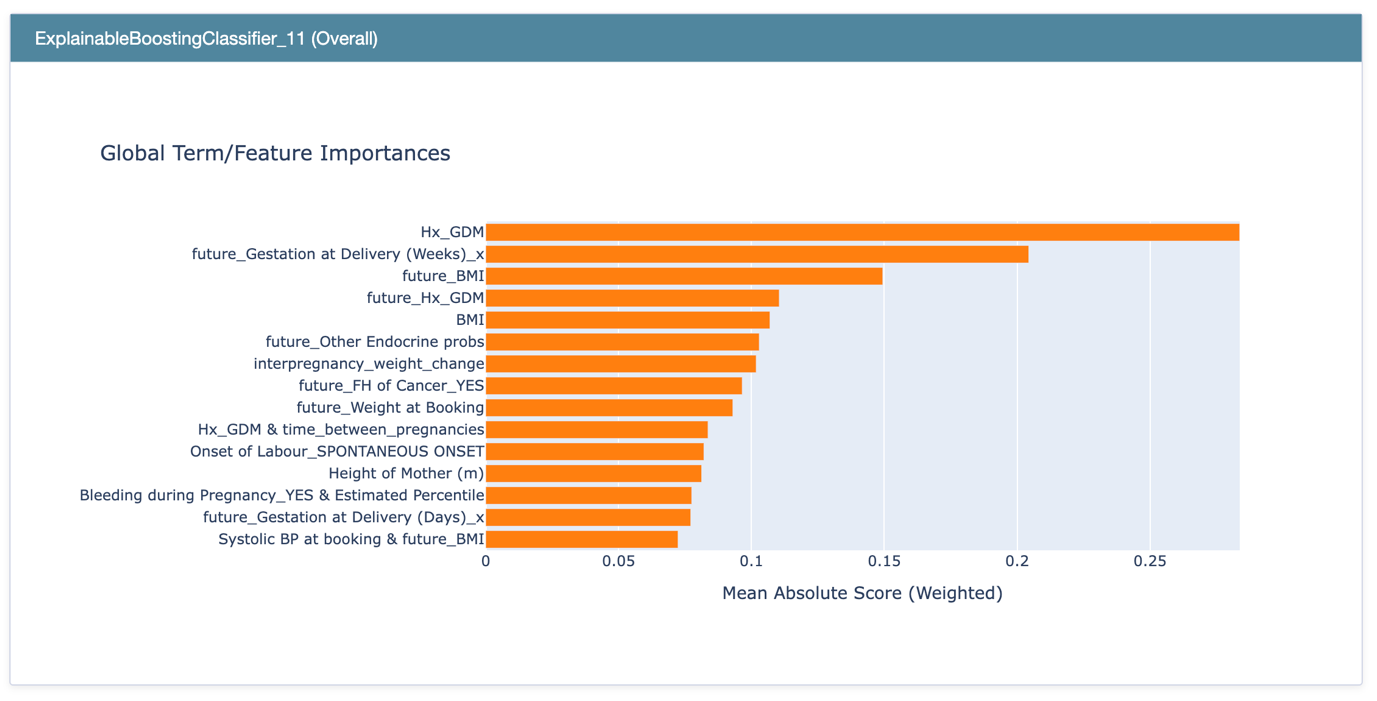

5. EBM Global explanations: SPM 1^st^ Trimester

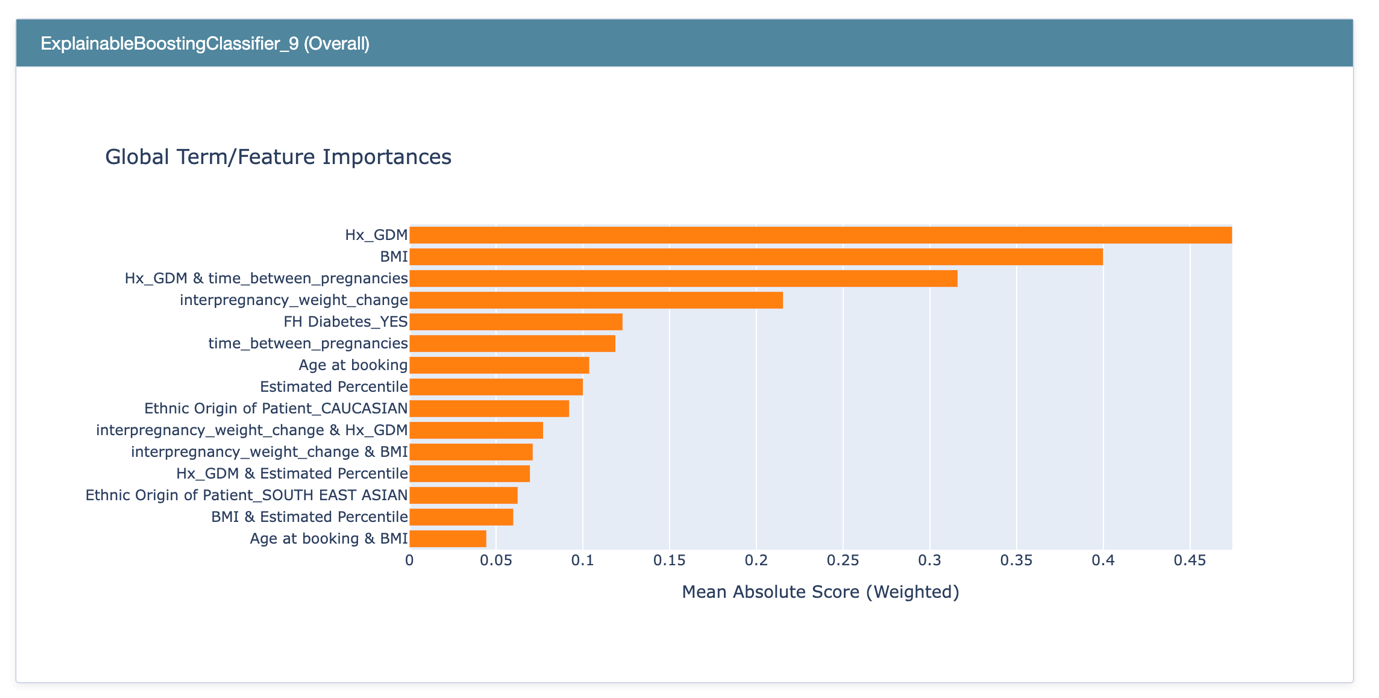

6. EBM Global explanations: SPM feature subset
