## Supplementary Figure 4. for "Evaluation of Machine Learning Models for Early Prediction of Gestational Diabetes Using Retrospective Electronic Health Records from Current and Previous Pregnancies"

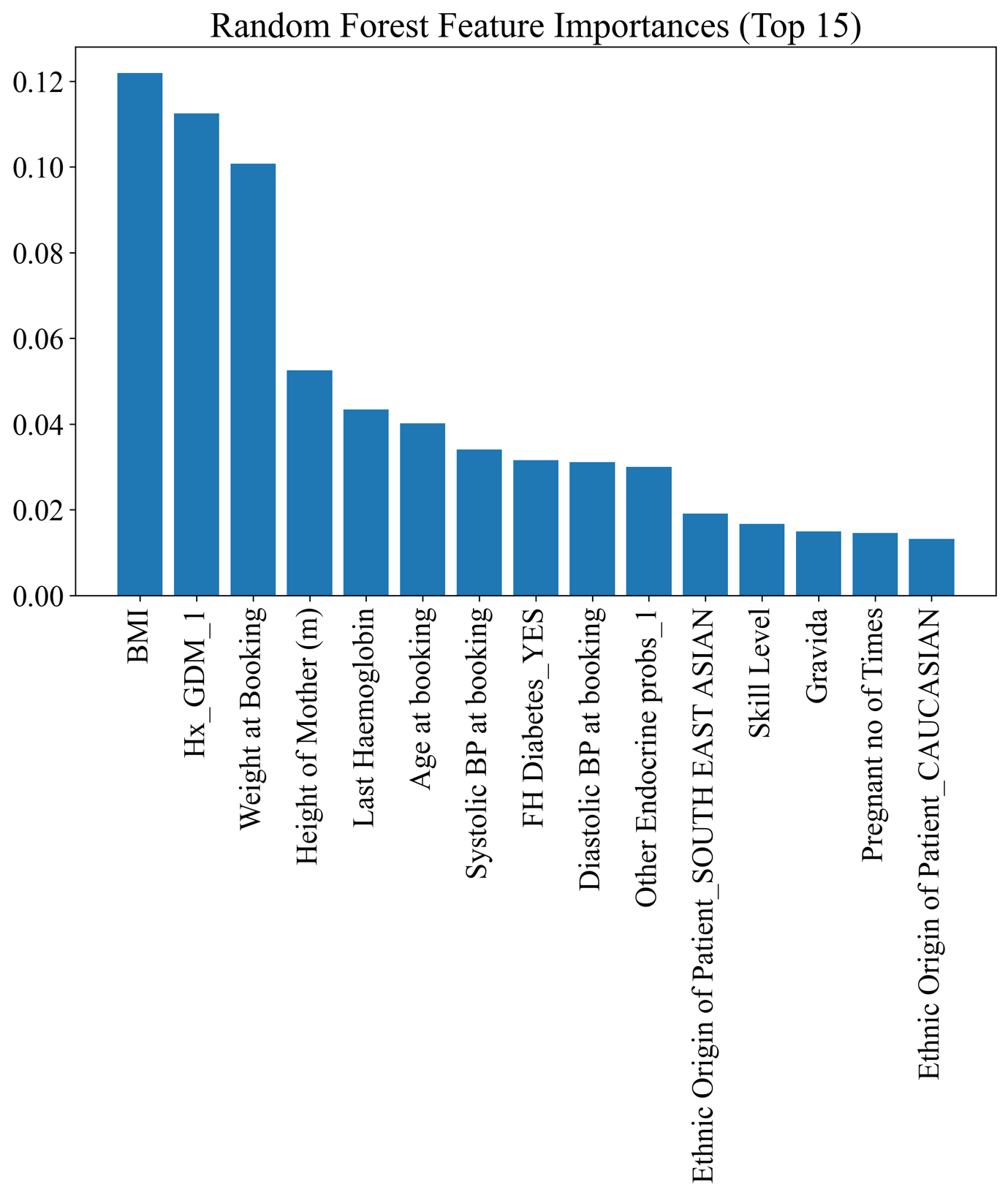

1. Random Forest Feature Importance: Feature Agnostic Model

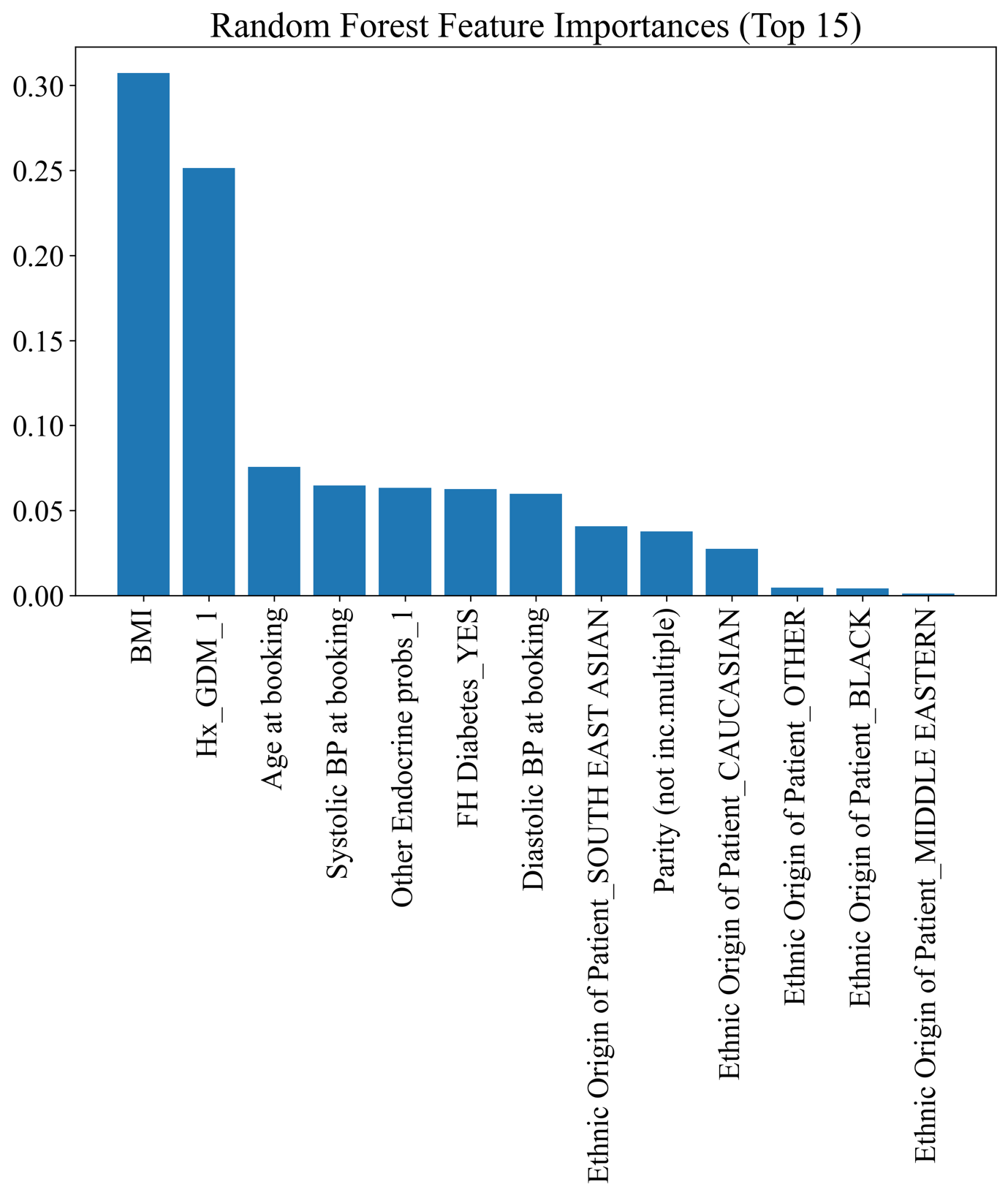

2. Random Forest Feature Importance: Feature Subset Model

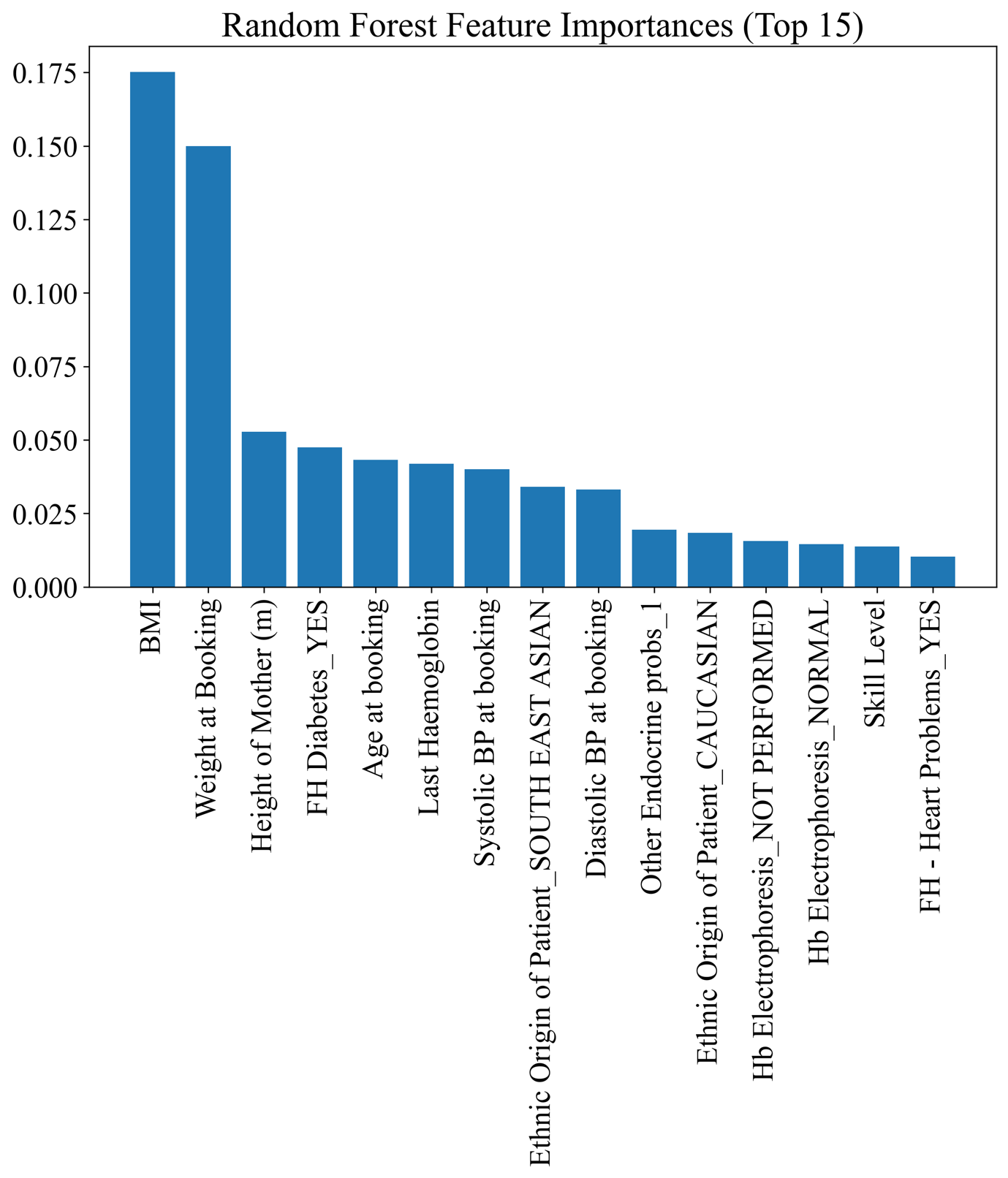

3. Random Forest Feature Importance: Nulliparous Model

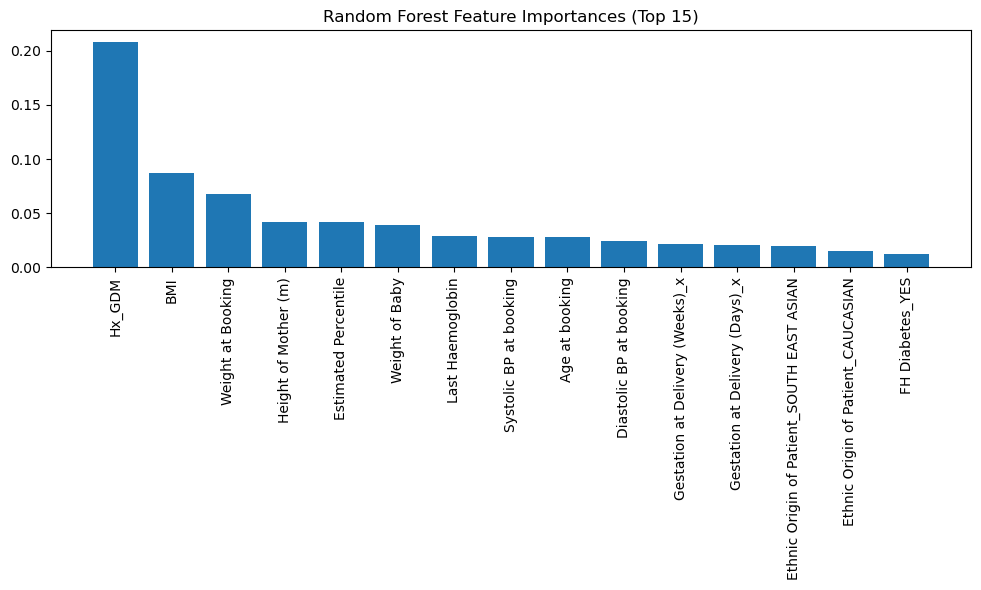

4. Random Forest Feature Importance: SPM Previous Pregnancy Only

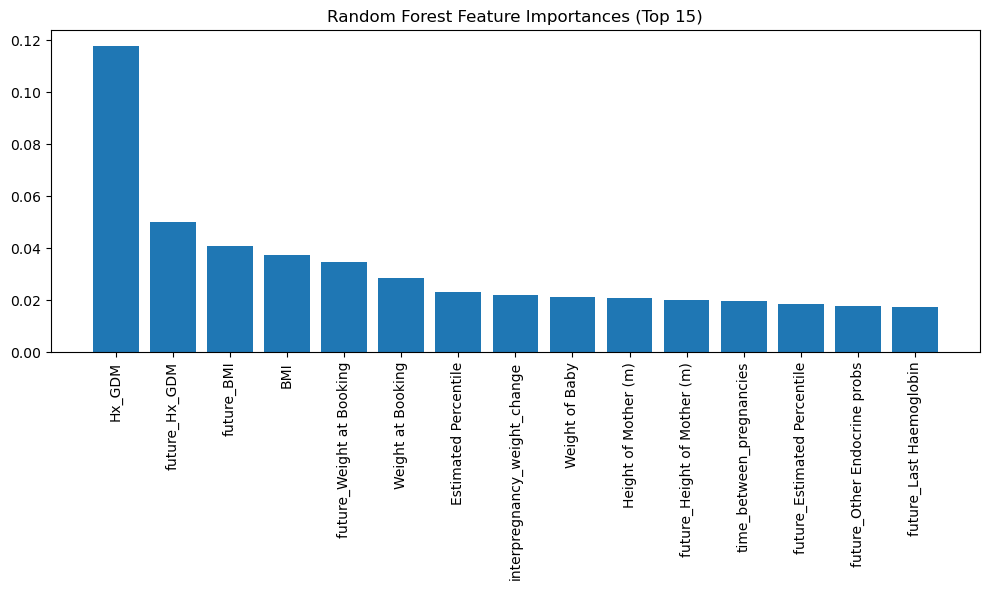

5. Random Forest Feature Importance: SPM with 1^st^ Trimester

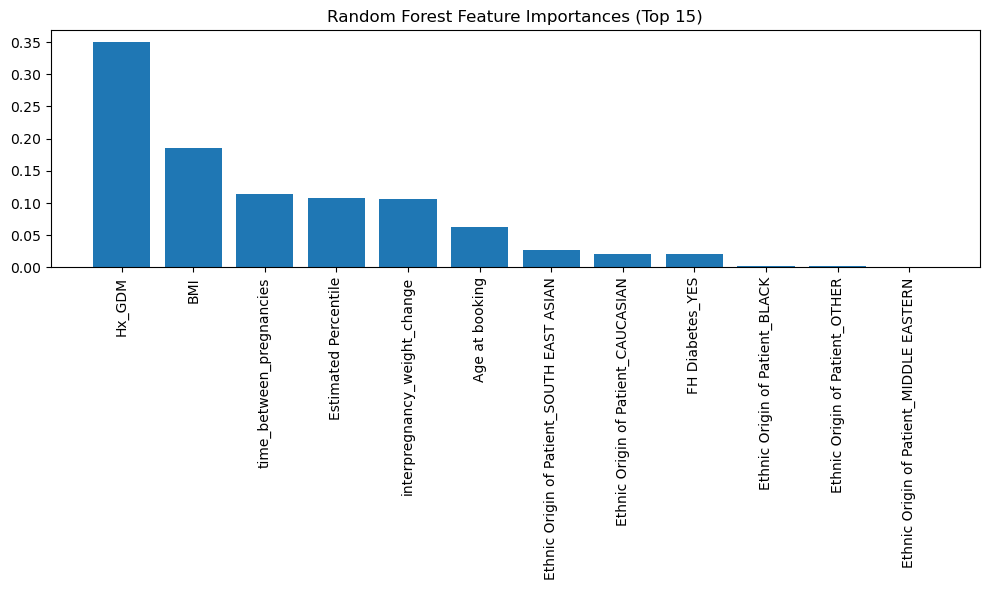

6. Random Forest Feature Importance: SPM Subset
