## Supplementary Figure 5. for "Evaluation of Machine Learning Models for Early Prediction of Gestational Diabetes Using Retrospective Electronic Health Records from Current and Previous Pregnancies"

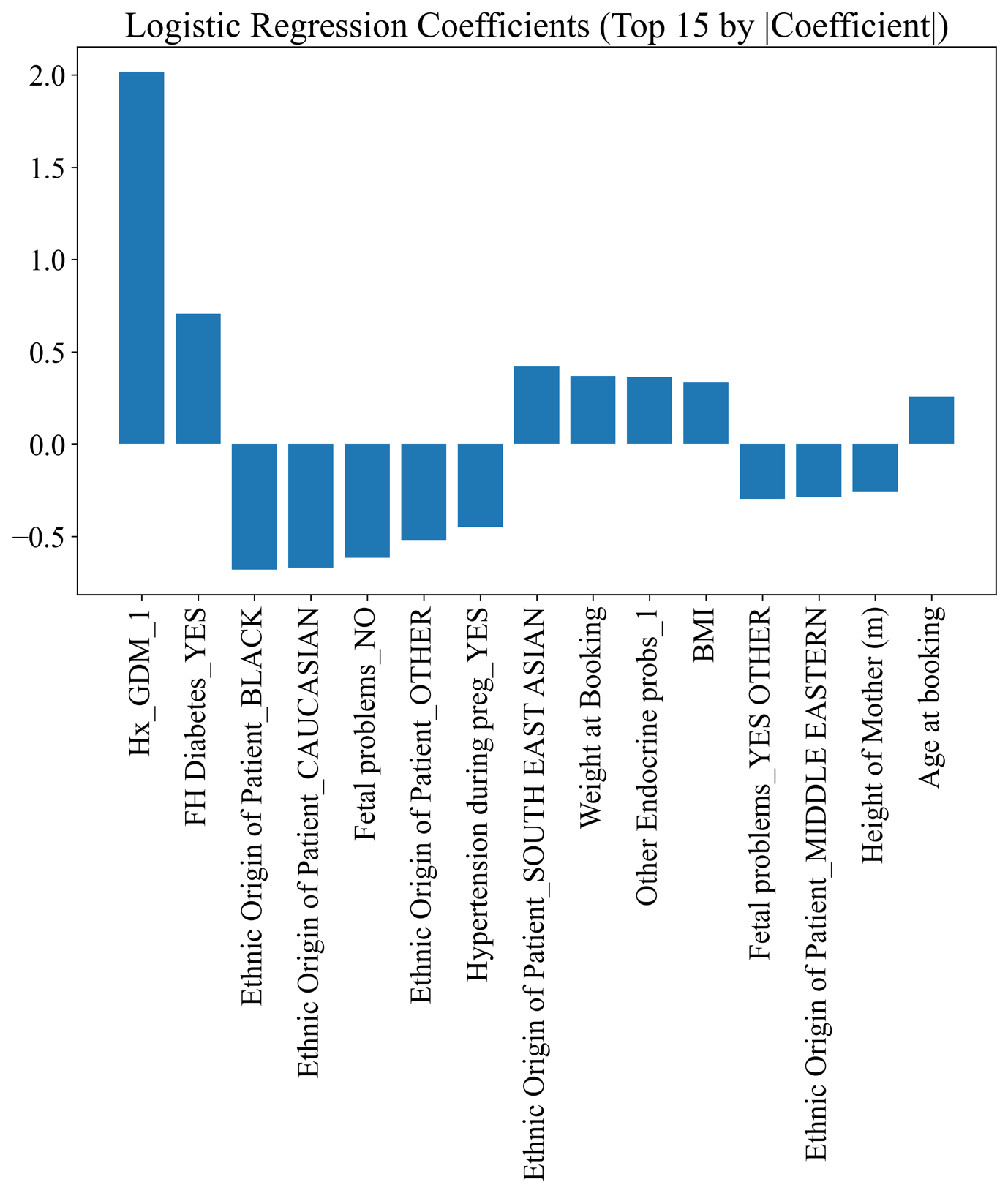

1. Logistic Regression Coefficients: Feature Agnostic Model

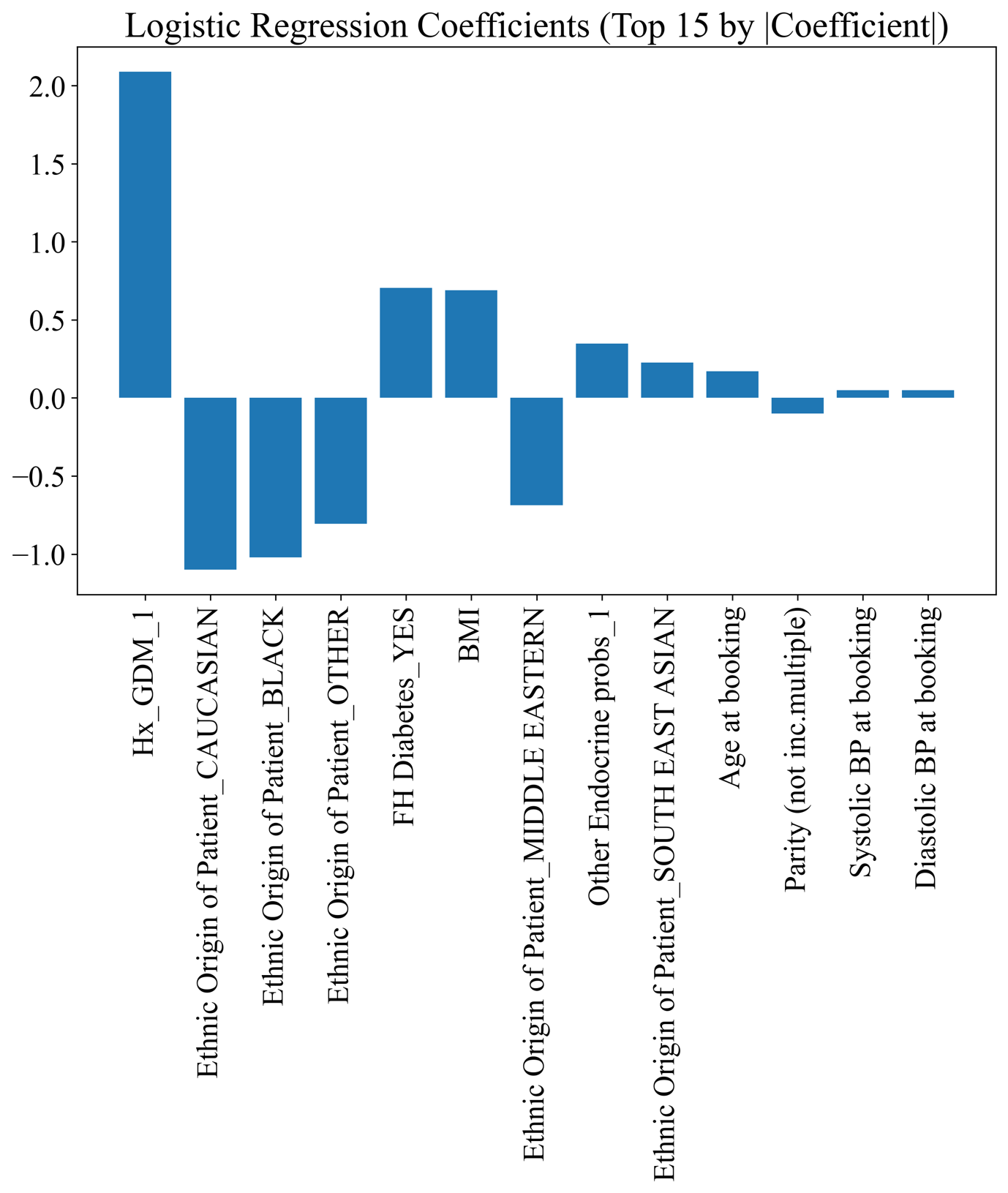

2. Logistic Regression Coefficients: Feature Subset Model

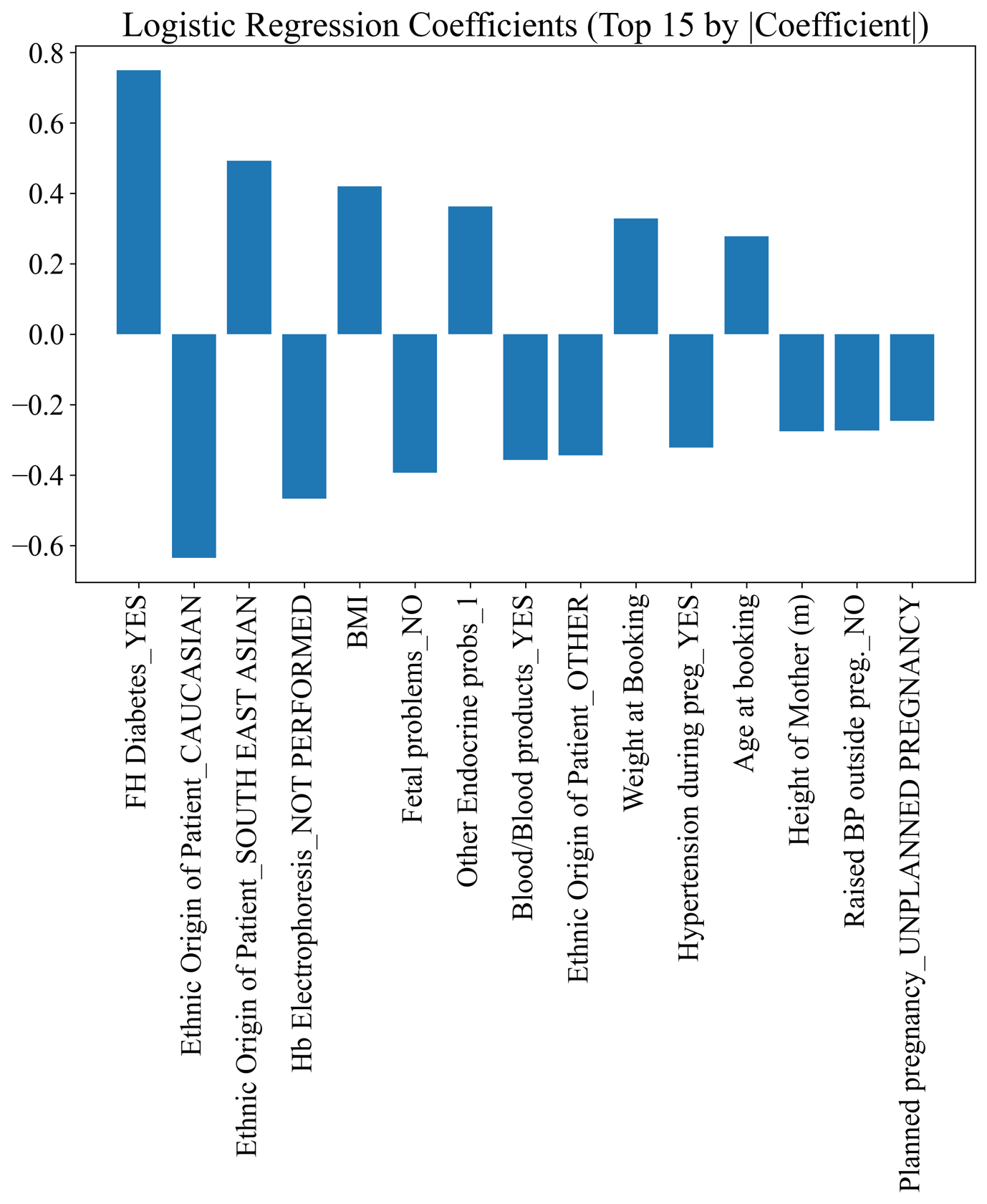

3. Logistic Regression Coefficients: Nulliparous Model

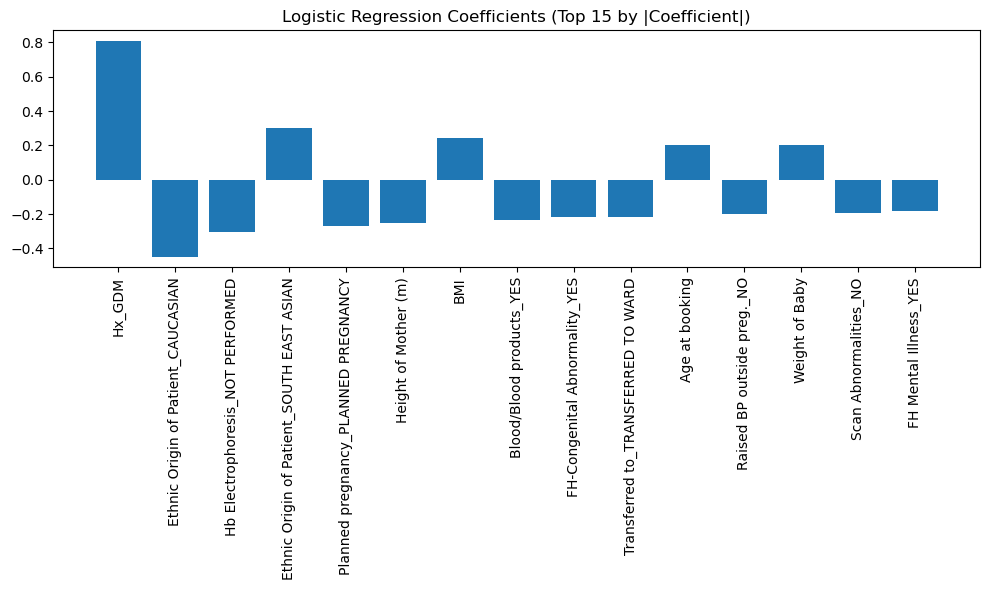

4. Logistic Regression Coefficients: SPM Previous Pregnancy Only

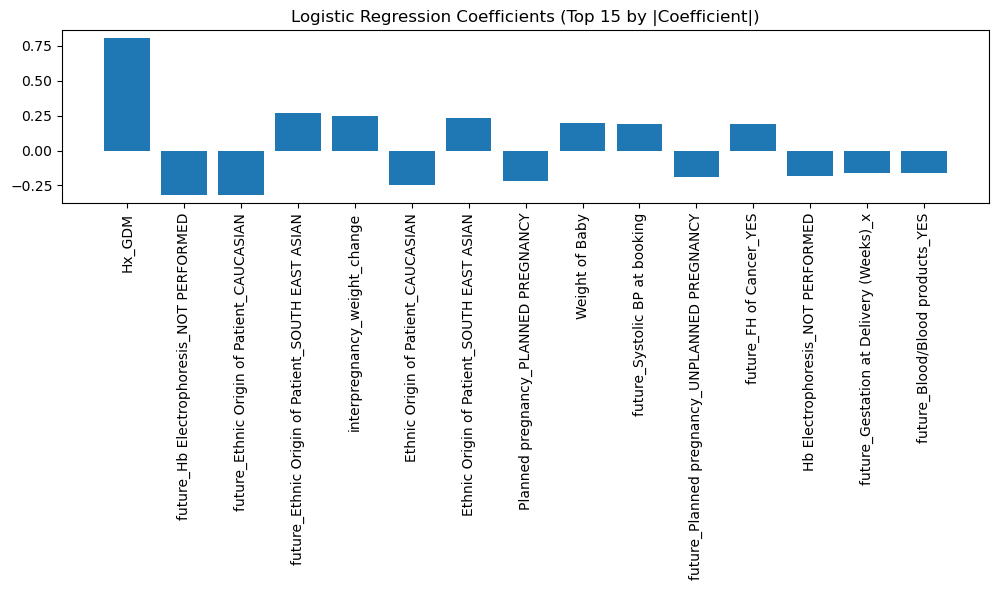

5. Logistic Regression Coefficients: SPM with 1^st^ Trimester

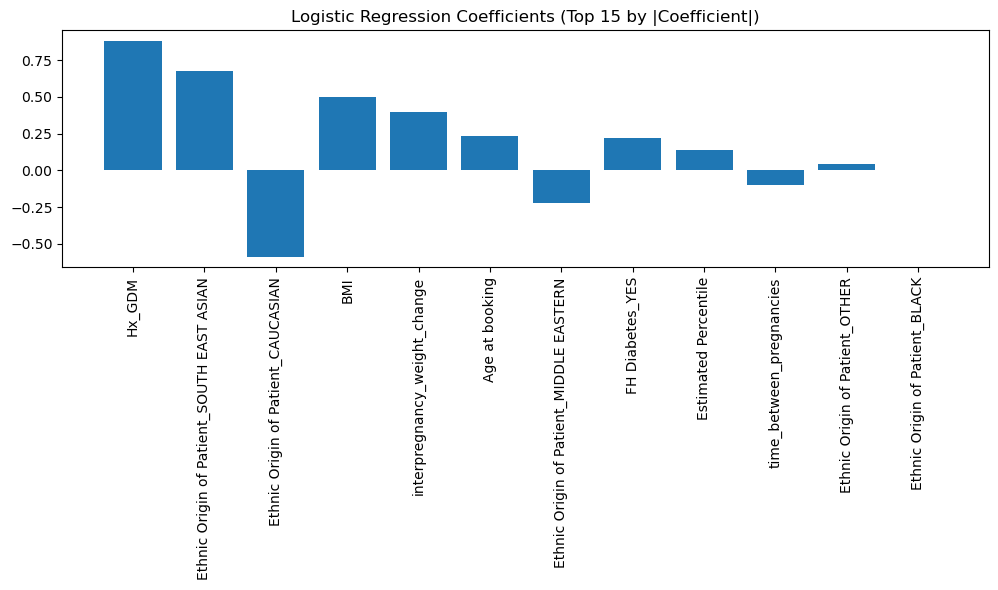

6. Logistic Regression Coefficients: SPM subset
